# Phylodynamic analysis of SARS-CoV-2 spread in Rio de Janeiro, Brazil, highlights how metropolitan areas act as dispersal hubs for new variants

**DOI:** 10.1101/2022.01.17.22269136

**Authors:** Alessandra P Lamarca, Luiz G P de Almeida, Ronaldo da Silva Francisco Junior, Liliane Cavalcante, Otávio Brustolini, Alexandra L Gerber, Ana Paula de C Guimarães, Thiago Henrique de Oliveira, Érica Ramos dos Santos Nascimento, Cintia Policarpo, Isabelle Vasconcellos de Souza, Erika Martins de Carvalho, Mario Sergio Ribeiro, Silvia Carvalho, Flávio Dias da Silva, Marcio Henrique de Oliveira Garcia, Leandro Magalhães de Souza, Cristiane Gomes Da Silva, Caio Luiz Pereira Ribeiro, Andréa Cony Cavalcanti, Claudia Maria Braga de Mello, Amilcar Tanuri, Ana Tereza R Vasconcelos

## Abstract

During the first semester of 2021, all of Brazil has suffered an intense wave of COVID-19 associated with the Gamma variant. In July, the first cases of Delta variant were detected in the state of Rio de Janeiro. In this work, we have employed phylodynamic methods to analyze more than 1,600 genomic sequences of Delta variant collected until September in Rio de Janeiro to reconstruct how this variant has surpassed Gamma and dispersed throughout the state. After the introduction of Delta, it has initially spread mostly in the homonymous city of Rio de Janeiro, the most populous of the state. In a second stage, dispersal occurred to mid- and long-range cities, which acted as new close-range hubs for spread. We observed that the substitution of Gamma by Delta was possibly caused by its higher viral load, a proxy for transmissibility. This variant turnover prompted a new surge in cases, but with lower lethality than was observed during the peak caused by Gamma. We reason that high vaccination rates in the state of Rio de Janeiro were possibly what prevented a higher number of deaths.

**Impact statement:** Understanding how SARS-CoV-2 spreads is vital to propose efficient containment strategies, especially when under the perspective of new variants emerging in the next year. Still, models of SARS-CoV-2 dispersal are still largely based in large cities from high-income countries, resulting in an incomplete view of the possible scenarios consequent of a new variant introduction. The work improves this discussion by reconstructing the spatio-temporal dispersal of Delta variant since its introduction in Rio de Janeiro, a densely populated region in South America. We also analyzed the epidemiological outcome of this spread, with a decrease in lethality rate uncommon to the observed in other countries.

**Data summary:** Four supplementary figures, one supplementary table and one supplementary file are available with the online version of this article. Raw short reads of the newly sequenced genomes are available at SRA-NCBI database (https://www.ncbi.nlm.nih.gov/sra) under the BioProject PRJNA774631 and the assembled genomes are deposited at GISAID database (https://www.gisaid.org/) under the accession numbers listed in Table S1. Other genomic sequences used in the analyses are listed in Table S2. Epidemiological data for the state of Rio de Janeiro was obtained from https://www.saude.rj.gov.br/informacao-sus/dados-sus/2020/11/covid-19.

## Introduction

The state of Rio de Janeiro, Brazil, has suffered an intense wave of COVID-19 in the first semester of 2021, mostly associated with the Gamma variant (P.1 and its sublineages in PANGO classification) (1). With an estimated population of more than 17.4 million people, the state had a maximum of 31,440 COVID-19 cases and 2,000 deaths from the disease in a week. From mid-May onwards, this number diminished to 13,232 cases and 609 deaths in a week at the end of June. The first cases of Delta variant in Rio de Janeiro were then detected in July, leading to the first communitary outbreak of the variant in Brazil (2). Since then, Delta has spread across different Brazilian states and became the dominant variant circulating in the country (2–5).

The state of Rio de Janeiro is the third most populous state of Brazil and the second most dense (399,15 hab/km^2^). It has the fourth-highest Human Development Index in the country and almost half of this population lives in the homonymous city of Rio de Janeiro. Previous works have shown that population density, proximity to international airports and city infrastructure influence the speed and routes in which SARS-CoV-2 spreads (6–13). Understanding how this virus disperses within and between urban regions is crucial to elaborate efficient and science-driven public health policies to contain the pandemic. The continued genomic surveillance of SARS-CoV-2 lineages conducted in the state of Rio de Janeiro (14) is a rare opportunity to investigate and monitor the dispersal of the Delta variant since its first introduction. In this work, we employed phylodynamic methods to analyze more than 1,600 Delta genomes collected between July and September and inferred the dispersal patterns and routes of the variant in the state.

## Material and Methods

### Epidemiological analyses

Daily number of confirmed COVID-19 cases, deaths and people vaccinated were obtained from the National Immunization Program (Programa Nacional de Imunização, PNI), esus-VE and SIVEP-Gripe databases through the COVID-19 portal (https://www.saude.rj.gov.br/informacao-sus/dados-sus/2020/11/covid-19) maintained by the Secretary of Health of the state of Rio de Janeiro. The lethality of COVID-19 in each age group was calculated by dividing the number of deaths by the number of cases that occurred in each week. Population size of each age group used to calculate the proportion of completed vaccinated (second dose of BNT162b2, ChAdOx1 or CoronaVac vaccines or a single dose of Ad26.CoV2.S vaccine) was obtained from the COVID-19 panel maintained by the Secretary of Health of the state of Rio de Janeiro (https://painel.saude.rj.gov.br/monitoramento/covid19.html). The number of tests for SARS-CoV-2 infection in Rio de Janeiro was considered consistent throughout 2021 (15).

### Sequencing

We randomly selected samples of SARS-CoV-2 obtained from a pool of nasopharyngeal swabs collected by the Noel Nutels Central Laboratory (LACEN-RJ) and Unidades de Apoio ao Diagnóstico da Covid-19 (UNADIG-RJ) to diagnose COVID-19 in residents of the state of Rio de Janeiro between June 6th and September 12th of 2021. The genetic material was extracted at the Laboratory of Molecular Virology at Universidade Federal do Rio de Janeiro (LVM-UFRJ) using MagMAX Viral/Pathogen Nucleic Acid Isolation kits KingFisher automatic platform. The cDNA was annealed with 8,5 µl of viral RNA extracted from each sample and the Illumina COVIDSeq Test (Illumina) was used according to the manufacturer’s protocol to construct libraries at the DFA/LNCC Genomics Unit. An aliquot of 5 µl of each library was combined and purified and we used the TapeStation (Agilent) system for quality control. The NextSeq 500/550 Mid Output Kit v2.5 (300 Cycles) was used to generate reads of 2×149 bp in the NextSeq (Illumina). The DRAGEN COVID Lineage v3.5.1 pipeline was used for sequence analysis, consensus building, and variant calling. The study was approved by the Ethics Committee (30161620.0.1001.5257 and 34025020.0.0000.5257). All newly sequenced and assembled genomes are publicly available at GISAID (Table S1) and SRA-NCBI (BioProject PRJNA774631) databases.. Finally, we used the PangoLEARN v1.2.103 model database to classify the lineage of our newly assembled genomes and removed from the dataset all non-Delta sequences. The remaining genomes (*n* = 1334) belonged to samples obtained from patients with ages ranging from 1 to 100 years old, 44.6% being male and 55.4% female, and residing in 85 (of 92) cities of Rio de Janeiro (Fig. S1).

### Evolutionary analyses and phylogeographic reconstruction

We obtained from the GISAID database all genomes classified as lineage B.1.617.2 or its sublineages (Delta variant) that were submitted until October 1^st^ and collected in the state of Rio de Janeiro, Brazil. We removed from this dataset any sequence that did not contain complete date information, was shorter than 29,000 pb, had more than 1% of Ns, more than 0,05% of unique amino acid mutations (“high-coverage” in GISAID) or didn’t have the locality/city information. We added our newly sequenced genomes collected in the state, totalling 1602 genome sequences. These sequences were independently aligned to the WH01 (EPI ISL 406798) sequence from Wuhan, China using MAFFT v.7 (16) with the --addfragments option. All genomes’ 3’ and 5’ ends were trimmed using the SeqKit toolkit (17). Maximum likelihood trees were then inferred with IQTREE2 (18) using the GTR+F+I+G4 model selected by the ModelFinder algorithm (19) with “-mset mrbayes” option and 1000 ultrafast-bootstrap replicates (20). The outgroup (WH01) was then removed and root-to-tip distances were calculated using TempEst. Using Cook’s distance, we removed from the alignment sequences that most influenced the correlation between root-to-tip and sampling date. This final dataset contained 1512 sequences and was used in all subsequent analyses. A new maximum likelihood tree was generated using the same parameters as before. This new tree was used as a fixed topology in the subsequent analyses (divergence dating and phylogeography).

We then moved to investigate whether the outbreak of Delta in Rio de Janeiro was originated by one or multiple introductions of the variant in the state. As representative sequences of the global diversity, we used the genomes belonging to the Delta clade in the SARS-CoV-2 global phylogeny (https://github.com/nextstrain/ncov) generated through the Nextstrain pipeline (21). We further tested the origin and monophyly of lineages AY.99.1 and AY.99.2 by obtaining from GISAID the 20 oldest sequences of each of these lineages that were not from the state of Rio de Janeiro (ten global sequences and ten from Brazil) and adding them to the background dataset. The background genomes were combined to our dataset from Rio de Janeiro, aligned to the WH01 genome and the 3’ and 5’ ends of the new sequences were trimmed. A maximum likelihood tree was inferred as described before.

Divergence dates were estimated using BEAST v.1.10.4 employing the strict clock model with a uniform prior (max substitution rate of 1e-3, min 6e-8), the GTR substitution model with empirical frequencies and the Gamma+Invariant sites model, and the Coalescent-Exponential Growth tree prior. The MCMC was run through a chain of 100,000,000 with sampling every 10,000th. The consensus tree was summarized with Tree Annotator and used as the fixed topology and branch lengths for the phylogeographic reconstruction of the spread of SARS-CoV-2 in Rio de Janeiro. To accomplish this, we used the BEAST software and the Relaxed Random Walk model with Cauchy’s distribution on coordinates randomly selected within each sample’s collection city. The MCMC ran through a chain of 100,000,000 with sampling every 10,000th. Dispersal routes were extracted from the consensus tree using the *seraphim* package and plotted using the *ggplot2* package, both in R software. Base map was obtained from Instituto Brasileiro de Geografia e Estatística (IBGE, https://www.ibge.gov.br/geociencias/organizacao-do-territorio/malhas-territoriais/15774-malhas.html?=&t=downloads).

Finally, we also inferred divergence dates and dispersal using TreeTime (22) to confirm our findings. For divergence dating, we kept the root of the maximum likelihood tree previously inferred (--keep-root), used the “skyline” coalescent model (--coalescent skyline) and assumed correlation between root-to-tip distances (--covariation). For the phylogeographic analysis, we used the “mugration” model inference with the sampling city as the discrete character to be analyzed. The results were analyzed and plotted using *ggtree* and *ggplot* packages in R.

## Results and Discussion

On June 16th of 2021, the first case of the Delta variant was confirmed in Rio de Janeiro, and a fast increase of the variant’s frequency in the state was reported (2). At the time, Rio de Janeiro was on the decline of COVID-19 cases caused by the outbreak of Gamma variant in the country (14,23). The fast spread of Delta observed in Rio de Janeiro incited concern for a new COVID-19 surge. Analyzing epidemiological data publicly available, we observed that there was indeed an increase of cases in the state (Fig. 1A), with a maximum of 23,086 cases in a week at the beginning of August (week 32). This number corresponds to approximately 73% of cases from the first peak (end of March, week 11) of the bimodal wave caused by Gamma and is very similar to the number of cases in the second peak (early May, week 19). However, the increase in deaths caused by this new variant was much smaller, with the 890 lives lost (week 33) being less than half of the maximum observed in the first semester of 2,021 (*n* = 2,016, week 11; Fig. 1B). While recent research suggests that Delta causes a higher number of hospitalizations and aggravated symptoms than previous lineages (24,25), we believe the advances in vaccination throughout the state has reduced the lethality from 7,6% (late March, week 13) to the approximately 3% at the peak of the Delta surge. Interestingly, in the last four weeks of the temporal series analyzed, there was an increase in lethality (max 6,5%) even though cases and deaths were decreasing. This increase was observed in all age groups above 40 years and infants until 4 years (Fig. 1C) and we could not identify the factor determining it. Age group proportions in the number of cases was similar throughout 2021 while the distribution in deaths saw first a decrease in the proportion of older age groups followed by normalization, possibly caused by health authorities prioritizing vaccination in such groups first (Fig. S2, Fig. 1D).

**Fig. 1.**
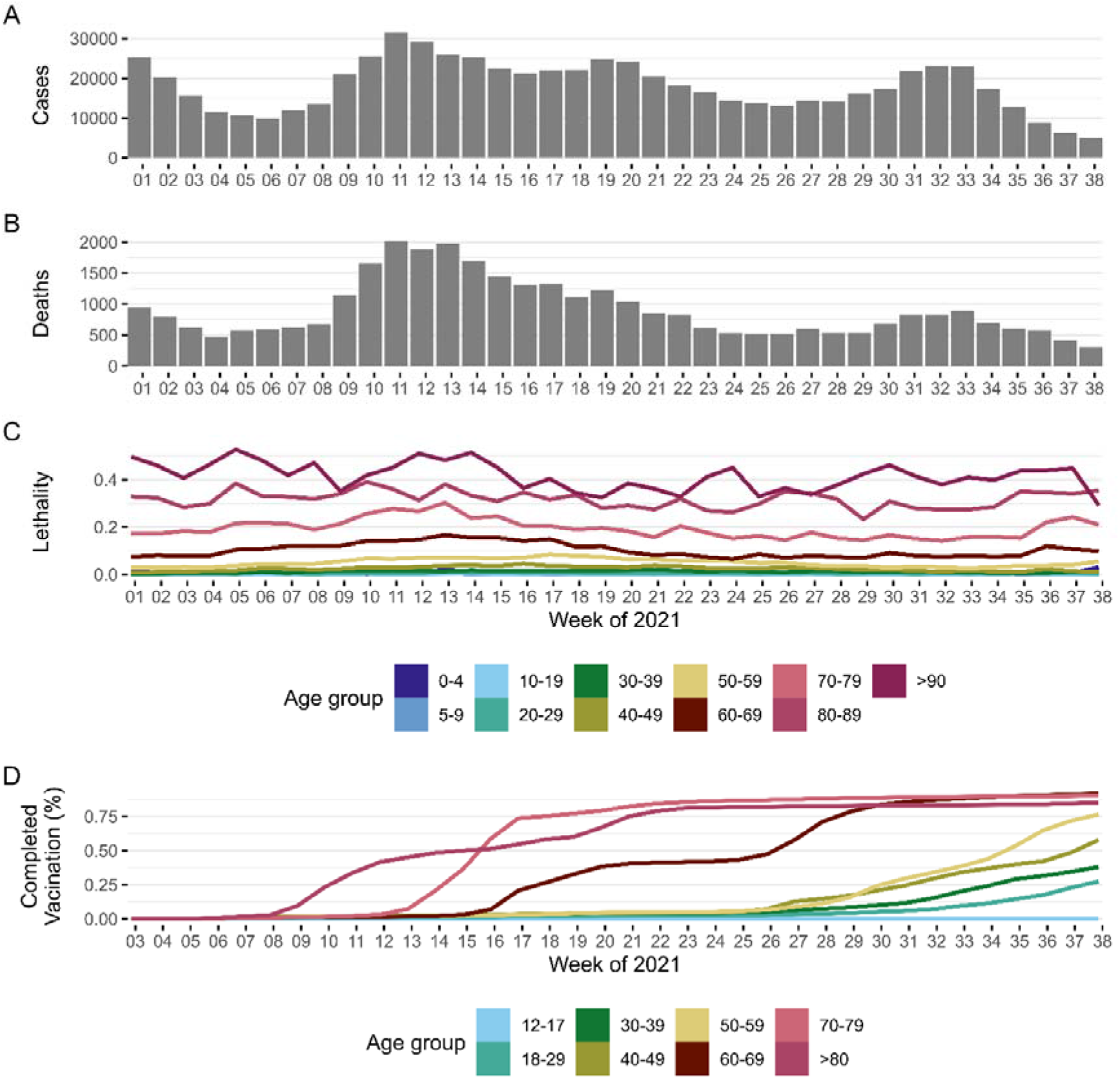
Epidemiological metrics of SARS-CoV-2 pandemic in the state of Rio de Janeiro in each week of 2021. (A) Number of cases, (B) number of deaths, (C) lethality rate in each age group and (D) completed vaccination (single dose or second dose) in each age group. Colors correspond to each age group analyzed.

This surge is concomitant to the jump in frequency of Delta from 67% (n = 156/232) to 89% (n = 130/146) of the samples sequenced in this work (Fig. 2, weeks 30-31). Substitution of Gamma, Alpha (B.1.1.7) and Beta (B.1.351) by Delta has been reported around the globe, being the dominant variant until the emergence of Omicron (B.1.1.529 and sublineages). While the replacement of Gamma by Delta occurred at an alarming rate, we detected at least 12 weeks of both variants co-circulating in the state. The presence of both lineages allows for coinfection events, which are the requirement for variant recombination. Events of recombination between variants were detected not only in the state of Rio de Janeiro during the period analyzed (26), but also in several countries and times (27–29). While the role of recombination in originating new Variants of Concern or Variants of Interest (30,31) is still being discussed, genomic surveillance should be enforced during periods of lineage replacement.

**Fig. 2.**
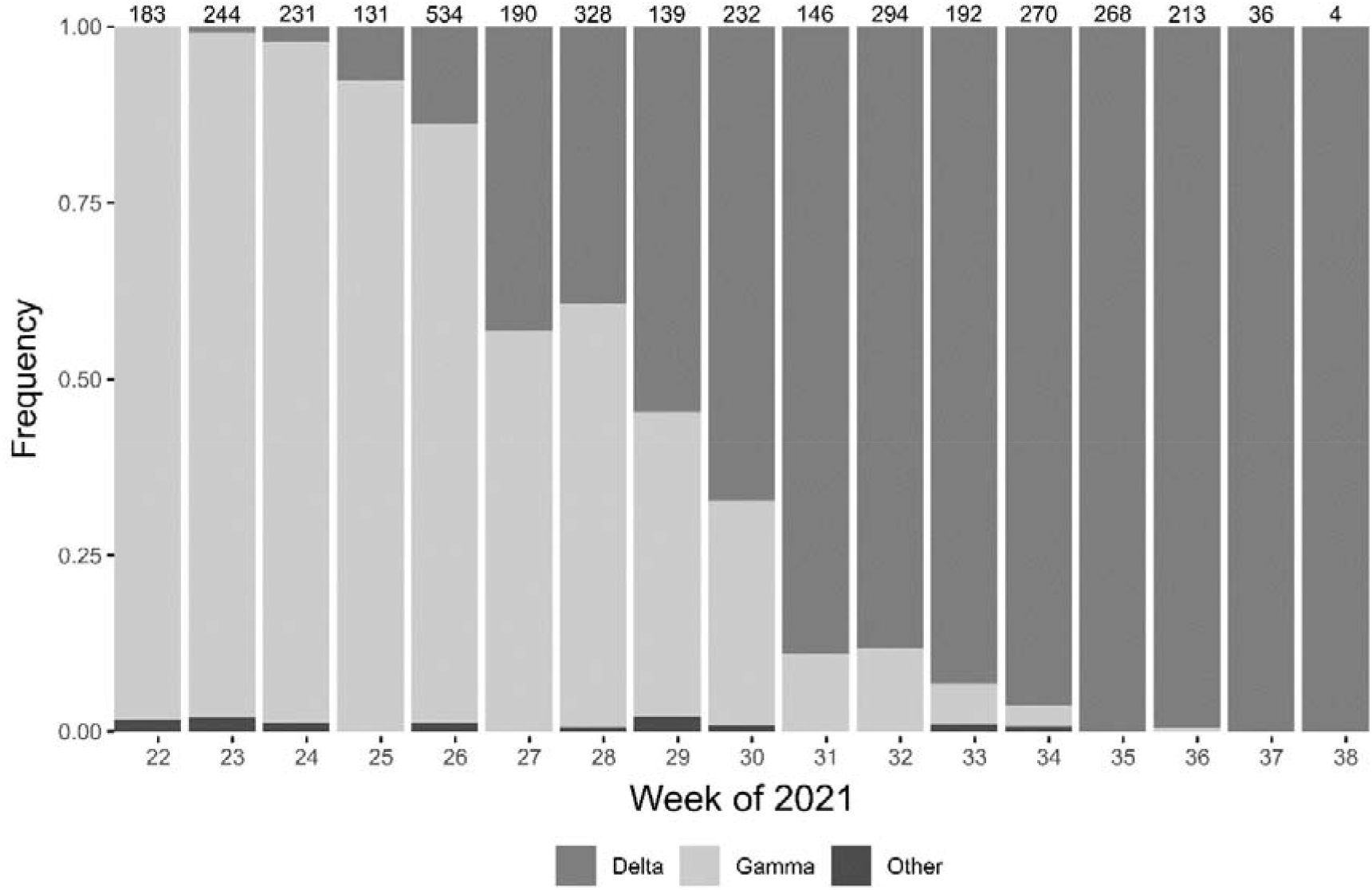
Relative frequency of variants in the state of Rio de Janeiro between June 1^st^ and 21^th^ September of 2021. Numbers on the top of each bar represent the total of sequences from the corresponding week.

To investigate how Delta surpassed Gamma in the state of Rio de Janeiro, we compared the relative quantification of viral load (RQVL = 2^−ΔCT^) of samples infected by variants circulating in the period analyzed in this study. We observed that samples from Delta exhibited significantly higher RQVL values than Gamma (Wilcoxon test, p.value = 0.01; Fig. S3a). While a few samples from Gamma accumulated more than 90% of the circulating viruses, more samples from Delta were required to represent the same amount of the viral load than Gamma (Fig. S3b). For example, 1% of the samples (n = 4) classified as Gamma harbored approximately 96% of all viruses circulating in the state at the analyzed period. On the other hand, 1% of Delta samples corresponded to 42% of the viral load suggesting an elevated number of supercarriers within Delta lineage. Viral load of Delta tends to be high until the 7th day, while viral load from Gamma samples in our database seems to decrease from the 5th day (Fig. S4). The Delta variant is known for its higher transmissibility (32,33) and elevated viral loads, even when compared to the Alpha variant (25,34,35). Altogether, our results add the information that Delta also exhibits a remarkable enhancement in viral load values when compared to Gamma. This could have facilitated the rapid spread of Delta in the state of Rio de Janeiro (36) and sheds light on possible mechanisms underlying the successive lineage replacement of both variants.

We have identified at least nine independent introductions of Delta in Rio de Janeiro, but 98% of the genomic sequences from the state have originated from a single introductory event of the AY.99 lineage (File S1). Within the state, it has diverged into lineages AY.99.1 and AY.99.2 between May and June of 2021 (Fig. 3). Both lineages have spread across the state from this point onwards, with AY.99.2 being dominant. This dispersal has occurred in two stages in the state of Rio de Janeiro (Fig. 4). Until July, most transmissions were detected within the heavily populated city of Rio de Janeiro, with a small number of long-distance dispersals to the other macro regions of the state. From August onwards, these introductions in new cities/regions culminated in a high number of local and short-distance transmissions.

**Fig. 3.**
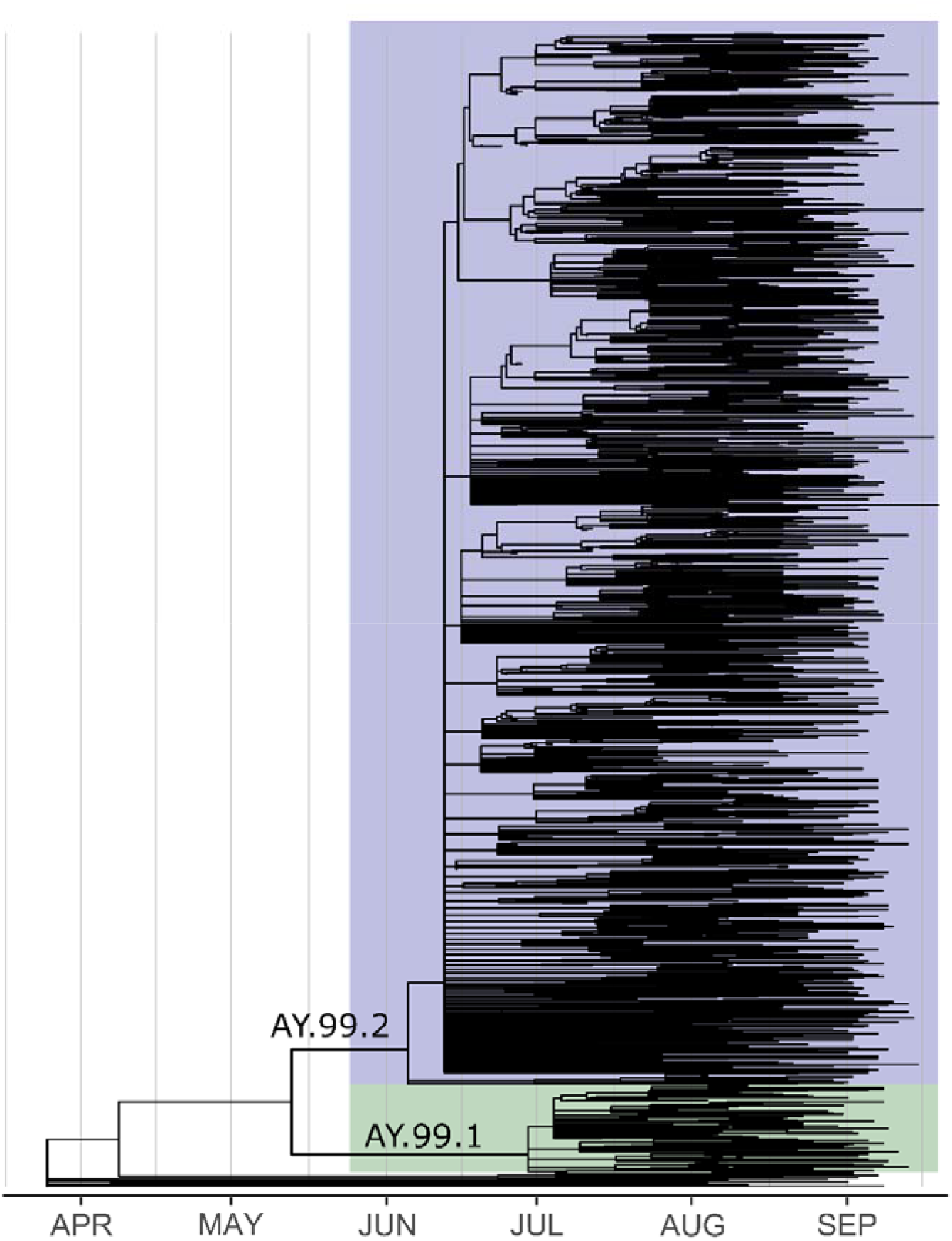
Time tree indicating the diversification of Delta into lineages AY.99.1 and AY.99.2 within the state of Rio de Janeiro. In highlight, the AY.99.1 and AY.99.2 clades.

**Fig. 4.**
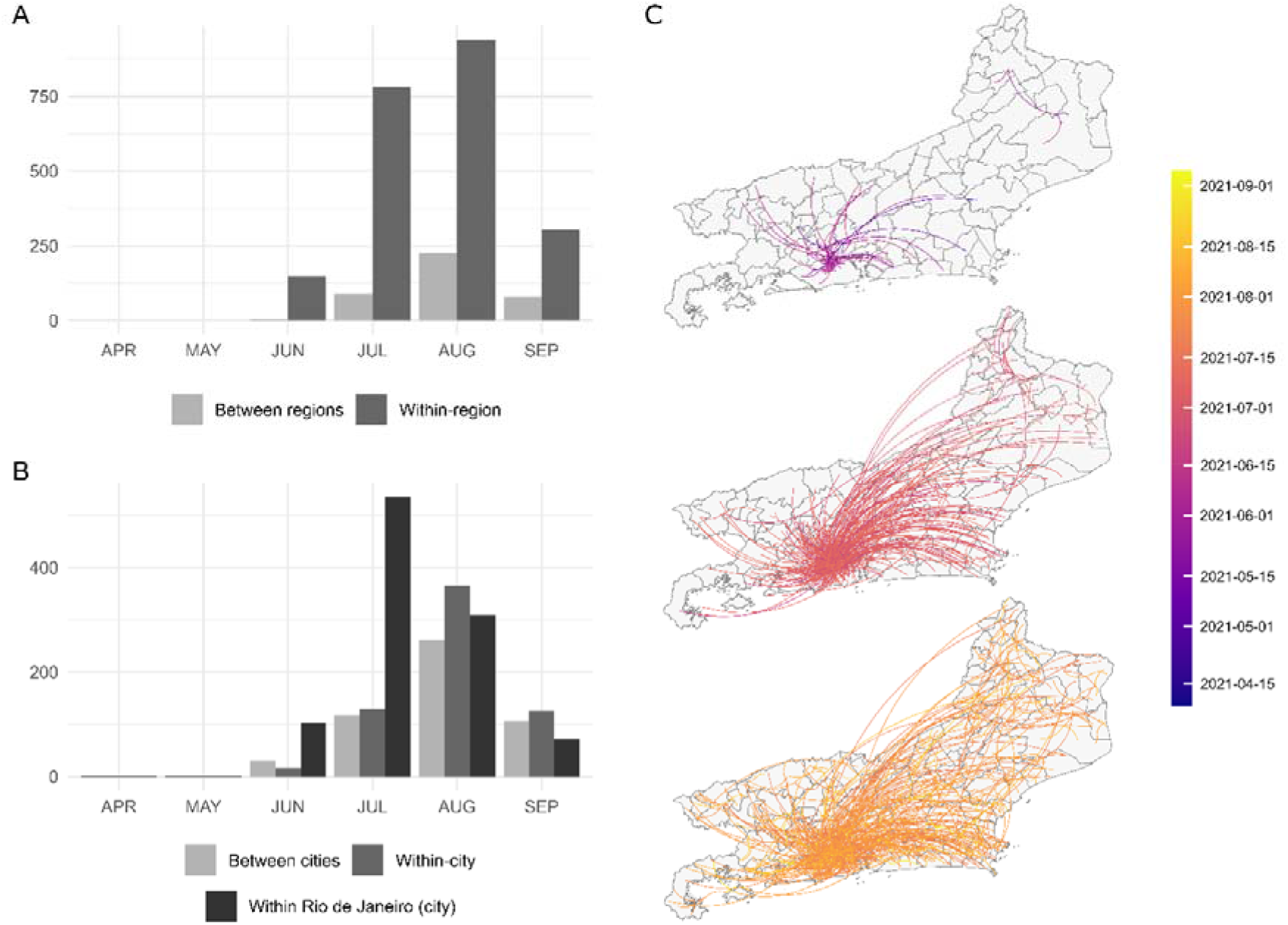
Spread of Delta variant across the state of Rio de Janeiro. Dispersal occurred more commonly within the macro regions of the state than between regions (A), demonstrating the importance of short- and medium-distance travels for terrestrial spread. When analyzing within-region transmissions (B), the city of Rio de Janeiro was the center of the spread in June and July. August onwards, this was surpassed by local transmission in the cities where the variant was introduced. In (C), the dispersal routes are indicated according to the inferred date of transmission.

These results consolidate the important role of large and dense urbanized areas as dispersal hubs immediately after introducing a new SARS-CoV-2 variant or lineage (6–13). Therefore, identification and surveillance of these hubs are of fundamental value to early control new variants that may emerge in the future. The results also suggest that non-medical interventions such as mass screening (37–40), use of masks (41–43), social distancing and lockdowns (44–46) in metropolitan areas might result in better long-term effects on pandemic control than when applied on small cities, because it may reduce the number of seeding events on small cities (47–49).

## Data Availability

All sequenced and assembled genomes are publicly available at GISAID (Table S1) and SRA-NCBI (BioProject PRJNA774631) databases

## Ethical approval

The study was approved by the Ethics Committee (30161620.0.1001.5257 and 34025020.0.0000.5257). Research protocol was approved without informed consent in accordance with Brazilian National Health Council’s Resolution 510/2016. All samples were residual COVID-19 clinical diagnostic samples de-identified before receipt by the researchers.

## Acknowledgements

We would like to thank all the authors and administrators of the GISAID database, which allowed this study of genomic epidemiology to be conducted properly. A complete list acknowledging the authors publishing data used in this study can be found in Table S2.

## Figures and tables

**Fig. S1.**
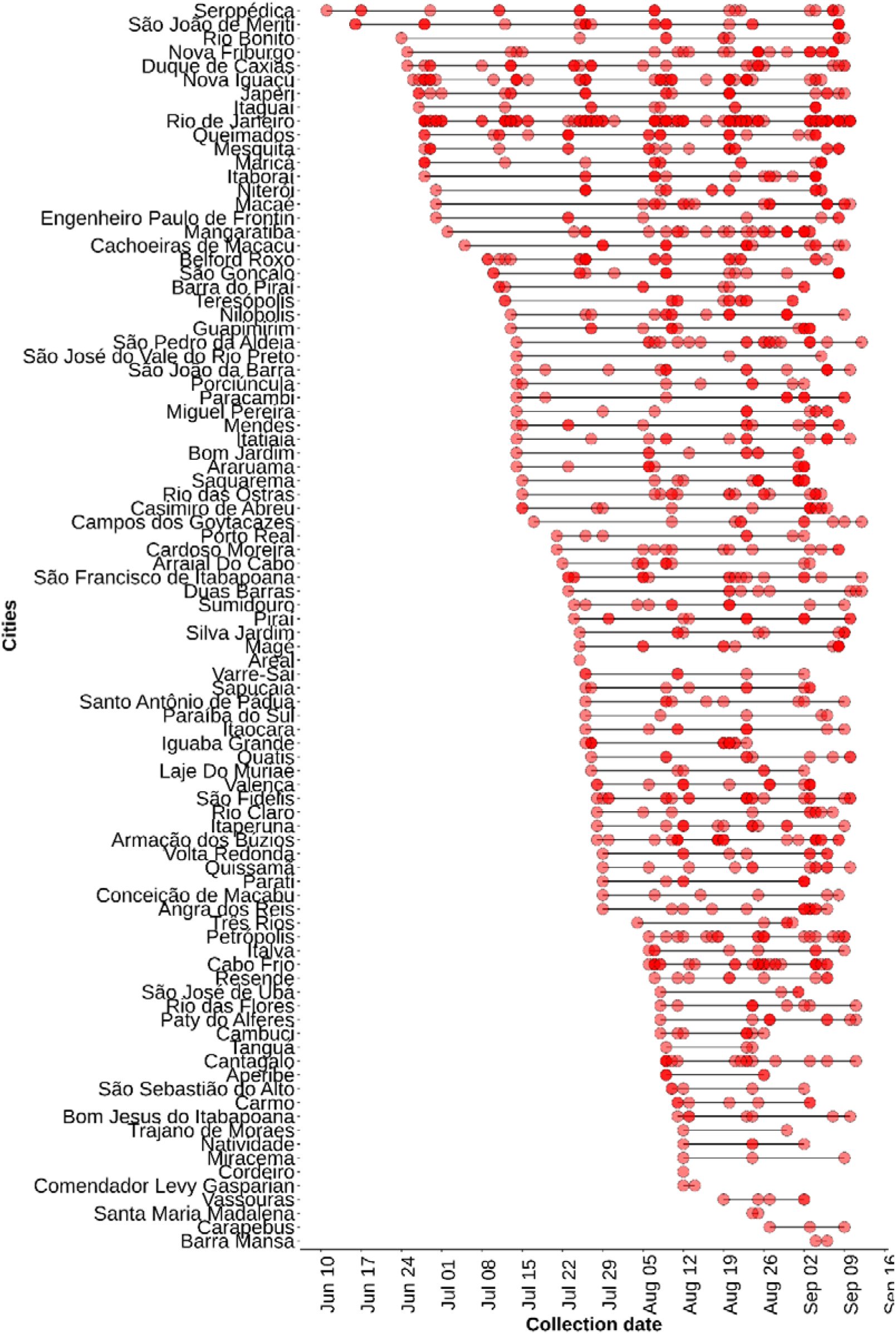
Sampling dates and localities of the Delta genomes sequenced in this work.

**Fig. S2.**
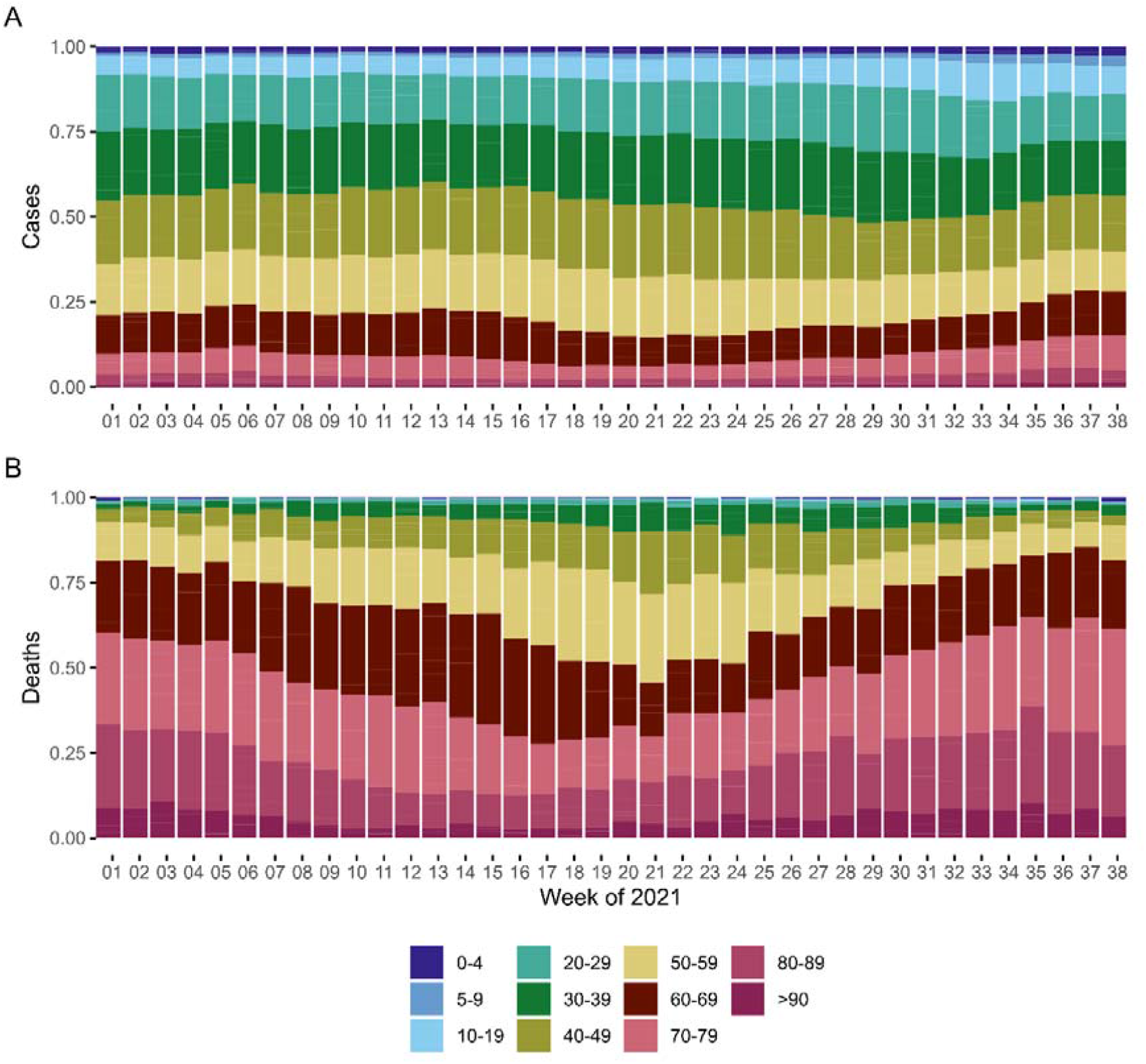
Age representation in total number of cases (A) and deaths (B) in the state of Rio de Janeiro during each week of 2021. Colors represent each age group analyzed.

**Fig. S3.**
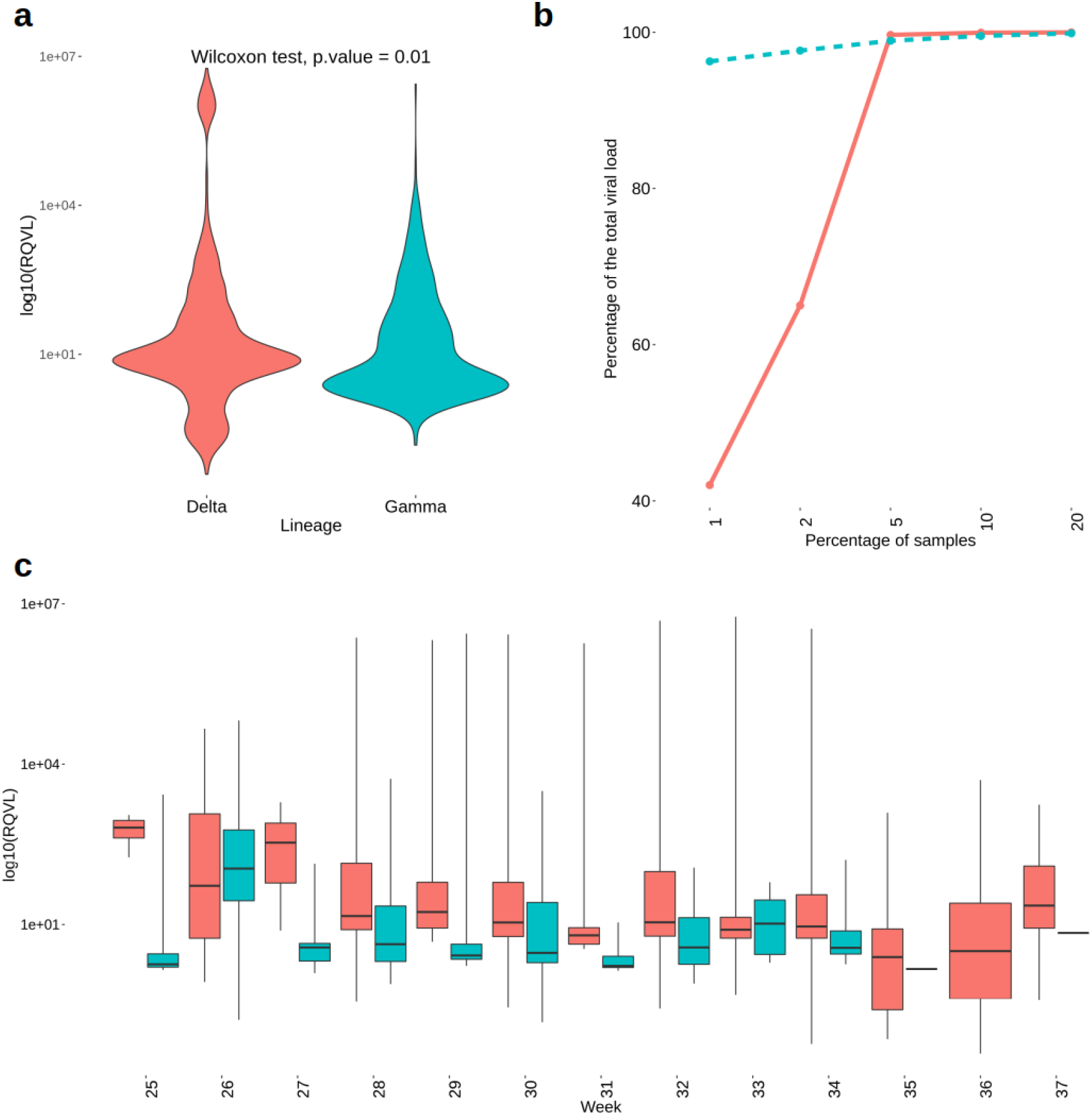
Viral load comparison between Delta and Gamma samples. **a)** Comparison between the RQVL values across Delta (red) and Gamma (blue). **b)** Cumulative percentage of total circulating virions from nasopharynx RQVL between Delta and Gamma. **c)** Distribution of RQVL in both VoCs over the period analyzed in this study.

**Fig. S4.**
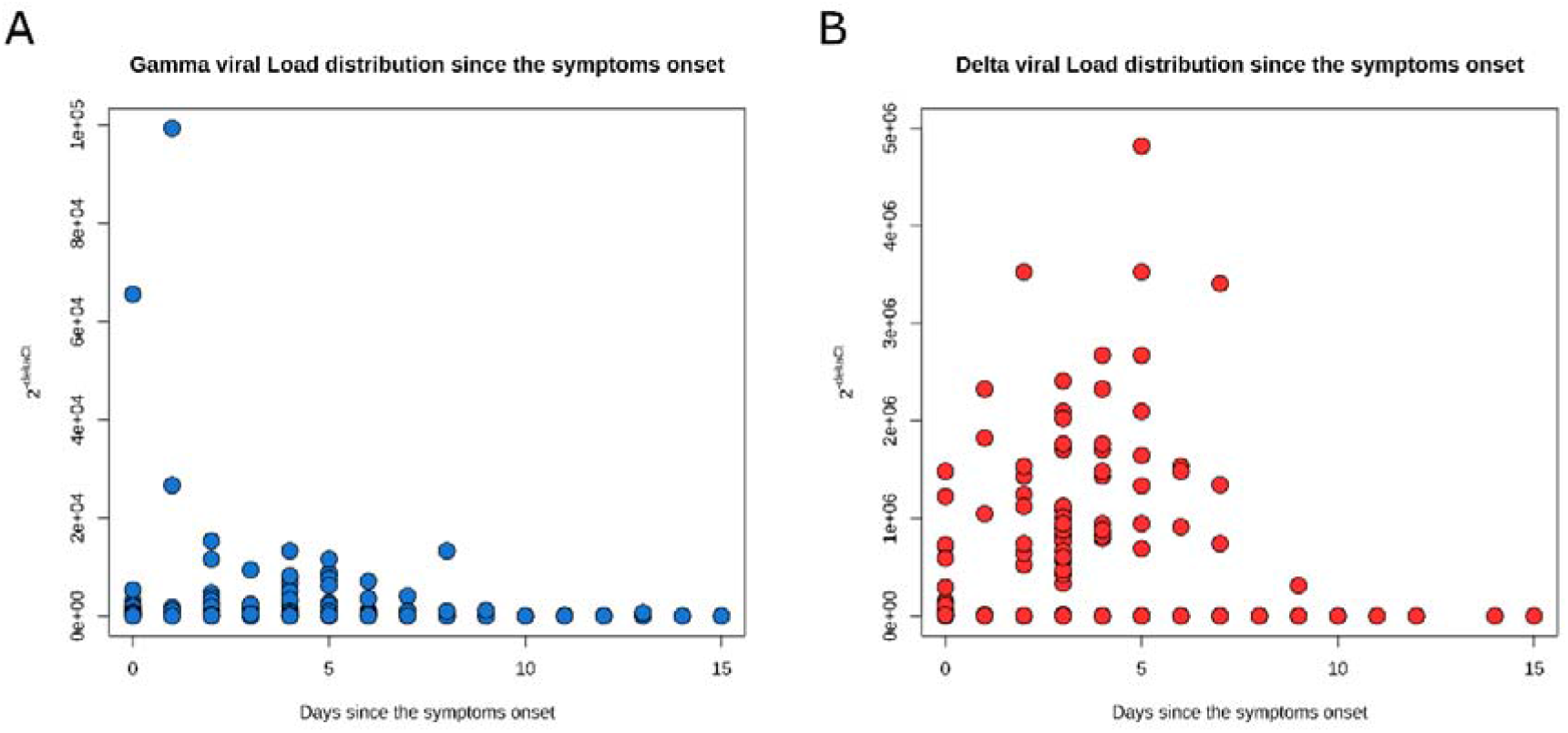
Viral load quantification over the days since the symptoms onset. A) Comparison between Gamma and B) Delta RQVL values and symptoms onset.

**Table S1. Accession number, sampling date and locality of the Delta genomes generated in this work**

**Table S2. Acknowledgement table from GISAID database**.

**File S1. Evolutionary tree in newick format generated using Delta genomes from Rio de Janeiro, Brazil, and around the world**.

