## Supplementary material for "Phylodynamic analysis of SARS-CoV-2 spread in Rio de Janeiro, Brazil, highlights how metropolitan areas act as dispersal hubs for new variants": Table S2

We gratefully acknowledge the following Authors from the Originating laboratories responsible for obtaining the specimens, as well as the Submitting laboratories where the genome data were generated and shared via GISAID, on which this research is based.

All Submitters of data may be contacted directly via www.gisaid.org

Authors are sorted alphabetically.

| Accession ID | Originating Laboratory | Submitting Laboratory | Authors |
|---|---|---|---|
| EPI_ISL_4539631 | "AR Dept. of Health-PHL, Molecular Diagnostics" | Centers for Disease Control and Prevention Division of Viral Diseases, Pathogen Discovery | Alex Burgin; Ben Rambo-Martin; Clinton Paden; Dakota Howard; Dave Wentworth; Dhwani Batra; Jasmine Padilla; Justin Lee; Krista Queen; Kristen Knipe; Kristine Lacek; Mark Burroughs; Matthew Schmerer; Meghan Bentz; Mili Sheth; Peter Cook; Sam Shepard; Sarah Nobles; Suxiang Tong; Vivien Dugan; Yvette Unoarumhi |
| EPI_ISL_3127938 | "CO Dept. of Public Health and Environment, Lab Services Division" | Centers for Disease Control and Prevention Division of Viral Diseases, Pathogen Discovery | Alex Burgin; Ben L. Rambo-Martin; Clinton R. Paden; Dakota Howard; Dave Wentworth; Dhwani Batra; Jasmine Padilla; Justin Lee; Krista Queen; Kristen Knipe; Kristine Lacek; Mark Burroughs; Matthew Schmerer; Meghan Bentz; Mili Sheth; Peter Cook; Sam Shepard; Sarah Nobles; Suxiang Tong; Vivien Dugan; Yvette Unoarumhi |
| EPI_ISL_3833906 | 4Cyte Pathology | NSW Health Pathology - Institute of Clinical Pathology and Medical Research; Westmead Hospital; University of Sydney | Arnott A.; Draper J.; Gall M.; Martinez E.; Rockett R.; Sintchenko V.; on behalf of ICPMR |
| EPI_ISL_4107883, EPI_ISL_4107943, EPI_ISL_4107957, EPI_ISL_4108092, EPI_ISL_4108241, EPI_ISL_4636632 | ACT Pathology | Schwessinger Lab | Ashley Jones; Benjamin Schwessinger; Carl McCombe; Carolina Correa Ospina; Craig Kennedy; Daniel Yu; Emma Crean; Karina Kennedy; Rene Riedelbauch; Robyn Hall |
| EPI_ISL_5020263, EPI_ISL_5336188 | AFIP | Instituto Butantan | Antonio Jorge Martins; Claudia Renata dos Santos Barros; David Schlesinger; Debora Botequio Moretti; Dimas Tadeu Covas; Elaine Cristina Marqueze; Elaine Vieira Santos; Evandra Strazza Rodrigues; Heidge Fukumasu; Jayme Augusto de Souza-Neto; José Salvatore Leister Patané; Luiz Alcantara; Luiz Lehmann Coutinho; Maria Carolina Elias; Maurício Lacerda Nogueira; Rafael dos Santos Bezerra; Raul Machado Neto; Rejane Maria Tommasini Grotto; Ricardo Haddad; Sandra Coccuzzo Sampaio Vessoni; Simone Kashima; Svetoslav Nanev Slavov; Vincent Louis Viala |
| EPI_ISL_3464546 | AREA DE SALUD ALAJUELA SUR | Inciensa, Instituto Costarricense de Investigación y Enseñanza en Nutrición y Salud | Adriana Godínez; Claudio Soto-Garita; Estela Cordero; Francisco Duarte; Hebleen Porras; Joselyn Prado & Juan Carlos Villalobos Ugalde; José Luis Vargas; Mariela Gutiérrez; Melany Calderón |
| EPI_ISL_5262733 | AREA DE SALUD CARIARI | Inciensa, Instituto Costarricense de Investigación y Enseñanza en Nutrición y Salud | Adriana Godínez; Claudio Soto-Garita; Estela Cordero; Francisco Duarte; Hebleen Porras; José Luis Vargas; Mariela Gutiérrez; Melany Calderón; Sofia Herrera & Leticia Gallegos Carrillo |
| EPI_ISL_3464520 | AREA DE SALUD COTO BRUS | Inciensa, Instituto Costarricense de Investigación y Enseñanza en Nutrición y Salud | Adriana Godínez; Claudio Soto-Garita; Estela Cordero; Francisco Duarte; Hebleen Porras; Joselyn Prado & Yendri Ramírez Alpízar; José Luis Vargas; Mariela Gutiérrez; Melany Calderón |
| EPI_ISL_3026022 | AREA DE SALUD EL GUARCO [EL GUARCO/CARTAGO] | Inciensa, Instituto Costarricense de Investigación y Enseñanza en Nutrición y Salud | Adriana Godínez; Caterina Guzmán; Claudio Soto-Garita; Estela Cordero; Francisco Duarte; Hebleen Porras; Joselyn Prado; José Luis Vargas; Mariela Gutiérrez; Melany Calderón; Nazareth Ruíz & Mónica Charpentier |
| EPI_ISL_5262735 | AREA DE SALUD FORTUNA | Inciensa, Instituto Costarricense de Investigación y Enseñanza en Nutrición y Salud | Adriana Godínez; Claudio Soto-Garita; Estela Cordero; Francisco Duarte; Hebleen Porras; José Luis Vargas; Mariela Gutiérrez; Melany Calderón; Sofia Herrera & Maria Fernanda Chacon Soto |
| EPI_ISL_3026027 | AREA DE SALUD FORTUNA [San Carlos/ALAJUELA] | Inciensa, Instituto Costarricense de Investigación y Enseñanza en Nutrición y Salud | Adriana Godínez; Caterina Guzmán; Claudio Soto-Garita; Estela Cordero; Francisco Duarte; Hebleen Porras; Joselyn Prado; José Luis Vargas; Mariela Gutiérrez; Melany Calderón; Nazareth Ruíz & Carolina Arrieta |
| EPI_ISL_5262712 | AREA DE SALUD GOICOECHEA 2 - CLINICA DR. JIMENEZ NUÑEZ | Inciensa, Instituto Costarricense de Investigación y Enseñanza en Nutrición y Salud | Adriana Godínez; Claudio Soto-Garita; Estela Cordero; Francisco Duarte; Hebleen Porras; José Luis Vargas; Mariela Gutiérrez; Melany Calderón; Sofia Herrera & Laura Marín López |
| EPI_ISL_5262729 | AREA DE SALUD HEREDIA-VIRILLA | Inciensa, Instituto Costarricense de Investigación y Enseñanza en Nutrición y Salud | Adriana Godínez; Claudio Soto-Garita; Estela Cordero; Francisco Duarte; Hebleen Porras; José Luis Vargas; Mariela Gutiérrez & Sofia Herrera; Melany Calderón |
| EPI_ISL_4601210 | AREA DE SALUD LIMON | Inciensa, Instituto Costarricense de Investigación y Enseñanza en Nutrición y Salud | Adriana Godínez; Claudio Soto-Garita; Estela Cordero; Francisco Duarte; Hebleen Porras; Joselyn Prado & Karolina Hall Loria; José Luis Vargas; Mariela Gutiérrez; Melany Calderón |
| EPI_ISL_4258642 | AREA DE SALUD OREAMUNO-PACAYAS-TIERRA BLANCA | Inciensa, Instituto Costarricense de Investigación y Enseñanza en Nutrición y Salud | Adriana Godínez; Claudio Soto-Garita; Estela Cordero; Francisco Duarte; Hebleen Porras; Joselyn Prado & Carolina Loria Acosta; José Luis Vargas; Mariela Gutiérrez; Melany Calderón |
| EPI_ISL_3638812 | AREA DE SALUD SAN JUAN-SAN DIEGO-CONCEPCION 2 | Inciensa, Instituto Costarricense de Investigación y Enseñanza en Nutrición y Salud | Adriana Godínez; Claudio Soto-Garita; Estela Cordero; Francisco Duarte; Hebleen Porras; José Luis Vargas; Mariela Gutiérrez & Joselyn Prado; Melany Calderón |
| EPI_ISL_5262758 | AREA DE SALUD SAN SEBASTIAN - PASO ANCHO | Inciensa, Instituto Costarricense de Investigación y Enseñanza en Nutrición y Salud | Adriana Godínez; Claudio Soto-Garita; Estela Cordero; Francisco Duarte; Hebleen Porras; José Luis Vargas; Mariela Gutiérrez & Sofia Herrera; Melany Calderón |
| EPI_ISL_3464539, EPI_ISL_5262716 | AREA DE SALUD SANTA CRUZ | Inciensa, Instituto Costarricense de Investigación y Enseñanza en Nutrición y Salud | Adriana Godínez; Claudio Soto-Garita; Estela Cordero; Francisco Duarte; Hebleen Porras; Joselyn Prado & Adriana Bermúdez Espinoza; José Luis Vargas; Mariela Gutiérrez; Melany Calderón; Sofia Herrera & Adriana Bermúdez Espinoza |
| EPI_ISL_3639134 | AREA DE SALUD TALAMANCA | Inciensa, Instituto Costarricense de Investigación y Enseñanza en Nutrición y Salud | Adriana Godínez; Claudio Soto-Garita; Estela Cordero; Francisco Duarte; Hebleen Porras; Joselyn Prado & Gloriana Barrantes; José Luis Vargas; Mariela Gutiérrez; Melany Calderón |
| EPI_ISL_3308832 | AULSS 2 Marca Trevigiana | Istituto Zooprofilattico Sperimentale delle Venezie | Adelaide Milani; Alessia Schivo; Alice Fusaro; Ambra Pastori; Annalisa Salviato; Antonia Ricci; Calogero Terregino; Edoardo Giussani; Elisa Palumbo; Erika Giorgia Quaranta; Isabella Monne; Luca Tassoni |
| EPI_ISL_5421069 | AUSTRAL-omics, UACh | AUSTRAL-omics, UACh | Andrea Silva; Carolina Encina; Cristian Molina; Daniela Plaza; Luis Guzmán; Suany Quesada |
| EPI_ISL_2833916 | Adan Hospital | Kuwait Cancer Control Center | Mona Alateeqi; Shakir Bahzad |
| EPI_ISL_5099123, EPI_ISL_5425682 | Addis Ababa University | CERI, Centre for Epidemic Response and Innvoation, Stellenbosch University and KRISP, KZN Research Innovation and Sequencing Platform, UKZN. | Abay Sisay; Abraham Tesfaye; Adey Feleke Desta; Giandhari Jennifer; Naidoo Yeshnee; Pillay Sureshnee; San James; Tegally Houriiyah; Tshiabuila Derek; Wilkinson Eduan; Yajna Ramphal; de Oliveira Tulio |
| EPI_ISL_4395235, EPI_ISL_4395250, EPI_ISL_4395252 | Addis Ababa University | KRISP, KZn Research Innovation and Sequencing Platform | Abay Sisay; Abraham Tesfaye; Adey Feleke Desta; Giandhari Jennifer; Naidoo Yeshnee; Pillay Sureshnee; San James; Tegally Houriiyah; Tshiabuila Derek; Wilkinson Eduan; Yajna Ramphal; de Oliveira Tulio |
| EPI_ISL_1690245, EPI_ISL_2875744, EPI_ISL_3430273, EPI_ISL_3845402, EPI_ISL_3846831, EPI_ISL_3849284, EPI_ISL_3853119, EPI_ISL_3855251, EPI_ISL_4376055, EPI_ISL_4382013, EPI_ISL_4495846, EPI_ISL_4554495, EPI_ISL_4981033, EPI_ISL_4986407, EPI_ISL_5111293 | | | |
| see above | Aegis Sciences Corporation | Centers for Disease Control and Prevention Division of Viral Diseases, Pathogen Discovery | Adrian Paskey; Alec Vest; Benjamin Rambo-Martin; Christopher Gulvick; Clinton Paden; Clinton R. Paden; Cyndi Clark; Dakota Howard; Darlene Wagner; Dhwani Batra; Dillon Nall; Duncan MacCannell; Erisa Sula; Ethan Sanders; Holly Houdeshell; Jason Caravas; Kara Moser; Kristine Lacek; Matthew Hardison; Matthew Schmerer; Ola Kvalvaag; Patrick Campbell; Peter Cook; Peter W. Cook; Rob Case; Scott Sammons; Shatavia Morrison; Shaun Westlund; Tymeckia Kendall; Victoria Caban Figueroa; Vikramsinha Ghorpade; Yvette Unoarumhi |
| EPI_ISL_4474366 | Africa_CDC - Angola | CERI, Centre for Epidemic Response and Innovation | Adriano Mendes; Amy Strydom; Emmanuel SJ; Giandhari J; Lessells R; Micheala Davids; Naidoo Y; Pillay S; Ramphal U; Sim Mayaphi and Marietjie Venter; Tegally H; Wilkinson E; de Oliveira T |
| EPI_ISL_3217412, EPI_ISL_3217415, EPI_ISL_3217424 | Airport Health Laboratory/Central Health Laboratory | Virology Department, Central Health Laboratory ,Victoria Hospital, Candos,Ministry of Health and Wellness, Mauritius | Bahadoor BS; Jannoo N; Manraj SS; Mathur H; Pattoo M; Ramuth M; Sonoo J; Sujeewon C |
| EPI_ISL_4739718 | Akershus University Hospital, Department for Microbiology and Infectious Disease Control | Norwegian Institute of Public Health, Department of Virology | Atiya R Ali; Debech Nadia; Engebretsen Serina Beate; Garcia Llorente Ignacio; Hilde Elshaug; Hilde Vollan; Jon Bråte; Kamilla Heddeland Instefjord; Karoline Bragstad; Kathrine Stene-Johansen; Line Victoria Moen; Marie Paulsen Madsen; Olav Hungnes; Pedersen Benedikte Nevjen; Rasmus Riis Kopperud |

| | | | |
|---|---|---|---|
| EPI_ISL_2966437, EPI_ISL_4846958 | Alaska State Virology Laboratory | Alaska State Virology Laboratory | Elva House; Jack Chen; Jacob Zidek; Lisa Smith; Ph.D.; Stephanie DeRonde |
| EPI_ISL_3720888, EPI_ISL_3720917, EPI_ISL_5032187, EPI_ISL_5032234 | Allergy, Immunology and Cell Biology Unit (AICBU) | Allergy, Immunology and Cell Biology Unit (AICBU) | Ayesha Wijesinghe; Chandima Jeewandara; Deshni Jayathilaka; Dinuka Ariyarathne; Diyanath Ranasinghe; Dumni Gunasinghe; Gathsaurie Neelika Malavige; Tibutius Thanesh |
| EPI_ISL_2633506, EPI_ISL_4254570 | Area of Virology, Serology and Virology Division (SAViD), New South Wales Health Pathology Randwick | Virology Research Laboratory; Area of Virology, Serology and Virology Division (SAViD), New South Wales Health Pathology Randwick | Au, J.; Bull, R.; Deveson, I.; Foster, C.; Rawlinson, W.; Ruiz Silva, M.; Van Hal, S. |
| EPI_ISL_3833928 | Austech Medical Laboratories | NSW Health Pathology - Institute of Clinical Pathology and Medical Research; Westmead Hospital; University of Sydney | Arnott A.; Draper J.; Gall M.; Martinez E.; Rockett R.; Sintchenko V.; on behalf of ICPMR |
| EPI_ISL_4552195, EPI_ISL_4556568, EPI_ISL_5305953 | Australian Clinical Labs (formerly Healthscope Pathology) | NSW Health Pathology - Institute of Clinical Pathology and Medical Research; Westmead Hospital; University of Sydney | Arnott A.; Draper J.; Gall M.; Martinez E.; Rockett R.; Sintchenko V.; on behalf of ICPMR |
| EPI_ISL_3231044, EPI_ISL_3307648, EPI_ISL_3546434, EPI_ISL_4060571, EPI_ISL_5031321, EPI_ISL_5063296 | Austrian Agency for Health and Food Safety (AGES) | Bergthaler laboratory, CeMM Research Center for Molecular Medicine of the Austrian Academy of Sciences | Andreas Bergthaler; Anna Schedl; Bekir Erguner; Benedikt Agerer; Christoph Bock; Fabian Amman; Jan Laine; Lukas Endler; Maelle Le Moing; Martin Senekowitsch; Matthew Thornton; Michael Schuster; Petr Triska; Thomas Penz |
| EPI_ISL_5020466 | BIOFAST | Instituto Butantan | Antonio Jorge Martins; Claudia Renata dos Santos Barros; David Schlesinger; Debora Botequio Moretti; Dimas Tadeu Covas; Elaine Cristina Marqueze; Elaine Vieira Santos; Evandra Strazza Rodrigues; Heidge Fukumasu; Jayme Augusto de Souza-Neto; José Salvatore Leister Patané; Luiz Alcantara; Luiz Lehmann Coutinho; Maria Carolina Elias; Maurício Lacerda Nogueira; Rafael dos Santos Bezerra; Raul Machado Neto; Rejane Maria Tommasini Grotto; Ricardo Haddad; Sandra Coccuzzo Sampaio Vessoni; Simone Kashima; Svetoslav Nanev Slavov; Vincent Louis Viala |
| EPI_ISL_2373545 | Banaras Hindu University | CSIR-Centre for Cellular and Molecular Biology-INSACOG | Amareshwar Vodapalli; Ara Sreenivas; Archana Bharadwaj Siva; Divya Tej Sowpati; Gunjan Rai; Gyaneshwer Chaubey; Karthik Bharadwaj Tallapaka; Lamuk Zaveri; Maneesha Upadhyay; Payel Mukherjee; Rakesh K Mishra; Royana Singh; Sharath Chandra Thota; Shivam Tiwari; Shivani Mishra; Shreekant Verma; Surendra Pratap Mishra; Tulasi Nagabandi; Umesh Choudhary; Valli Nagalakshmi Undamatla |
| EPI_ISL_3274186 | Baptist Health Medical Center | Center for Global Health, University of New Mexico Health Sciences Center | Amanda Novack; Darrell Dinwiddie; Daryl Domman; Dirk Haselow; Joshua L. Kennedy; Kurt Schwalm; Valerie Morley |
| EPI_ISL_5055870, EPI_ISL_5332116 | Białostockie Centrum Analiz Medycznych | 1. Academic Center for Pathomorphological and Genetic-Molecular Diagnostics ltd, Bialystok, Poland 2. National Institute of Public Health - National Institute of Hygiene, Warsaw, Poland | Anetta Sulewska; Jacek Nikliński; Janusz Dzięcioł; Joanna Kiśluk; Katarzyna Zacharczuk; Konrad Raczkowski; Magdalena Nowakowska; Małgorzata Sadkowska-Todys; Piotr Karabowicz; Piotr Majewski; Przemysław Biecek. Joanna Reszeć; Radosław Charkiewicz; Tomasz Wołkowicz |
| EPI_ISL_3146408, EPI_ISL_3146499, EPI_ISL_3334363, EPI_ISL_5391795, EPI_ISL_5391796, EPI_ISL_5392385, EPI_ISL_5392673, EPI_ISL_5392702 | | | |
| see above | BioneXt Lab | Laboratoire national de sante, Microbiology, Microbial Genomics Platform | Anke Wienecke-Baldacchino; Catherine Ragimbeau; Elodie Solarino; Fatu Djabi; Jessica Tapp; Lise Pignon; Raoul Salmon; Tamir Abdelrahman; Thibault Ferrandon; Virginie Jover |
| EPI_ISL_3390947 | Biopole Antilles | Department of Virology, Henri Mondor University Hospital, Assistance Publique Hôpitaux de Paris, Université Paris-Est Créteil, INSERM U955 | Alexandre Soulier; Christophe Rodriguez; Elisabeth Trawinski; Guillaume Gricourt; Jean-Michel Pawlotsky; Melissa N'Debi; Slim Fourati; Vanessa Demontant |
| EPI_ISL_4489990, EPI_ISL_4490091 | Biorepository and Clinical Virology Lab, UCH | Africa Centre for Excellence for Genomics of Infectious Diseases (ACEGID), Redeemer's University | A.T.; Abechi; Ajogbasile; Akano; C.A.; C.T.; Eromon; F.V.; Folarin, O.; Happi; I.B.; J.N.; J.U.; K.O.; Kayode; Nosamiefan, I.; Oguzie; Olawoye; Olumade; Oluniyi; P.E.; P.S.; T.J.; Ugwu; Uwanibe |
| EPI_ISL_3495645, EPI_ISL_4438092 | Bioscientia Labor Wermsdorf | Robert Koch Institute | |
| EPI_ISL_3948408, EPI_ISL_4299843, EPI_ISL_4299844 | Botswana Harvard AIDS Institute Partnership | Botswana Harvard HIV Reference Laboratory | Boitumelo Zuze; Botshelo Radibe; Dorcas Maruapula; Joseph Makhema; Keoratile Ntshambiwa; Kgomotso Moruisi; Legodile Kooepile; Mosepele Mosepele; Mphaphi B. Mbulawa; Ontlametse T. Bareng; Pamela Smith-Lawrence; Patrick T. Mokgethi; Roger Shapiro; Sefetogi Ramaologa; Shahin Lockman; Sikhulile Moyo; Simani Gaseitsiwe; Thongbotho Mphoyakgosi; Wonderful T. Choga |
| EPI_ISL_2820343, EPI_ISL_2820357, EPI_ISL_2820445, EPI_ISL_2868373 | Botswana Harvard HIV Reference Laboratory | Botswana Harvard HIV Reference Laboratory | Boitumelo Zuze; Botshelo Radibe; Dorcas Maruapula; Godfrey Simoonga; Joseph Makhema; Keoratile Ntshambiwa; Kereng Mphoyakgosi; Legodile Kooepile; Madisa Mine; Modisa Motswaledi; Mosepele Mosepele; Motlalepule L. Pone; Ontlametse T. Bareng; Roger Shapiro; Shahin Lockman; Sikhulile Moyo; Simani Gaseitsiwe; Thongbotho Mphoyakgosi; Wonderful T. Choga |
| EPI_ISL_3602791, EPI_ISL_5222716 | British Columbia Centre For Disease Control | BCCDC Public Health Laboratory | Ana Pacagnella; Corrinne Ng; Dan Fornika; John Tyson; Kim Macdonald; Kimia Kamelian; Linda Hoang; Loretta Janz; Mel Krajden; Prystajecky Natalie; Robert Azana; Shannon Russell |
| EPI_ISL_3007334, EPI_ISL_3007481, EPI_ISL_3407572, EPI_ISL_3669393, EPI_ISL_3669403, EPI_ISL_3839016, EPI_ISL_5009817, EPI_ISL_5252252 | | | |
| see above | Broad Institute Clinical Research Sequencing Platform | Infectious Disease Program, Broad Institute of Harvard and MIT | Adams, G.; B.L.; B.W.; Bauer, M.; Birren; Blumenstiel, B.; Brown, C.; Carter, A.; Chaluvadi, S.; D.J.; DeFelice, M.; DeRuff, K.; Dodge, S.; Gabriel, S.; Gallagher, G.; Gladden-Young, A.; Granger, B.; J.E.; K.J.; Lagerborg, K.; Larkin, K.; Lee, M.; Lemieux; Lennon, N.; Loreth, C.; Madoff, L.; McGovern, S.; Meldrim, J.; Normandin, E.; P.C.; Park; Pearlman, L.; Reilly, S.; Rudy, M.; Sabeti; Siddle; Smole, S.; Tomkins-Tinch, C.; Vicente, G.; and MacInnis |
| EPI_ISL_4607032 | CERBAILLIANCE LA REUNION | CNR Virus des Infections Respiratoires - France SUD | Antonin Bal; Bruno Lina; Gregory Destras; Gwendolyne Burfin; Hadrien Regue; Laurence Josset; Martine Valette; Quentin Semanas |
| EPI_ISL_2895286, EPI_ISL_5304517 | CHTMAD | Instituto Nacional de Saude (INSA) | Borges et al |
| EPI_ISL_2989032, EPI_ISL_3307080 | CHU Sao Joao, Porto | Instituto Nacional de Saude (INSA) | Borges et al |
| EPI_ISL_3948487 | CLINICA BIBLICA | Inciensa, Instituto Costarricense de Investigación y Enseñanza en Nutrición y Salud | Adriana Godínez; Claudio Soto-Garita; Estela Cordero; Francisco Duarte; Hebleen Porras; José Luis Vargas; Mariela Gutiérrez & Joselyn Prado; Melany Calderón |
| EPI_ISL_5313732 | CNR Virus des Infections Respiratoires - France SUD | CNR Virus des Infections Respiratoires - France SUD | Antonin Bal; Bruno Lina; Gregory Destras; Gwendolyne Burfin; Hadrien Regue; Laurence Josset; Martine Valette; Quentin Semanas |
| EPI_ISL_2373426 | CSIR-Centre for Cellular and Molecular Biology | CSIR-Centre for Cellular and Molecular Biology-INSACOG | Amareshwar Vodapalli; Ara Sreenivas; Archana Bharadwaj Siva; B Himasri; Blessy B John; Divya Tej Sowpati; Karthik Bharadwaj Tallapaka; Lamuk Zaveri; Payel Mukherjee; Rakesh K Mishra; Sharath Chandra Thota; Shreekant Verma; Sofia Banu; Tulasi Nagabandi; Valli Nagalakshmi Undamatla; Viswagithe S L |
| EPI_ISL_4917645 | Cantacuzino National Military-Medical Institute, Viral Respiratory Infections Laboratory | Cantacuzino Institute Virology | Carmen Cherciu; Luiza Ustea; Mihaela Lazar; Mihaela Oprea; Nicoleta Paraschiv; Sorin Dinu |
| EPI_ISL_5241315, EPI_ISL_5393384, EPI_ISL_5393426, EPI_ISL_5393443 | Cantacuzino National Military-Medical Institute, Viral Respiratory Infections Laboratory | Cantacuzino National Military-Medical Institute, Viral Respiratory Infections Laboratory | Carmen Cherciu; Luiza Ustea; Mihaela Lazar; Mihaela Oprea; Nicoleta Paraschiv; Sorin Dinu |
| EPI_ISL_2811947, EPI_ISL_2811958, EPI_ISL_3477088, EPI_ISL_3477089, EPI_ISL_3477094, EPI_ISL_3543460, EPI_ISL_3869457, EPI_ISL_4571452, EPI_ISL_5348878 | | | |
| see above | Canterbury Health Laboratories | Institute of Environmental Science and Research (ESR) | Anja Werno; Antje van der Linden; Arlo Upton; Chris Mansell; Clare Gebbie; David Hammer; Dhanisha Patel; Dragana Drinkovic; Erasmus Smit; Gary McAuliffe; Hana Sofia Andersson; Hermes Perez; James Ussher; Jill Sherwood; Jing Wang; Joep de Ligt; Josh Freeman; Julia Howard; Juliet Elvy; Lauren Jelly; Mary DeAlmeida; Matt Blakiston; Matt Storey; Matthew Rogers; Max Bloomfield; Michael Addidle; Michelle Balm; Muhammad Faisal; Nikki Freed; Olin Silander; Olivia Stroeven; Paula scholes; Rachel Boyle; Sally Roberts; SallyAnn Harbison; Sarah Cockerton; Sarah Jefferies; Sharmini Muttaiyah; Susan Lin; Susan Morpeth; Susan Taylor; Timothy Blackmore; Vani Sathyendran; Veronica Playle; Virginia Hope; Xiaoyun Ren |
| EPI_ISL_4108122, EPI_ISL_4263241, EPI_ISL_4731639 | Capital Pathology | Schwessinger Lab | Ashley Jones; Benjamin Schwessinger; Carl McCombe; Carolina Correa Ospina; Daniel Yu; Emma Crean; Karina Kennedy; Paul Whiting; Rene Riedelbauch; Robyn Hall; Sandra Molloy |
| EPI_ISL_4003130, EPI_ISL_5017929, EPI_ISL_5017952, EPI_ISL_5017955, EPI_ISL_5017964, EPI_ISL_5017968, EPI_ISL_5017983, EPI_ISL_5017986, EPI_ISL_5017989, EPI_ISL_5018011 | | | |
| see above | Central Laboratory, Bureau of Public Health (BOG) and Academic Hospital Paramaribo | Erasmus Medical Center | Bas B Oude Munnink; Cherise Beek; Consuella Partowidjojo; Dion Gajadin; Ed PF IJzerman; Emmanuelle Munger; Gary Gummels; Ingrid SK Krishnadath; Lycke Woittiez; Marion PG Koopmans; Mireille Van de Veer; Phyllis Pinas; Princes Wongsowidjojo; Radjesh Ori; Ranisha Doerbalie; Rohma Banwari; Soeradj Harkisoen; Stephen Vreden; Tilotmadebie Ramlal; Verne Nanhoe |
| EPI_ISL_4296407 | Central Medical Laboratory | Baylor College of Medicine | Adrianna Maliga; Alexander Kneubehl; Allison Lino; Gerhaldine Morazan; Kristy Murray; Russell Manzanero; Sarah Gunter; Sarah Strobel; Shannon Ronca |
| EPI_ISL_4948709, EPI_ISL_4948749, EPI_ISL_4948762, EPI_ISL_4948780, EPI_ISL_4948781, EPI_ISL_4948904, EPI_ISL_4948916, EPI_ISL_4948927, EPI_ISL_4948965, EPI_ISL_4948995, EPI_ISL_4949012 | | | |

| | | | |
|---|---|---|---|
| see above | Central Public Health Laboratory | Microbiological Diagnostic Unit - Public Health Laboratory (MDU-PHL) | Horan, K.; Kumbu, J.; N.L.; Palou, T.; Seemann, T.; Sherry; Susapu, M. |
| EPI_ISL_4462214, EPI_ISL_4462240 | Central Public Health Laboratory, National Public Health Organization | Central Public Health Laboratory, National Public Health Organization | Antigoni Katsoulidou; Gregory Spanakos; Kyriaki Tryfinopoulou; Olga Papa et al |
| EPI_ISL_4548657 | Centrala Laboratorija | Latvian Biomedical Research and Study Centre | Davids Fridmanis; Elina Dimina; Guntars Zarins; Ivars Silamikelis; Jana Osite; Janis Klovins; Janis Pjalkovskis; Jurijs Perevoscikovs; Kaspars Megnis; Laila Silamikele; Lauma Freimane; Laura Ansone; Liga Birzniece; Marta Priedite; Monta Briviba; Nikita Zrelovs; Uga Dumpis; Una Krumina; Vita Rovite |
| EPI_ISL_5394127 | Centre Hospitalier du Nord | Laboratoire national de sante, Microbiology, Microbial Genomics Platform | Anke Wienecke-Baldacchino; Catherine Ragimbeau; Elodie Solarino; Fatiha Boulmerka; Fatu Djabi; Jacqueline Parmentier; Jessica Tapp; Lise Pignon; Raoul Salmon; Tamir Abdelrahman; Virginie Jover |
| EPI_ISL_4704012, EPI_ISL_4704580, EPI_ISL_4708211, EPI_ISL_5031200 | Centre de Recherches Médicales de Lambaréné | Centre de Recherches Médicales de Lambaréné | Ayong More; Bertrand Lell; Gédéon Prince Manouana; Jean Bernard Lekana-Douki; Joël-Fleury Djoba Siawaya and Ayola Akim Adegnika; Moustapha Nzamba Maloum; Rodrigue Bikangui; Roméo Aimé Laclong Lontchi; Sam O'neilla Oye Bingono; Samira Zoa Assoumou; Srinivas reddy Pallerla; Steffen Borrmann |
| EPI_ISL_1970418 | Centre for Dengue Research and AICBU, Department of Immunology and Molecular Medicine | Centre for Dengue Research and AICBU, Department of Immunology and Molecular Medicine | Chandima Jeewandara; Deshan Madhusanka; Deshni Jayathilaka; Dinuka Ariyaratne; Diyanath Ranasinghe; Gathsaurie Neelika Malavige; Laksiri Gomes |
| EPI_ISL_5063558 | Centre of Excellence in Molecular Biology | Quadram Institute Bioscience | Aamir Ashiq; Alexander J Trotter; Alison E. Mather; Almina Shafiq; Alp Aydin; Ana P. Tedim; Anastasia Kolyva; Andrew Bell; Andrew J. Page; Asghar javeed; Claire Stuart; Dave J. Baker; Gemma L. Kay; Hazir Rahman; Iqra Jamil; Javed Akram; John Wain; Justin O'Grady; Khushbakhth kirn; Leonardo de Oliveira Martins; Lizzie Meadows; Maria Diaz; Mark Webber; Martin Lott; Michaela Matthews; Muhammad Asif Naveed; Muhammad Bilal Sarwar; Muhammad Roman; Muhammed Yasir; Nabil-Fareed Alikhan; Nadeem Afzal; Ngozi Elumogo; Nicholas M. Thomson; Rachael Stanley; Rachel Gilroy; Reenesh Prakash; Samir Dervisevic; Samuel Bloomfield; Shah Jahan; Sidra-tul-muntaha; Steven Rudder; Thanh Le-Viet; Waseem Iqbal |
| EPI_ISL_3805347, EPI_ISL_4549959 | Centro de Investigacion Biomedica de Occidente (CIBO) | Unidad de Genomica Avanzada | ; Alejandra Garcia-Gasca; Alejandra Hernandez-Teran; Alejandro Sanchez-Flores; Alfredo Herrera-Estrella; Alicia Ocaña-Mondragon; Andreu Comas-Garcia; Angel Gustavo Salas-Lais; Antonio Loza Roman; Bernardo Martinez-Miguel; Blanca Taboada; Brenda Irasema Maldonado-Meza; Bruno Gomez-Gil; Carla Ivon Herrera-Najera; Carlos F. Arias; Celia Boukadida; Celida Duque Molina; Celida Martinez- Rodriguez; Clara Esperanza Santacruz-Tinoco; Concepcion Grajales-Muñiz; Consorcio Mexicano de Vigilancia Genomica (CoViGen-Mex). Authors (in alphabetical order): Julio Elias Alvarado-Yaah; Cristobal Chaidez-Quiroz; Daniel Fregoso-Rueda; Daniel Lira Morales; Eduardo Becerril-Vargas; Fernando Fontove-Herrera; Fidencio Mejia-Nepomuceno; Francisco Pulido; Gloria Elena Espinosa-Ayala; Gloria Maria Molina-Salinas; Gloria Vazquez; Hector Esteban Paz-Juarez; Hector Montoya-Fuentes; Helen Haydee Fernanda Ramirez-Plascencia; Irvin Gonzalez-Lopez; Jean Pierre Gonzalez; Jesus Hernandez; Joel Armando Vazquez-Perez.; Jorge Salas-Hernandez; Jose Antonio Enciso-Moreno; Jose Arturo Martinez-Orozco; Jose Esteban Muñoz-Medina; Jose de Jesus Nuñez-Contreras; Juan Bautista Chale-Dzul; Julissa Enciso-Ibarra; Luis Alberto Ochoa-Carrera; Margarita Matias-Florentino; Maria Guadalupe Santiago-Mauricio; Maria Guadalupe de Jesus Mireles-Rivera; Mario Mujica-Sanchez; Marissa Perez-Garcia; Nelly Selem-Mojica; Pavel Isa; Ricardo Ciria Merce; Ricardo Grande; Rosa Maria Gutierrez Rios; Santiago avila-Rios; Selene Zarate; Susana Lopez; Veronica Mata-Haro; Victor Eduardo Garcia-Arias; Victor Hugo Borja-Aburto |
| EPI_ISL_2942512, EPI_ISL_2942776 | Centro de Investigacion Biomedica del Noreste (CIBIN) | Unidad de Genomica Avanzada | ; Alejandra Garcia-Gasca; Alejandra Hernandez-Teran; Alejandro Sanchez-Flores; Alfredo Herrera-Estrella; Alicia Ocaña-Mondragon; Andreu Comas-Garcia; Angel Gustavo Salas-Lais; Antonio Loza Roman; Bernardo Martinez-Miguel; Blanca Taboada; Brenda Irasema Maldonado-Meza; Bruno Gomez-Gil; Carla Ivon Herrera-Najera; Carlos F. Arias; Celia Boukadida; Celida Duque Molina; Celida Martinez- Rodriguez; Clara Esperanza Santacruz-Tinoco; Concepcion Grajales-Muñiz; Consorcio Mexicano de Vigilancia Genomica (CoViGen-Mex). Authors (in alphabetical order): Julio Elias Alvarado-Yaah; Cristobal Chaidez-Quiroz; Daniel Fregoso-Rueda; Daniel Lira Morales; Eduardo Becerril-Vargas; Fernando Fontove-Herrera; Fidencio Mejia-Nepomuceno; Francisco Pulido; Gloria Elena Espinosa-Ayala; Gloria Maria Molina-Salinas; Gloria Vazquez; Hector Esteban Paz-Juarez; Hector Montoya-Fuentes; Helen Haydee Fernanda Ramirez-Plascencia; Irvin Gonzalez-Lopez; Jean Pierre Gonzalez; Jesus Hernandez; Joel Armando Vazquez-Perez.; Jorge Salas-Hernandez; Jose Antonio Enciso-Moreno; Jose Arturo Martinez-Orozco; Jose Esteban Muñoz-Medina; Jose de Jesus Nuñez-Contreras; Juan Bautista Chale-Dzul; Julissa Enciso-Ibarra; Luis Alberto Ochoa-Carrera; Margarita Matias-Florentino; Maria Guadalupe Santiago-Mauricio; Maria Guadalupe de Jesus Mireles-Rivera; Mario Mujica-Sanchez; Marissa Perez-Garcia; Nelly Selem-Mojica; Pavel Isa; Ricardo Ciria Merce; Ricardo Grande; Rosa Maria Gutierrez Rios; Santiago avila-Rios; Selene Zarate; Susana Lopez; Veronica Mata-Haro; Victor Eduardo Garcia-Arias; Victor Hugo Borja-Aburto |
| EPI_ISL_4299729, EPI_ISL_5194145, EPI_ISL_5194178 | Centro de Investigación Biomédica de Occidente (CIBO) | Centro de Investigación en Enfermedades Infecciosas (CIENI), Instituto Nacional de Enfermedades Respiratorias (INER) | Alejandra García-Gasca; Alejandra Hernández-Terán; Alejandro Sánchez-Flores; Alfredo Herrera-Estrella; Alicia Ocaña-Mondragón; Andreu Comas-García; Angel Gustavo Salas-Lais; Antonio Loza Román; Bernardo Martínez-Miguel; Blanca Taboada; Brenda Irasema Maldonado-Meza; Bruno Gómez-Gil; Carla Ivón Herrera-Najera; Carlos F. Arias; Celia Boukadida; Clara Esperanza Santacruz-Tinoco; Concepción Grajales-Muñiz; Consorcio Mexicano de Vigilancia Genómica (CoViGen-Mex). Authors (in alphabetical order): Julio Elias Alvarado-Yaah; Cristóbal Cháidez-Quiróz; Célida Duque Molina; Célida Martínez-Rodríguez; Daniel Fregoso-Rueda; Daniel Lira Morales; Eduardo Becerril-Vargas; Eduardo Rivera-Martínez; Fernando Fontove-Herrera; Fidencio Mejía-Nepomuceno; Francisco Pulido; Gabriel Chavira-Trujillo; Gloria Elena Espinosa-Ayala; Gloria María Molina-Salinas; Gloria Vazquez; Hector Montoya-Fuentes; Helen Haydee Fernanda Ramírez-Plascencia; Irvin González-López; Jean Pierre González; Jesús Hernández; Joel Armando Vázquez-Pérez.; Jorge Salas-Hernández; José Antonio Enciso-Moreno; José Arturo Martínez-Orozco; José Esteban Muñoz-Medina; José de Jesús Nuñez-Contreras; Juan Bautista Chale-Dzul; Julissa Enciso-Ibarra; Kathia Elizabeth Tapia-Diaz; Luis Alberto Ochoa-Carrera; Margarita Matías-Florentino; Mario Mújica-Sánchez; Marissa Perez-Garcia; María Eugenia Jiménez-Corona; María Guadalupe Santiago-Mauricio; María Guadalupe de Jesús Mireles-Rivera; Nelly Sélem-Mojica; Pavel Isa; Ricardo Ciria Merce; Ricardo Grande; Rosa María Gutiérrez Rios; Rosario Vazquez-Larios; Santiago Ávila-Ríos; Selene Zárate; Susana Lopez; Verónica Mata-Haro; Victor Eduardo García-Arias; Víctor Hugo Borja-Aburto |
| EPI_ISL_3347812, EPI_ISL_4006647, EPI_ISL_4878877 | Centro de Investigación Biomédica de Occidente (CIBO) | Instituto de Biotecnología de la UNAM | ; Alejandra García-Gasca; Alejandra Hernández-Terán; Alejandro Sánchez-Flores; Alfredo Herrera-Estrella; Alicia Ocaña-Mondragón; Andreu Comas-García; Angel Gustavo Salas-Lais; Antonio Loza Román; Bernardo Martínez-Miguel; Blanca Taboada; Brenda Irasema Maldonado-Meza; Bruno Gómez-Gil; Carla Ivón Herrera-Najera; Carlos F. Arias; Celia Boukadida; Clara Esperanza Santacruz-Tinoco; Concepción Grajales-Muñiz; Consorcio Mexicano de Vigilancia Genómica (CoViGen-Mex). Authors (in alphabetical order): Julio Elias Alvarado-Yaah; Cristóbal Cháidez-Quiróz; Célida Duque Molina; Célida Martínez-Rodríguez; Daniel Fregoso-Rueda; Daniel Lira Morales; Eduardo Becerril-Vargas; Fernando Fontove-Herrera; Fidencio Mejía-Nepomuceno; Francisco Pulido; Gloria Elena Espinosa-Ayala; Gloria María Molina-Salinas; Gloria Vazquez; Hector Esteban Paz-Juárez; Hector Montoya-Fuentes; Helen Haydee Fernanda Ramírez-Plascencia; Irvin González-López; Jean Pierre González; Jesús Hernández; Joel Armando Vázquez-Pérez).; Jorge Salas-Hernández; José Antonio Enciso-Moreno; José Arturo Martínez-Orozco; José Esteban Muñoz-Medina; José de Jesús Nuñez-Contreras; Juan Bautista Chale-Dzul; Julissa Enciso-Ibarra; Kathia Elizabeth Tapia-Diaz; Luis Alberto Ochoa-Carrera; Margarita Matías-Florentino; Mario Mújica-Sánchez; Marissa Perez-Garcia; María Guadalupe Santiago-Mauricio; María Guadalupe de Jesús Mireles-Rivera; Nelly Sélem-Mojica; Pavel Isa; Ricardo Ciria Merce; Ricardo Grande; Rosa María Gutiérrez Rios; Santiago Ávila-Ríos; Selene Zárate; Susana Lopez; Verónica Mata-Haro; Victor Eduardo García-Arias; Víctor Hugo Borja-Aburto |
| EPI_ISL_3155395, EPI_ISL_4299491, EPI_ISL_5194129, EPI_ISL_5194233, EPI_ISL_5194251 | Centro de Investigación Biomédica del Noreste (CIBIN) | Centro de Investigación en Enfermedades Infecciosas (CIENI), Instituto Nacional de Enfermedades Respiratorias (INER) | ; Alejandra García-Gasca; Alejandra Hernández-Terán; Alejandro Sánchez-Flores; Alfredo Herrera-Estrella; Alicia Ocaña-Mondragón; Andreu Comas-García; Angel Gustavo Salas-Lais; Antonio Loza Román; Bernardo Martínez-Miguel; Blanca Taboada; Brenda Irasema Maldonado-Meza; Bruno Gómez-Gil; Carla Ivón Herrera-Najera; Carlos F. Arias; Celia Boukadida; Clara Esperanza Santacruz-Tinoco; Concepción Grajales-Muñiz; Consorcio Mexicano de Vigilancia Genómica (CoViGen-Mex). Authors (in alphabetical order): Julio Elias Alvarado-Yaah; Cristóbal Cháidez-Quiróz; Célida Duque Molina; Célida Martínez-Rodríguez; Célida Martínez-Rodríguez; Daniel Fregoso-Rueda; Daniel Lira Morales; Eduardo Becerril-Vargas; Eduardo Rivera-Martínez; Fernando Fontove-Herrera; Fidencio Mejía-Nepomuceno; Francisco Pulido; Gabriel Chavira-Trujillo; Gloria Elena Espinosa-Ayala; Gloria María Molina-Salinas; Gloria Vazquez; Hector Esteban Paz-Juárez; Hector Montoya-Fuentes; Helen Haydee Fernanda Ramírez-Plascencia; Irvin González-López; Jean Pierre González; Jesús Hernández; Joel Armando Vázquez-Pérez.; Jorge Salas-Hernández; José Antonio Enciso-Moreno; José Arturo Martínez-Orozco; José Esteban Muñoz-Medina; José de Jesús Nuñez-Contreras; Juan Bautista Chale-Dzul; Julissa Enciso-Ibarra; Kathia Elizabeth Tapia-Diaz; Luis Alberto Ochoa-Carrera; Margarita Matías-Florentino; Mario Mújica-Sánchez; Marissa Perez-Garcia; María Eugenia Jiménez-Corona; María Guadalupe Santiago-Mauricio; María Guadalupe de Jesús Mireles-Rivera; Nelly Sélem-Mojica; Pavel Isa; Ricardo Ciria Merce; Ricardo Grande; Rosa María Gutiérrez Rios; Rosario Vazquez-Larios; Santiago Ávila-Ríos; Selene Zárate; Susana Lopez; Verónica Mata-Haro; Victor Eduardo García-Arias; Víctor Hugo Borja-Aburto |
| EPI_ISL_3347654, EPI_ISL_4006256 | Centro de Investigación Biomédica del Noreste (CIBIN) | Instituto de Biotecnología de la UNAM | ; Alejandra García-Gasca; Alejandra Hernández-Terán; Alejandro Sánchez-Flores; Alfredo Herrera-Estrella; Alicia Ocaña-Mondragón; Andreu Comas-García; Angel Gustavo Salas-Lais; Antonio Loza Román; Bernardo Martínez-Miguel; Blanca Taboada; Brenda Irasema Maldonado-Meza; Bruno Gómez-Gil; Carla Ivón Herrera-Najera; Carlos F. Arias; Celia Boukadida; Clara Esperanza Santacruz-Tinoco; Concepción Grajales-Muñiz; Consorcio Mexicano de Vigilancia Genómica (CoViGen-Mex). Authors (in alphabetical order): Julio Elias Alvarado-Yaah; Cristóbal Cháidez-Quiróz; Célida Duque Molina; Célida Martínez-Rodríguez; Daniel Fregoso-Rueda; Daniel Lira Morales; Eduardo Becerril-Vargas; Fernando Fontove-Herrera; Fidencio Mejía-Nepomuceno; Francisco Pulido; Gloria Elena Espinosa-Ayala; Gloria María Molina-Salinas; Gloria Vazquez; Hector Esteban Paz-Juárez; Hector Montoya-Fuentes; Helen Haydee Fernanda Ramírez-Plascencia; Irvin González-López; Jean Pierre González; Jesús Hernández; Joel Armando Vázquez-Pérez).; Jorge Salas-Hernández; José Antonio Enciso-Moreno; José Arturo Martínez-Orozco; José Esteban Muñoz-Medina; José de Jesús Nuñez-Contreras; Juan Bautista Chale-Dzul; Julissa Enciso-Ibarra; Kathia Elizabeth Tapia-Diaz; Luis Alberto Ochoa-Carrera; Margarita Matías-Florentino; Mario Mújica-Sánchez; Marissa Perez-Garcia; María Guadalupe Santiago-Mauricio; María Guadalupe de Jesús Mireles-Rivera; Nelly Sélem-Mojica; Pavel Isa; Ricardo Ciria Merce; Ricardo Grande; Rosa María Gutiérrez Rios; Santiago Ávila-Ríos; Selene Zárate; Susana Lopez; Verónica Mata-Haro; Victor Eduardo García-Arias; Víctor Hugo Borja-Aburto |
| EPI_ISL_4755987, EPI_ISL_4755991, EPI_ISL_4755992 | China P3 Laboratory | Central Public Health Reference Laboratory | Agnes K. Dumbuya; Alie Tia; Allan K.O. Campbell; Anu Jegede-Williams; Doris Harding; Foday M. Thoronka; Julian Campbell; Prof. Foday Sahr; Prof. Isatta Wurie; Prof. Sahr Gevao; Samuel Sorie; Simon Peter Ruhweza; Warren Cyrus Yiaba; Zein Souma |
| EPI_ISL_4919764 | Clinical Emergency County Hospital, Craiova | Cantacuzino Institute Virology | Carmen Cherciu; Luiza Ustea; Mihaela Lazar; Mihaela Oprea; Nicoleta Paraschiv; Sorin Dinu |
| EPI_ISL_3020158 | Clinical Hospital of Infectious Diseases „Sfanta Parascheva” Iasi | National Institute of Infectious Diseases-Prof. Dr. Matei Bals Molecular Diagnostics Laboratory | Corina Casangiu; Dan Otelea; Leontina Banica; Marius Surleac; Petre Milu; Robert Hohan; Simona Paraschiv |
| EPI_ISL_3988980 | Colorado Department of Public Health and Environment | Colorado Department of Public Health and Environment | Alexandria Rossheim; Diana Ir; Emily A. Travanty; Laura Bankers; Mandy Waters; Michael Martin; Molly C. Hetherington-Rauth; Sarah Elizabeth Totten; Shannon R. Matzinger |
| EPI_ISL_3303926, EPI_ISL_3947115, EPI_ISL_3947448 | Commonwealth Healthcare Center | Centers for Disease Control and Prevention Division of Viral Diseases, Pathogen Discovery | Alex Burgin; Ben L. Rambo-Martin; Ben Rambo-Martin; Clinton Paden; Clinton R. Paden; Dakota Howard; Dave Wentworth; Dhwani Batra; Jasmine Padilla; Justin Lee; Krista Queen; Kristen Knipe; Kristine Lacek; Mark Burroughs; Matthew Schmerer; Meghan Bentz; Mili Sheth; Peter Cook; Sam Shepard; Sarah Nobles; Suxiang Tong; Vivien Dugan; Yvette Unoarumhi |
| EPI_ISL_2932649, EPI_ISL_3462103, EPI_ISL_3663285, EPI_ISL_3663293, EPI_ISL_3914624, EPI_ISL_4105232, EPI_ISL_5032813, EPI_ISL_5032817, EPI_ISL_5032867, EPI_ISL_5032872 | | | |
| see above | Communicable Disease Laboratory, Public Health Directorate | Communicable Disease Laboratory, Public Health Directorate | AlAbbas, Z.; AlHujairi, Z.; Almoamen, G.; Altaif, Z.; Alwasti, H.; Marhoon, A.; Touq, M. |
| EPI_ISL_4030419, EPI_ISL_5439137 | Creighton University Medical Center | Creighton COVID Consortium | Cynthia Watson; Holly Stessman; Jake Siedlik; Michael Belshan; Richard Goering |
| EPI_ISL_4413543, EPI_ISL_4414151 | DASA | DASA | Adriano Bonaldi; Angelica Hristov; Annelise Lopes; Bianca Cota; Cristina Oliveira; Jose Levi; Lidia Yamamoto; Paulo Pierry; Rodrigo Guarischi; Rodrigo Salazar |
| EPI_ISL_3925023 | DH, PORT HEALTH (HARBOUR) | Hong Kong Department of Health | Alan K.L. Tsang; Edman T.K. Lam; Ken H.L. Ng; Peter C.W. Yip; Rickjason C.W. Chan |
| EPI_ISL_2983222 | DNA Labs | National Reference Laboratory, Nigeria Centre for Disease Control | Anthony Ahumibe; Catherine Okoi; Chimaobi Chukwu; Dr Chikwe Ihekweazu; Dr Ndodo Nnaemeka; Dr Omoare Adesuyi; Nwando Mba; Olusola Anuoluwapo Akanbi |
| EPI_ISL_3692638, EPI_ISL_3692762 | DPHL | Delaware Public Health Lab | Rebecca Savage |
| EPI_ISL_5427023, EPI_ISL_5428817, EPI_ISL_5429003, EPI_ISL_5429005, EPI_ISL_5429011 | DSMRC | DSMRC | Khine Zaw Oo; Ko Ko Lwin; Ko Ko Win; Kyee Myint; Nay Myo Aung; Sett Paing Htoo; Thet Wai Oo; Zaw Win Htun |
| EPI_ISL_5314476 | DYOMEDEA-LABORATOIRE DE LA SAUVEGARDE | CNR Virus des Infections Respiratoires - France SUD | Antonin Bal; Bruno Lina; Gregory Destras; Gwendolyne Burfin; Hadrien Regue; Laurence Josset; Martine Valette; Quentin Semanas |
| EPI_ISL_5428720 | Department Medical Microbiology, Baerum Hospital, Vestre Viken Health Trust | Norwegian Institute of Public Health, Department of Virology | Atiya R Ali; Debech Nadia; Engebretsen Serina Beate; Garcia Llorente Ignacio; Hilde Elshaug; Hilde Vollan; Jon Bråte; Kamilla Heddeland Instefjord; Karoline Bragstad; Kathrine Stene-Johansen; Line Victoria Moen; Marie Paulsen Madsen; Olav Hungnes; Pedersen Benedikte Nevjen; Rasmus Riis Kopperud |
| EPI_ISL_4572477, EPI_ISL_4659774 | Department for Virology, Molecular Biology and Genome Research, R. G. Lugar Center for Public Health Research, National Center for Disease Control and Public Health (NCDC) of Georgia. | Department for Virology, Molecular Biology and Genome Research, R. G. Lugar Center for Public Health Research, National Center for Disease Control and Public Health (NCDC) of Georgia. | Adam Kotorashvili; Amiran Gamkrelidze.; Ana Papkiauri; Ann Machablishvili; Anna Kasradze; Davit Tsaguria; Ekaterine Khmaladze; Ekaterine Zangaladze; Ekaterine Zhghenti; Giorgi Gogoladze; Giorgi Tomashvili; Gvantsa Brachveli; Gvantsa Chanturia; Irma Burjanadze; Ketevan Sidamonidze; Khatuna Zakhashvili; Lela Sabadze; Lela Urushadze; Magda Dgebuadze; Maia Alkhazashvili; Mari Gavashelidze; Mariam Zakalashvili; Marine Murtskhvaladze; Meri Pantsulaia; Nato Kotaria; Nino Berishvili; Paata Imnadze; Roena Sukhiashvili; Salome Javashvili; Tamar Jashiashvili; Tata Imnadze; Tea Tevdoradze |
| EPI_ISL_2840855, | Department of Acute Infectious | Department of Acute Infectious | Jienan Zhou; Meiling Zhang; Senquan Jia; Xiaoqing Fu; Zhaosheng Liu |

| | | | |
|---|---|---|---|
| EPI_ISL_2896219 | Diseases Control and Prevention, Yunnan Provincial Center for Disease Control and Prevention | Diseases Control and Prevention, Yunnan Provincial Center for Disease Control and Prevention | |
| EPI_ISL_2668554, EPI_ISL_2784372, EPI_ISL_3028864, EPI_ISL_3072449, EPI_ISL_3073623, EPI_ISL_3103102, EPI_ISL_3127104, EPI_ISL_3162633, EPI_ISL_3280281, EPI_ISL_3989543, EPI_ISL_4050522, EPI_ISL_4355529, EPI_ISL_4448829, EPI_ISL_4942972, EPI_ISL_5173423, EPI_ISL_5174757 | | | |
| see above | Department of Bacteria, Parasites and Fungi, Statens Serum Institut, Copenhagen, Denmark | Statens Serum Institut Bioinformatics and Microbial Genomics | Danish Covid-19 Genome Consortium |
| EPI_ISL_4968439 | Department of Health Technology and Informatics, The Hong Kong Polytechnic University | Department of Health Technology and Informatics, The Hong Kong Polytechnic University | Alan Ka-Lun Wu; Alex Yat-Man Ho; Barry Kin-Chung Wong; Chloe Toi-Mei Chan; David Ho-Keung Shum; Denise Sze-Hang Wong; Gilman Kit-Hang Siu; Hiu-Yin Lao; Hoi-Ching Jim; Ivan Tak-Fai Wong; Jake Siu-Lun Leung; Kam-Tong Yip; Kenneth Siu-Sing Leung; Kingsley King-Gee Tam; Kitty Sau-Chun Fung; Kristine Luk; Lam-Kwong Lee; Miranda Chong-Yee Yau; Sandy Ka-Yee Chau; Shea Ping Yip; Tak-Lun Que; Timothy Ting-Leung Ng; Wing Cheong Yam; Wing-Hei Lo; Wing-Kin To; Yvette Wai-Man Lai |
| EPI_ISL_3505808 | Department of Microbiology, National Institute for Public Health of Kosova | Charité Universitätsmedizin Berlin, Institut für Virologie | Aferdita Hyseni; Barbara Mühlemann; Blendi Jerliu; Christian Drosten; Donjeta Hajdari; Julia Schneider; Jörn Beheim-Schwarzbach; Nazmi Mehmeti; Pranvera Abazi; Talitha Veith; Terry Jones; Victor M Corman; Xhevat Jakupi; Zana Deva |
| EPI_ISL_4545719, EPI_ISL_4545762, EPI_ISL_4550688, EPI_ISL_4550715, EPI_ISL_5144526, EPI_ISL_5429540, EPI_ISL_5429558, EPI_ISL_5429643, EPI_ISL_5429645 | | | |
| see above | Department of Microbiology, National Institute of Public Health of Kosovo | Department of Microbiology, National Institute of Public Health of Kosovo | Aferdita Hyseni; Aferdita Kuqi-Hyseni; Blendi Jerliu; Donjeta Hajdari; Nazmi Mehmeti; Pranvera Abazi; Robert Ramadani; Xhevat Jakupi; Zana Deva |
| EPI_ISL_5105922 | Department of Public Health Braila | National Institute of Infectious Diseases-Prof. Dr. Matei Bals Molecular Diagnostics Laboratory | Corina Casangiu; Dan Otelea; Leontina Banica; Marius Surleac; Ovidiu Vlaicu; Petre Milu; Robert Hohan; Simona Paraschiv |
| EPI_ISL_4458432, EPI_ISL_4458433, EPI_ISL_4630046, EPI_ISL_4630049, EPI_ISL_5427759 | Department of Virology | Department of Virology | Aamer Ikram; Massab Umair; Muhammad Ammar; Muhammad Salman; Nazish Badar; Syed Adnan Haider; Zaira Rehman |
| EPI_ISL_3134993, EPI_ISL_3644691, EPI_ISL_4092397, EPI_ISL_4093109, EPI_ISL_4093580, EPI_ISL_5161225, EPI_ISL_5161265, EPI_ISL_5161298 | | | |
| see above | Department of Virology and Immunology, University of Helsinki and Helsinki University Hospital, Huslab Finland | Department of Virology, Faculty of Medicine, University of Helsinki, Helsinki, Finland | Essi Korhonen; Hanna Jarva; Hanna Liimatainen; Hannimari Kallio-Kokko; Harri Kangas; Hussein Alburkat; Jenni Virtanen; Maija Lappalainen; Maija Suvanto; Olli Vapalahti; Pekka Ellonen; Phuoc Truong; Ravi Kant; Sari Hannula; Satu Kurkela; Teemu Smura |
| EPI_ISL_4999592, EPI_ISL_5314664 | Dept. of Microbiology and Infection Control, Akershus University Hospital HF | Dept. of Microbiology and Infection Control, Akershus University Hospital HF | Alexander Hesselberg Løvestad; Hege Vangstein Aamot |
| EPI_ISL_3233253, EPI_ISL_5331704 | Diagnostyka. Laboratoria Medyczne. | 1. ViroGenetics - BSL3 Laboratory of Virology, Małopolska Centre of Biotechnology, Jagiellonian University; 2. genXone SA, Research & Development Laboratory | Aleksandra Gidlewicz; Anna Brylak; Gromowski, T.; Grzegorz Nowicki; Jakub Grabowski; Karol Szeszko; Kowalski, M.; Labaj; Maciej Sykulski; Mazur-Panasiuk, N.; Michał Kaszuba; Natalia Drweska-Matelska; P.P.; Pyrc, K.; Ruslan Herasymenko; Sylwia Januszczak; Szulc, P.; Wydmanski, W.; Łukasz Krych |
| EPI_ISL_3260960, EPI_ISL_3547072 | Dipartimento di Medicina di Laboratorio, Azienda sanitaria universitaria Friuli Centrale (ASU FC) | Dipartimento di Medicina di Laboratorio, Azienda sanitaria universitaria Friuli Centrale (ASU FC) | Catia Mio; Chiara Dal Secco; Corrado Pipan; Francesco Curcio; Stefania Marzinotto |
| EPI_ISL_3544888, EPI_ISL_3870067, EPI_ISL_4204353 | Division of Emerging Infectious Diseases, Bureau of Infectious Diseases Diagnosis Control, Korea Disease Control and Prevention Agency | Division of Emerging Infectious Diseases, Bureau of Infectious Diseases Diagnosis Control, Korea Disease Control and Prevention Agency | Ae Kyung Park; Chae Young Lee; Eun-Jin Kim; Heui Man Kim; Il-Hwan Kim; Jeong-Ah Kim |
| EPI_ISL_3827777 | Division of Medical Virology, National Health Laboratory Service (NHLS), Tygerberg Hospital / Stellenbosch University | CERI, Centre for Epidemic Response and Innovation, Stellenbosch University and CERI-KRISP, KZN Research Innovation and Sequencing Platform | Alvera Vorster; Bronwyn Kleinhans; Carel J van Heerden; Gert van Zyl; Giandhari Jennifer; Kamela Mahlakwane; Karabo Phadu; Mathilda Claassen; Naidoo Yeshnee; Pillay Sureshnee; Ren Veikondis; San James; Shannon Wilson; Susan Engelbrecht; Tania Stander; Tegally Houriiyah; Tongai Maponga; Tshiabuila Derek; Wilkinson Eduan; Wolfgang Preiser; Yajna Ramphal; de Oliveira Tulio |
| EPI_ISL_3847727, EPI_ISL_3856288, EPI_ISL_5018697, EPI_ISL_5264685, EPI_ISL_5264691, EPI_ISL_5264709, EPI_ISL_5264713 | | | |
| see above | Division of Medical Virology, National Health Laboratory Service (NHLS), Tygerberg Hospital / Stellenbosch University | Division of Medical Virology, National Health Laboratory Service (NHLS), Tygerberg Hospital / Stellenbosch University | Bronwyn Kleinhans; Gert van Zyl; Kamela Mahlakwane; Shannon Wilson; Susan Engelbrecht; Tongai Maponga; Wolfgang Preiser |
| EPI_ISL_2379633 | Dr S Raju, Director of Public Heath and Preventive Medicine | inStem NCBS - INSACOG | Uma Ramakrishnan Dasaradhi Palakodeti Aswin SaiNarain |
| EPI_ISL_5428891 | Dr. Risch AG | Microbiology | Dominique Fabien Hilti; Faina Wehrli; Lorenz Risch; Martin Risch; Nadia Wohlwend; Sinem Kas; Thomas Bodmer |
| EPI_ISL_5069763, EPI_ISL_5162567, EPI_ISL_5162603, EPI_ISL_5428900 | Dr. Risch Ostschweiz AG | Microbiology | Dominique Fabien Hilti; Faina Wehrli; Lorenz Risch; Martin Risch; Nadia Wohlwend; Sinem Kas; Thomas Bodmer |
| EPI_ISL_3388900, EPI_ISL_3389002, EPI_ISL_3570034, EPI_ISL_3635714, EPI_ISL_3798258, EPI_ISL_3798322 | Dr. Risch Ostschweiz AG | Microbiology, Dr. Risch | Dominique Fabien Hilti; Faina Wehrli; Lorenz Risch; Martin Risch; Nadia Wohlwend; Sinem Kas; Thomas Bodmer |
| EPI_ISL_5079479 | Dr. Risch Ostschweiz AG | Microbiology_DrRisch_Buchs | Dominique Fabien Hilti; Faina Wehrli; Lorenz Risch; Martin Risch; Nadia Wohlwend; Sinem Kas; Thomas Bodmer |
| EPI_ISL_2981159, EPI_ISL_2981892, EPI_ISL_3056741, EPI_ISL_3056793, EPI_ISL_3057817, EPI_ISL_3136570, EPI_ISL_3137118, EPI_ISL_3138355, EPI_ISL_3138372, EPI_ISL_3138380, EPI_ISL_3138413, EPI_ISL_3138425, EPI_ISL_3257603, EPI_ISL_3257604, EPI_ISL_3257872, EPI_ISL_3257896, EPI_ISL_3257937, EPI_ISL_3257952, EPI_ISL_3257995, EPI_ISL_3259218, EPI_ISL_3259332, EPI_ISL_3389389, EPI_ISL_3389756, EPI_ISL_3389790, EPI_ISL_3390528, EPI_ISL_3390689, EPI_ISL_3731686, EPI_ISL_3731809, EPI_ISL_3732496, EPI_ISL_3732938, EPI_ISL_3733687, EPI_ISL_3733717, EPI_ISL_3733944, EPI_ISL_3917483, EPI_ISL_3917836, EPI_ISL_3918545, EPI_ISL_3918557, EPI_ISL_3918559, EPI_ISL_3918567, EPI_ISL_3918593, EPI_ISL_3918600, EPI_ISL_3918602, EPI_ISL_3918619, EPI_ISL_3918622, EPI_ISL_3918908, EPI_ISL_4075890, EPI_ISL_4076734, EPI_ISL_4076777, EPI_ISL_4076895, EPI_ISL_4077062, EPI_ISL_4077079, EPI_ISL_4401867, EPI_ISL_4401875, EPI_ISL_4543474, EPI_ISL_4543475, EPI_ISL_4543610, EPI_ISL_4543994, EPI_ISL_4838557, EPI_ISL_4839300, EPI_ISL_4839429, EPI_ISL_4839499, EPI_ISL_4839626, EPI_ISL_4846030, EPI_ISL_4846106, EPI_ISL_4846188, EPI_ISL_4992790, EPI_ISL_4992871, EPI_ISL_4993282, EPI_ISL_4993433, EPI_ISL_4993614, EPI_ISL_4993754, EPI_ISL_5209094, EPI_ISL_5209098, EPI_ISL_5209099, EPI_ISL_5209113, EPI_ISL_5209114, EPI_ISL_5209128, EPI_ISL_5209130, EPI_ISL_5209132, EPI_ISL_5209143, EPI_ISL_5209239, EPI_ISL_5209545, EPI_ISL_5209562, EPI_ISL_5429663, EPI_ISL_5429667, EPI_ISL_5429668, EPI_ISL_5429672, EPI_ISL_5429674, EPI_ISL_5429679, EPI_ISL_5429682, EPI_ISL_5429687, EPI_ISL_5429688, EPI_ISL_5429696, EPI_ISL_5429709, EPI_ISL_5429739, EPI_ISL_5429765, EPI_ISL_5429766, EPI_ISL_5429773, EPI_ISL_5429824, EPI_ISL_5430070, EPI_ISL_5430156, EPI_ISL_5430661 | | | |
| see above | Dutch COVID-19 response team | National Institute for Public Health and the Environment (RIVM) | Adam Meijer; AnneMarie van den Brandt; Annelies Kroneman; Bas van der Veer; Chantal Reusken; Dennis Schmitz; Dirk Eggink; Euníce Then; Florian Zwagemaker; Harry Vennema; Ivo van Walle; Jeroen Cremer; Karim Hajji; Kim Freriks; Lisa Wijsman; Lynn Aarts; Melissa van Tuil; Ryanne Jaarsma; Sanne Bos; Sharon van den Brink; Stijn van Rossum; on behalf of the national COVID-19 response team |
| EPI_ISL_4548624 | E. Gulbja Laboratorija | Latvian Biomedical Research and Study Centre | Davids Fridmanis; Dmitrijs Perminovs; Elina Dimina; Guntars Zarins; Ivars Silamikelis; Janis Klovins; Janis Pjalkovskis; Jurijs Perevoscikovs; Kaspars Megnis; Laila Silamikele; Lauma Freimane; Laura Ansone; Liga Birzniece; Mikus Gavars; Monta Briviba; Nikita Zrelovs; Uga Dumpis; Una Krumina; Vita Rovite |
| EPI_ISL_4054992, EPI_ISL_4561723 | Edmonton Provincial Lab | Public Health Agency of Canada (PHAC) National Microbiology Laboratory | Buss; Croxen M; Deo A; Dieu P; E; Ferrato C; Gill K; Khan F; Koleva P; Li V; Lloyd C; Lynch T; Ma R; Murphy S; Pabbaraju K; Shokoples S; Thayer J; Tipples G; Whitehouse M; Wong A; Yu C; Zelyas N |
| EPI_ISL_4490404, EPI_ISL_4768588 | Ekiti State Emergency Operation Centre | Africa Centre for Excellence for Genomics of Infectious Diseases (ACEGID), Redeemer's University | A.T.; Abechi; Ajogbasile; Akano; C.A.; C.T.; Eromon; F.V.; Folarin, O.; Happi; I.B.; J.N.; J.U.; K.O.; Kayode; Nosamiefan, I.; Oguzie; Olawoye; Olumade; Oluniyi; P.E.; P.S.; T.J.; Ugwu; Uwanibe |
| EPI_ISL_5409537 | Enfer Medical | ELDA biotech | Elaine M. Kenny |
| EPI_ISL_4819760, EPI_ISL_4819825, EPI_ISL_4819872, EPI_ISL_4819903, EPI_ISL_4819952, EPI_ISL_4819978, EPI_ISL_4819992, EPI_ISL_4820106, EPI_ISL_4820113, EPI_ISL_4820242, EPI_ISL_4820264 | | | |
| see above | Fiji Centre for Communicable Disease Control | Microbiological Diagnostic Unit - Public Health Laboratory (MDU-PHL) | Cabemaiwai, T.; Faktaufon, D.; Horan, K.; N.L.; Sahukhan, A.; Seemann, T.; Sherry; Singh, S. |
| EPI_ISL_3020940 | Fimlab Laboratoriot Oy Tampere | Expert Microbiology, National Institute for Health and Welfare | Carita Savolainen-Kopra; Erika Lindh; Haider al-Hello; Jani Halkilahti; Kirsi Liitsola; Niina Ikonen; Olli Vapalahti; Pekka Ellonen; Phuoc Truong; Päivi Laurila; Ravi Kant; Sari Hannula; Soile Blomqvist; Teemu Smura |
| EPI_ISL_2889854 | Fondation Congolaise pour la recherche medicale (FCRM) | Fondation Congolaise pour la Recherche Médicale | Abel Lissom; Batchi-Bouyou Armel Landry; Francine Ntoumi; Jean Claude Djontu; Mfoutou Mapanguy Claujens Chastel; Thirumalaisamy P. Velavan |
| EPI_ISL_3040129, EPI_ISL_3040130, EPI_ISL_4572232, EPI_ISL_4572263, EPI_ISL_4574522, EPI_ISL_4724347, EPI_ISL_4724354, EPI_ISL_4724402, EPI_ISL_4724408 | | | |

| | | | |
|---|---|---|---|
| see above | Fondation Congolaise pour la recherche medicale (FCRM), Francine Ntoumi | Fondation Congolaise pour la Recherche Médicale | Batchi-Bouyou Armel Landry; Dr. Abel Lissom; Dr. Jean Claude Djontu; Mfoutou Mapanguy Claujens Chastel; Prof. Dr. Thirumalaisamy P. Velavan; Prof. Francine Ntoumi |
| EPI_ISL_3100575 | Fondazione IRCCS Ca' Granda Ospedale Maggiore Policlinico | Fondazione IRCCS Ca' Granda Ospedale Maggiore Policlinico | Ferruccio Ceriotti; Sara Uceda Renteria |
| EPI_ISL_3345000, EPI_ISL_3349172, EPI_ISL_4034874, EPI_ISL_4901517, EPI_ISL_5092595, EPI_ISL_5231980, EPI_ISL_5307952, EPI_ISL_5307954, EPI_ISL_5307975, EPI_ISL_5318987, EPI_ISL_5319085 | | | |
| see above | Fulgent Genetics | Centers for Disease Control and Prevention Division of Viral Diseases, Pathogen Discovery | Adrian Paskey; Becky Tsai; Benafsh Sapra; Benjamin Rambo-Martin; Christopher Gulvick; Clinton Paden; Clinton R. Paden; Dakota Howard; Darlene Wagner; Dhwani Batra; Doreen Ng; Duncan MacCannell; Erisa Sula; Harry Gao; James Xie; Jason Caravas; John Gao; Joseph Fierro; Kara Moser; Kristine Lacek; Matthew Schmerer; Mickey Li; Peter Cook; Peter W. Cook; Scott Sammons; Shatavia Morrison; Tymeckia Kendall; Victoria Caban Figueroa; Yan Meng; Yvette Unoarumhi |
| EPI_ISL_2927810 | Furst Medical Laboratory | Norwegian Institute of Public Health, Department of Virology | Atiya R Ali; Debech Nadia; Engebretsen Serina Beate; Garcia Llorente Ignacio; Hilde Elshaug; Hilde Vollan; Jon Bråte; Kamilla Heddeland Instefjord; Karoline Bragstad; Kathrine Stene-Johansen; Line Victoria Moen; Marie Paulsen Madsen; Olav Hungnes; Pedersen Benedikte Nevjen; Rasmus Riis Kopperud |
| EPI_ISL_5058669 | Gandhi Medical College | CDFD | Arunkumar Karunanidhi; Ashwin Dalal; Asmita Gupta; Divya Vashisht; Murali Bashyam; Nagamani Kammili; Vinay Donipadi |
| EPI_ISL_406798 | General Hospital of Central Theater Command of People's Liberation Army of China | BGI & Institute of Microbiology, Chinese Academy of Sciences & Shandong First Medical University & Shandong Academy of Medical Sciences & General Hospital of Central Theater Command of People's Liberation Army of China | Weifeng Shi and Zhenhong Hu; Weijun Chen; Yuhai Bi |
| EPI_ISL_4636133, EPI_ISL_4656218, EPI_ISL_5431370, EPI_ISL_5431371, EPI_ISL_5431413, EPI_ISL_5431446, EPI_ISL_5431477, EPI_ISL_5431538 | | | |
| see above | Genetica Molecular and Subdepartamento de Virologia ISP Chile | Instituto de Salud Publica de Chile | Andres Castillo; Barbara Parra; Constanza Campano; Gisselle Barra; Javier Tognarelli; Jorge Fernandez; Karen Orostica; Loredana Arata; Patricia Bustos; Rodrigo Fasce; Soledad Ulloa |
| EPI_ISL_5305440 | Global Medical Center | Virology Lab, Jaber Al Ahmad Hospital | Dr. Ebaa Al-Awadhi; Dr. Zahrah Buhamad; Estabraq Kathim; Haroon Masih; Khubaid-ur-Rehman |
| EPI_ISL_2985778, EPI_ISL_5115557 | Gravity Diagnostics, LLC | Gravity Diagnostics, LLC | Gravity Diagnostics |
| EPI_ISL_5313333 | Greek Genome Center, Biomedical Research Foundation of the Academy of Athens (BRFAA) | Greek Genome Center, Biomedical Research Foundation of the Academy of Athens (BRFAA) | Dimitrios Thanos; Dimitris Vrachnos; Emmanouil Athanasiadis; Giannis Vatsellas; Katerina Zoi; Theodoros Loupis |
| EPI_ISL_3506467 | Groote Schuur Hospital wc GSH | NHLS/UCT | Arash Iranzadeh; Bruna Galvao; Carolyn Williamson; Deelan Doolabh; Diana Hardie; Gert Marais; Innocent Mudau; Lynn Tyers; Marvin Hsiao; Rageema Joseph; Stephen Korsman |
| EPI_ISL_4539468, EPI_ISL_4539521 | Guam Public Health Laboratory | Centers for Disease Control and Prevention Division of Viral Diseases, Pathogen Discovery | Alex Burgin; Ben Rambo-Martin; Clinton Paden; Dakota Howard; Dave Wentworth; Dhwani Batra; Jasmine Padilla; Justin Lee; Krista Queen; Kristen Knipe; Kristine Lacek; Mark Burroughs; Matthew Schmerer; Meghan Bentz; Mili Sheth; Peter Cook; Sam Shepard; Sarah Nobles; Suxiang Tong; Vivien Dugan; Yvette Unoarumhi |
| EPI_ISL_5438976 | HLAGYN - Laboratorio de Imunologia de Transplantes de Goias | HLAGYN - Laboratorio de Imunologia de Transplantes de Goias | Alessandro Leonardo Alvares Magalhaes; Erika Lopes Rocha Batista; Fernando Antonio Vinhal dos Santos; Frederico Rodrigues Vinhal; Kamila Oliveira Reis De Freitas.; Lucas Carlos Gomes Pereira; Sabrina Sara Moreira Duarte |
| EPI_ISL_4601220 | HOSPITAL DE CIUDAD NEILY | Inciensa, Instituto Costarricense de Investigación y Enseñanza en Nutrición y Salud | Adriana Godínez; Claudio Soto-Garita; Estela Cordero; Francisco Duarte; Hebleen Porras; Joselyn Prado & Daniel Solano Sanchez; José Luis Vargas; Mariela Gutiérrez; Melany Calderón |
| EPI_ISL_5262697 | HOSPITAL DR. RAFAEL ANGEL CALDERON GUARDIA | Inciensa, Instituto Costarricense de Investigación y Enseñanza en Nutrición y Salud | Adriana Godínez; Claudio Soto-Garita; Estela Cordero; Francisco Duarte; Hebleen Porras; José Luis Vargas; Mariela Gutiérrez; Melany Calderón; Sofia Herrera & Brenda Sánchez Cabezas |
| EPI_ISL_3948542 | HOSPITAL GOLFITO MANUEL MORA VALVERDE | Inciensa, Instituto Costarricense de Investigación y Enseñanza en Nutrición y Salud | Adriana Godínez; Claudio Soto-Garita; Estela Cordero; Francisco Duarte; Hebleen Porras; José Luis Vargas; Mariela Gutiérrez & Joselyn Prado; Melany Calderón |
| EPI_ISL_4168723 | HOSPITAL MUNICIPAL DE ILHABELA GOV MARIO COVAS JR | Instituto Butantan | Antonio Jorge Martins; Claudia Renata dos Santos Barros; David Schlesinger; Debora Botequio Moretti; Dimas Tadeu Covas; Elaine Cristina Marqueze; Elaine Vieira Santos; Evandra Strazza Rodrigues; Heidge Fukumasu; Jayme Augusto de Souza-Neto; José Salvatore Leister Patané; Luiz Alcantara; Luiz Lehmann Coutinho; Maria Carolina Elias; Maurício Lacerda Nogueira; Rafael dos Santos Bezerra; Raul Machado Neto; Rejane Maria Tommasini Grotto; Ricardo Haddad; Sandra Coccuzzo Sampaio Vessoni; Simone Kashima; Svetoslav Nanev Slavov; Vincent Louis Viala |
| EPI_ISL_3761783 | HOSPITAL NACIONAL DE NIÑOS | Inciensa, Instituto Costarricense de Investigación y Enseñanza en Nutrición y Salud | Cristian Pérez-Corrales & Valeria Peralta-Barquero |
| EPI_ISL_3639005 | HOSPITAL SAN FRANCISCO DE ASIS | Inciensa, Instituto Costarricense de Investigación y Enseñanza en Nutrición y Salud | Adriana Godínez; Claudio Soto-Garita; Estela Cordero; Francisco Duarte; Hebleen Porras; Joselyn Prado & Adrián Fallas Mora; José Luis Vargas; Mariela Gutiérrez; Melany Calderón |
| EPI_ISL_3375173 | Haukeland University Hospital, Dept. of Microbiology | Norwegian Institute of Public Health, Department of Virology | Atiya R Ali; Debech Nadia; Engebretsen Serina Beate; Garcia Llorente Ignacio; Hilde Elshaug; Hilde Vollan; Jon Bråte; Kamilla Heddeland Instefjord; Karoline Bragstad; Kathrine Stene-Johansen; Line Victoria Moen; Marie Paulsen Madsen; Olav Hungnes; Pedersen Benedikte Nevjen; Rasmus Riis Kopperud |
| EPI_ISL_2829265 | Health Services Laboratories | Wellcome Sanger Institute for the COVID-19 Genomics UK (COG-UK) Consortium | Cordelia Langford; David K. Jackson; Dominic Kwiatkowski; Ewan Harrison; Health Services Laboratories and Alex Alderton; Ian Johnston; Jeffrey Barrett; John Sillitoe on behalf of the Wellcome Sanger Institute COVID-19 Surveillance Team; Roberto Amato; Sonia Goncalves |
| EPI_ISL_5203041, EPI_ISL_5203906, EPI_ISL_5240436, EPI_ISL_5240695, EPI_ISL_5240708 | Helix | Centers for Disease Control and Prevention Division of Viral Diseases, Pathogen Discovery | Benjamin Rambo-Martin; Christopher Gulvick; Clinton Paden; Dakota Howard; Dhwani Batra; Duncan MacCannell; Erisa Sula; Helix CA; Jason Caravas; Kristine Lacek; Matthew Schmerer; Peter Cook; Scott Sammons; Shatavia Morrison; Tymeckia Kendall; Victoria Caban Figueroa; Yvette Unoarumhi |
| EPI_ISL_3689138, EPI_ISL_3689188 | Hellenic National Blood Transfusion Center - EKEA | Greek Genome Center, Biomedical Research Foundation of the Academy of Athens (BRFAA) | Dimitrios Thanos; Efthimia Petinaki; Emmanouil Athanasiadis; Giannis Vatsellas; Katerina Zoi; Kostas Stamoulis; Theodoros Loupis |
| EPI_ISL_5064720 | Hetauda Hospital | National Public Health Laboratory | National Public Health Laboratory Team |
| EPI_ISL_3398580, EPI_ISL_3833894, EPI_ISL_5305692 | Histopath | NSW Health Pathology - Institute of Clinical Pathology and Medical Research; Westmead Hospital; University of Sydney | Arnott A.; Draper J.; Gall M.; Martinez E.; Rockett R.; Sintchenko V.; on behalf of ICPMR |
| EPI_ISL_5332931 | Home Quarantine Taskforce | Hong Kong Department of Health | Alan K.L. Tsang; Edman T.K. Lam; Ken H.L. Ng; Peter C.W. Yip; Rickjason C.W. Chan |
| EPI_ISL_3542891, EPI_ISL_3542959, EPI_ISL_4236927 | Hospital | National Reference Center for Viruses of Respiratory Infections, Institut Pasteur, Paris | Angela Brisebarre; Camille Capel; Christophe Malabat; Corinne Maufrais; Didier Mattera; Etienne Simon-Lorière; Frédéric Lemoine; Hub de Bioinformatique et Biostatistique; Julien Fumey; Louise Lefrançois; Marion Barbet; Maud Vanpeene; Méline Bizard; Olivier Dejoux; Slim El Khiari; Sylvie Behillil; Sylvie Van der Werf; Vincent Enouf |
| EPI_ISL_5417637 | Hospital Canselor Tuanku Muhriz (HCTM) | UKM Medical Molecular Biology Institute (UMBI) | Mira Farzana binti Mohamad Mokhtar |
| EPI_ISL_3147656 | Hospital Center Emile Mayrisch | Laboratoire national de sante, Microbiology, Microbial Genomics Platform | Anke Wienecke-Baldacchino; Catherine Ragimbeau; Cynthia Oxacelay; Elodie Solarino; Fatu Djabi; Jessica Tapp; Lise Pignon; Raoul Salmon; Tamir Abdelrahman; Virginie Jover |
| EPI_ISL_5329478, EPI_ISL_5329529, EPI_ISL_5329732 | Hospital General Ajusco Medio | Instituto Nacional de Medicina Genomica | Cedro-Tanda A; Cruz-Islas Jazmin; Escobar-Arrazola MA; Garnica-Lopez Dora; Herrera-Montalvo LA.; Hidalgo-Miranda A; Mendoza-Vargas A; Ramirez-Vega O; Rangel-DeLeon D; Reyes-Grajeda JP; Yair Alfaro-Mora |
| EPI_ISL_4395995 | Hospital General Universitario Gregorio Marañón | Hospital General Universitario Gregorio Marañón | Cristina Rodriguez-Grande; Darío García de Viedma; Julia Suárez; Laura Pérez-Lago; Marta Herranz Martin; Patricia Muñoz; Pedro Sola Campoy; Pilar Catalán; Sergio Buenestado Serrano; Victor Manuel de la Cueva |
| EPI_ISL_5055104, EPI_ISL_5055105 | Hospital Margarita Maza de Juárez | Microbial Genomics Laboratory | ; Alejandra García-Gasca; Alejandra Hernández-Terán; Alejandro Sánchez-Flores; Alfredo Herrera-Estrella; Alicia Ocaña-Mondragón; Andreu Comas-García; Angel Gustavo Salas-Lais; Antonio Loza Román; Bernardo Martínez-Miguel; Blanca Taboada; Brenda Irasema Maldonado-Meza; Bruno Gómez-Gil; Carla Ivón Herrera-Najera; Carlos F. Arias; Celia Boukadida; Clara Esperanza Santacruz-Tinoco; Concepción Grajales-Muñiz; Consorcio Mexicano de Vigilancia Genómica (CoViGen-Mex). Authors (in alphabetical order): Julio Elias Alvarado-Yaah; Cristóbal Cháidez-Quiróz; Célida Duque Molina; Célida Martínez-Rodríguez; Daniel Fregoso-Rueda; Daniel Lira Morales; Eduardo Becerril-Vargas; Fernando Fontove-Herrera; Fidencio Mejía-Nepomuceno; Francisco Pulido; Gloria Elena Espinosa-Ayala; Gloria María Molina-Salinas; Gloria Vazquez; Hector Esteban Paz-Juárez; Hector Montoya-Fuentes; Helen Haydee Fernanda Ramírez-Plascencia; Irvin González-López; Jean Pierre González; Jesús Hernández; Joel Armando Vázquez-Pérez.; Jorge Salas-Hernández; José Antonio Enciso-Moreno; José Arturo Martínez-Orozco; José Esteban Muñoz-Medina; José de Jesús Nuñez-Contreras; Juan Bautista Chale-Dzul; Julissa Enciso-Ibarra; Luis Alberto Ochoa-Carrera; Margarita Matías-Florentino; Mario Mújica-Sánchez; Marissa Perez-Garcia; María Guadalupe Santiago-Mauricio; María Guadalupe de Jesús Mireles-Rivera; Nelly Sélem-Mojica; Pavel Isa; Ricardo Ciria Merce; Ricardo Grande; Rosa María Gutiérrez Rios; Santiago Ávila-Ríos; Selene Zárate; Susana Lopez; Verónica Mata-Haro; Victor Eduardo García-Arias; Víctor Hugo Borja-Aburto |
| EPI_ISL_3374758 | Hospital Universitari Arnau de Vilanova | Hospital Universitari Vall d'Hebron - Vall d'Hebron Institut de Recerca | Alejandra González-Sánchez; Andrés Antón; Ariadna Rando; Carla Castillo; Cristina Andrés; Damir Garcia-Cehic; Josep Quer; Juliana Esperalba; Karen García; Maria Carmen Martin; Maria Gema Codina; Maria Piñana; Rodrigo Vásquez; Tomàs Pumarola |
| EPI_ISL_3061797, EPI_ISL_3401938, | Hrvatski zavod za javno zdravstvo | Hrvatski zavod za javno zdravstvo | Irena Tabain; Ivana Ferenčak |

| | | | |
|---|---|---|---|
| EPI_ISL_5012937, EPI_ISL_5196141 | | | |
| EPI_ISL_4461318 | IMP - Research Institute of Molecular Pathology | Bergthaler laboratory, CeMM Research Center for Molecular Medicine of the Austrian Academy of Sciences | Andreas Bergthaler; Anna Schedl; Bekir Erguner; Benedikt Agerer; Christoph Bock; Fabian Amman; Jan Laine; Lukas Endler; Maelle Le Moing; Martin Senekowitsch; Matthew Thornton; Michael Schuster; Petr Triska; Thomas Penz |
| EPI_ISL_3098780, EPI_ISL_5406133 | IN State Department of Health Laboratory Services | IN State Department of Health Laboratory Services | Brian Pope; Cassandra Campion; Jamie Yeadon; Kyle Brownlee; Lixia Liu; Mark Glazier; Melissa Hindenlang |
| EPI_ISL_3189423 | INSACOG-Mizoram | National Institute of Biomedical Genomics – INSACOG | Arindam Maitra; Gracy Laldinmawii; N Senthil Kumar; Nidhan Kumar Biswas; Saumitra Das; Sreedhar Chinnaswamy; Swagnik Roy |
| EPI_ISL_2676033 | INSACOG-WB | National Institute of Biomedical Genomics – INSACOG | Ajay Chakraborti; Arindam Maitra; Bhaswati Bandyopadhyay; Nidhan Kumar Biswas; Saumitra Das; Sreedhar Chinnaswamy; Tamal Ghosh |
| EPI_ISL_4469851 | INSIDE DIAGNÓSTICOS | Instituto Butantan | Antonio Jorge Martins; Claudia Renata dos Santos Barros; David Schlesinger; Debora Botequio Moretti; Dimas Tadeu Covas; Elaine Cristina Marqueze; Elaine Vieira Santos; Evandra Strazza Rodrigues; Heidge Fukumasu; Jayme Augusto de Souza-Neto; José Salvatore Leister Patané; Luiz Alcantara; Luiz Lehmann Coutinho; Maria Carolina Elias; Maurício Lacerda Nogueira; Rafael dos Santos Bezerra; Raul Machado Neto; Rejane Maria Tommasini Grotto; Ricardo Haddad; Sandra Coccuzzo Sampaio Vessoni; Simone Kashima; Svetoslav Nanev Slavov; Vincent Louis Viala |
| EPI_ISL_2895665, EPI_ISL_3274100 | INSPI-CRN DE INFLUENZA Y OTROS VIRUS RESPIRATORIOS | NIC-INSPI | Alfredo Bruno; Daniel Ramos; Domenica de Mora.; Jimmy Garcés; Johanna Laines; Lizbeth Patiño; Manuel Gonzalez; Maria Angelica Becerra; Maritza Olmedo; Mayra Wilca; Michelle Páez |
| EPI_ISL_5094814 | Illinois Department of Public Health - Springfield Lab | Illinois Department of Public Health - Springfield Lab | Bryan Sim; Gordon McCall |
| EPI_ISL_3540049, EPI_ISL_3540053, EPI_ISL_3540064, EPI_ISL_3540065, EPI_ISL_3540081, EPI_ISL_3924333, EPI_ISL_3924338 | | | |
| see above | Indira Gandhi Memorial Hospital | Indiar Gandhi Memorial Hospital | D. Fathmath Nazla Rafeeq; Mr. Ibrahim Nishan Ahmed; Ms. Aishath Shuhudha; Ms. Aminath Nazfa; Ms. Fathimath Zimna |
| EPI_ISL_5051616, EPI_ISL_5061889, EPI_ISL_5061906 | Indira Gandhi Memorial Hospital | Indira Gandhi Memorial Hospital | D. Fathmath Nazla Rafeeq; Dr. Ibrahim Afzal; Mr. Ibrahim Nishan Ahmed; Ms. Aishath Shuhudha; Ms. Aminath Shazleena Abdul Rahman; Ms. Fathimath Zimna |
| EPI_ISL_4904872, EPI_ISL_5093331, EPI_ISL_5208456, EPI_ISL_5208653, EPI_ISL_5209287, EPI_ISL_5221089, EPI_ISL_5318049, EPI_ISL_5380596 | | | |
| see above | Infinity Biologix | Centers for Disease Control and Prevention Division of Viral Diseases, Pathogen Discovery | Benjamin Rambo-Martin; Chirayu Goswami; Christian Bixby; Christopher Gulvick; Clinton Paden; Dakota Howard; Dhwani Batra; Duncan MacCannell; Erisa Sula; Jason Caravas; Jonathan Schultz; Kristine Lacek; Matthew Schmerer; Peter Cook; Robin Grimwood; Russ Hager; Scott Sammons; Shatavia Morrison; Tymeckia Kendall; Victoria Caban Figueroa; Yihe Wang; Yvette Unoarumhi |
| EPI_ISL_3915027, EPI_ISL_3915056, EPI_ISL_3915061, EPI_ISL_3915113, EPI_ISL_3915124, EPI_ISL_3915134 | Institut National D'hygiene, Lome, Togo | Noguchi Memorial Institute for Medical Research, University of Ghana, Legon, Ghana | Afiwa W. Halatoko; Ameyo Dorkenoo; Anoumou Dagnran; Djimabi Salah; Hamadi Assane; Hilda Opoku Frempong; Issaka Maman; Joseph H.K. Bonney; Joyce Appiah-Kubi; Keren O. Attiku; Komlan Kossi; Lallepak Lamboni; Mounerou Salou; Peace O. Uche; Quaneeta Mohktar; Sena Awunyo; Seyram B. Agbenyo; Yao Layibo; Yawo A. Sadji; Zoukaneirii Issa; and Bright Adu |
| EPI_ISL_3453321, EPI_ISL_4270843, EPI_ISL_4271528, EPI_ISL_5212878, EPI_ISL_5330003, EPI_ISL_5333354, EPI_ISL_5333561, EPI_ISL_5333562 | | | |
| see above | Institute of Microbiology and Immunology, Faculty of Medicine, University of Ljubljana | Institute of Microbiology and Immunology, Faculty of Medicine, University of Ljubljana | Alen Suljič; Andraž Celar; Domen Lazar; Dominika Šturm; Doroteja Vlaj; Mario Poljak; Matic Brvar; Miša Korva; Patricija Pozvek; Samo Zakotnik; Tatjana Avšič – Županc; Tina Gabrovšek; Tina Živič; Tomaž Mark Zorec; Špela Pleh |
| EPI_ISL_3505616, EPI_ISL_4511383, EPI_ISL_4511384, EPI_ISL_4653332 | Institute of Microbiology, Universidad San Francisco de Quito | Institute of Microbiology, Universidad San Francisco de Quito | Belén Prado-Vivar; Bernardo Gutiérrez; Erika B. Muñoz; Fernanda Zurita; Francisco Cordova; Gabriel Trueba; Juan José Guadalupe; Killen Briones-Claudette Verónica Barragán; Killen Briones-Zamora; Manuel Jaramillo; Mateo Carvajal; Michelle Grunauer; Monica Becerra-Wong; Ninfa Henriquez; Patricio Rojas-Silva; Paúl Cárdenas; Rafael Parra; Ramiro Echeverría Tapia; Sully Márquez; Verónica Barragán |
| EPI_ISL_4239896, EPI_ISL_4239946, EPI_ISL_4239950 | Institute of Public Health | Institute of Public Health | Branka Culibrk; Dijana Vukajlovic; Milica Celic; Pava Dimitrijevic; Zeljka Sumic |
| EPI_ISL_3556742 | Institute of Virology, Biomedical Research Center of the Slovak Academy of Sciences, Bratislava | Faculty of Mathematics, Physics and Informatics, Comenius University, Bratislava | Boris Klempa; Brona Brejova; Jozef Nosek; Juraj Kopacek; Kristina Borsova; Lubomira Lukacikova; Martina Lickova; Martina Nebohacova; Monika Slavikova; Sabina Fumacova Havlikova; Tomas Vinar; Veronika Vanova; Viktoria Cabanova |
| EPI_ISL_4468843, EPI_ISL_4468856, EPI_ISL_4468859, EPI_ISL_4468867 | Institute of Virology, Vaccines and Sera "Torlak" | Institute of microbiology and Immunology, Faculty of Medicine, University of Belgrade | Banko A.; Cupic M.; Jankovic, M.; Jovanovic, T.; Knezevic, A.; Lazarevic I.; Milicevic, O.; Miljanovic D.; Sekler, M.; Tesovic, B.; Vidanovic, D. |
| EPI_ISL_3859848, EPI_ISL_4575076 | Instituto Nacional de Investigação em Saúde | CERI, Centre for Epidemic Response and Innvoation, Stellenbosch University and KRISP, KZN Research Innovation and Sequencing Platform, UKZN. | Afonso P; David K; Emmanuel SJ; Freitas RH; Giandhari J; Inglês L; Lutucuta S; Miranda J; Morais J; Mufinda M; Naidoo Y; Neto Z; Paulo A Carralero RR Paixão JP; Pereira A; Pillay S; Tegally H; Wilkinson E; de Oliveira T |
| EPI_ISL_3548347, EPI_ISL_4232454 | Instituto Nacional de Medicina Genomica | Instituto Nacional de Medicina Genomica | Cedro-Tanda A; Cruz-Islas Jazmin; Escobar-Arrazola MA; Garnica-Lopez Dora; Herrera-Montalvo LA.; Hidalgo-Miranda A; Mendoza-Vargas A; Munguia-Garza P; Ramirez-Vega O; Rangel-DeLeon D; Reyes-Grajeda JP; Yair Alfaro-Mora |
| EPI_ISL_3663587, EPI_ISL_3663595, EPI_ISL_5425640, EPI_ISL_5425680, EPI_ISL_5425687, EPI_ISL_5425738, EPI_ISL_5425740, EPI_ISL_5425741, EPI_ISL_5425755, EPI_ISL_5425778, EPI_ISL_5425780 | | | |
| see above | Instituto Nacional de Saude (INS), Mozambique | CERI, Centre for Epidemic Response and Innvoation, Stellenbosch University and KRISP, KZN Research Innovation and Sequencing Platform, UKZN. | Emmanuel S; Giandhari J; Giandhari Jennifer; Nadia Sitoe; Naidoo Yeshnee; Nalia Ismael; Nedio Mabunda; Paulo Arnaldo; Pillay S; Pillay Sureshnee; San James; Tegally H; Tegally Houriiyah; Tshabuila Derek; Tshiabuila Derek; Wilkinson E; Wilkinson Eduan; Yajna Ramphal; de Oliveira T; de Oliveira Tulio |
| EPI_ISL_2620888, EPI_ISL_2887857 | Iressef Genomics lab | IRESSEF | Abdou PADANE; Ambroise AHOUIDI; Aminata DIA; Aminata MBOUP; Astou Gaye GAYE; Barada CISSE; Birahim Piere NDIAYE; Diabou Diagne; Gora LO; Khadim GUEYE; Moustapha MBOW; Nafisatou LEYE; Ndeye Coumba Toure KANE; Papa Alassane DIAW; Samba Ndiour; Seni Ndiaye; Souleymane MBOUP; Yacine DIA |
| EPI_ISL_3087588, EPI_ISL_3087599 | Jaber Quarantine Station | Virology Lab, Jaber Al Ahmad Hospital | Adil Afridi; Dr. Ebaa Al-Awadhi; Dr. Zahrah Buhamad; Haroon Masih; Khubaid-ur-Rehman |
| EPI_ISL_3866905 | Jarallah German Specialized Clinic | Virology Lab, Jaber Al Ahmad Hospital | Dr. Ebaa Al-Awadhi; Dr. Zahrah Buhamad; Estabraq Kathim; Haroon Masih; Khubaid-ur-Rehman |
| EPI_ISL_5324028, EPI_ISL_5324072, EPI_ISL_5324090 | Jordan Royal Medical Services /Aqaba Molecular Lab | Jordan Royal Medical Services Genomics Core Lab | Abdullah Almuhasen; Alanood Alhabashnah; Ali Alhuniti; Dr. Rame Hamdi khasawneh; Mohammad Barmawi; Osama Alshdaifat |
| EPI_ISL_5137568 | Jordan Royal Medical Services Aqaba Molecular Lab | Jordan Royal Medical Services Genomics Core Lab | Abdullah Almuhasen; Alanood Alhabashnah; Ali Alhuniti; Dr. Rame Hamdi khasawneh; Mohammad Barmawi; Osama Alshdaifat |
| EPI_ISL_4849563, EPI_ISL_5067986 | Jordan Royal Medical Services Mobile Biological Lab | Jordan Royal Medical Services Genomics Core Lab | Abdullah Almuhasen; Alanood Alhabashnah; Ali Alhuniti; Dr. Rame Hamdi khasawneh; Dr. Rame Hamdi khasawneh; Mohammad Barmawi; Osama Alshdaifat |
| EPI_ISL_3049528, EPI_ISL_3049530, EPI_ISL_3049683, EPI_ISL_3049838, EPI_ISL_3049856, EPI_ISL_3049865, EPI_ISL_4196981, EPI_ISL_4196988, EPI_ISL_4196994, EPI_ISL_4197000, EPI_ISL_4197008, EPI_ISL_4197034, EPI_ISL_4197090 | | | |
| see above | KEMRI-Wellcome Trust Research Programme,Kilifi | KEMRI-Wellcome Trust Research Programme,Kilifi | Agoti C.; Githinji G.; Lambisia A.; Matoke D.; Mburu M.W.; Mohamed K.S.; Morobe J.; Ndwiga L.; Ochola I; Ong'era M.Edidah; Thiongo K.; de Laurent Z. |
| EPI_ISL_4174900 | KSL Diagnostics | University at Buffalo Genomics and Bioinformatics Core | Alyssa Pohlman; Amanda Boccolucci; Brandon Marzullo; Donald Yergeau; Jennifer Surtees; Jonathan Bard; Natalie Lamb; Norma Nowak |
| EPI_ISL_4365265, EPI_ISL_4651117, EPI_ISL_5368519, EPI_ISL_5385470, EPI_ISL_5391251, EPI_ISL_5429063 | KU Leuven, Rega Institute, Clinical and Epidemiological Virology | KU Leuven, Rega Institute, Clinical and Epidemiological Virology | Bert Vanmechelen; Joan Marti-Carerras; Piet Maes; Tony Wawina-Bokalanga |
| EPI_ISL_2983507, EPI_ISL_4004765, EPI_ISL_5424027 | Kansas Health and Environmental Lab | Kansas Health and Environmental Lab | Amanda Bradley; Ben Olsen; Carrie Welch; Gary Buruss; Jonathan Barnell; Katherine Wiggins; Meg Wise; Mike Grose; Victor Anderson; and Phil Adam |
| EPI_ISL_4563913 | Karachay-Cherkess Republican Clinical Hospital | WHO National Influenza Centre Russian Federation | Andrey Komissarov; Artem Fadeev; Daria Danilenko; Dmitry Lioznov; Elena Nabieva; Georgii Bazykin; Kirill Varchenko; Ksenia Safina; Kseniya Komissarova; Maria Pisareva; Mikhail Bakaev; Nikita Yolshin; Oula Mansour; Tamila Musaeva; Veronika Eder |
| EPI_ISL_5050057 | Karolinska University Hospital Solna | Karolinska University Hospital | Annelie Bjerkner; Isak Sylvin; Jan Albert; Karolina Ininbergs; Lina Guerra Blomqvist; Lynda Eneh; Martin Ekman; Martina Wahlund; Robert Dyrdak; Sandra Broddesson; Tanja Normark; Tobias Allander; Valtteri Wirta; Zhibing Yun |
| EPI_ISL_5332508 | Klang Hospital | Division of Genomic Medicine and | Archawin Rojanawiwat; Jirapha Pakdee; Natthakul Bunneang; Nuanjun Wichukchinda; Penpitcha Thawong; Pilailuk Akkapaiboon Okada; Pundharika Piboonsiri; Surakameth Mahasirimongkol; Waritta Sawaengdee |

| | | | |
|---|---|---|---|
| | | Innovation support,Department of Medical Sciences, Ministry of Public Health, Thailand | |
| EPI_ISL_3631157, EPI_ISL_3731194, EPI_ISL_5195816 | Klinika za infektivne bolesti "Dr. Fran Mihaljević" | Hrvatski zavod za javno zdravstvo | Irena Tabain; Ivana Ferenčak |
| EPI_ISL_5196177 | Klinički bolnički centar Rijeka | Hrvatski zavod za javno zdravstvo | Irena Tabain; Ivana Ferenčak |
| EPI_ISL_2938095 | LABORATORIO DR PAULO EMILIO DALESSANDRO PINDAMONHANGABA | Instituto Butantan | Antonio Jorge Martins; Claudia Renata dos Santos Barros; David Schlesinger; Debora Botequio Moretti; Dimas Tadeu Covas; Elaine Cristina Marqueze; Elaine Vieira Santos; Evandra Strazza Rodrigues; Heidge Fukumasu; Jayme Augusto de Souza-Neto; José Salvatore Leister Patané; Luiz Alcantara; Luiz Lehmann Coutinho; Maria Carolina Elias; Maurício Lacerda Nogueira; Rafael dos Santos Bezerra; Raul Machado Neto; Rejane Maria Tommasini Grotto; Ricardo Haddad; Sandra Coccuzzo Sampaio Vessoni; Simone Kashima; Svetoslav Nanev Slavov; Vincent Louis Viala |
| EPI_ISL_4107327 | LAC UCoimbra | Instituto Nacional de Saude (INSA) | Borges et al |
| EPI_ISL_3912402 | LACLIM | Analytical Competence Molecular Epidemiology Lab/ACME, Oswaldo Cruz Foundation, Ceara (FIOCRUZ CE) | Cleber Furtado Aksenen; Fabio Miyajima; Fernando Braga Stehling; Francisco Eder de Moura Lopes; Jamille Maria Mendes Bezerra; Joaquim Cesar do Nascimento Sousa Junior; Pedro Miguel Carneiro Jeronimo; Suzana Porto Almeida & Lucas Delerino on behalf of COVID-19 FIOCRUZ Genomic Network; Thais Ferreira de Oliveira; Thais de Oliveira Costa; Ticiane Cavalcante de Souza; Veridiana Pessoa Miyajima |
| EPI_ISL_4605034 | LAM CERBALLIANCE | CNR Virus des Infections Respiratoires - France SUD | Antonin Bal; Bruno Lina; Gregory Destras; Gwendolyne Burfin; Hadrien Regue; Laurence Josset; Martine Valette; Quentin Semanas |
| EPI_ISL_2965582, EPI_ISL_3385133, EPI_ISL_3385139, EPI_ISL_4233778, EPI_ISL_4926911, EPI_ISL_4926915, EPI_ISL_5438914 | | | |
| see above | LATE - Laboratório de Técnicas Especiais - Hospital Israelita Albert Einstein | LATE - Laboratório de Técnicas Especiais - Hospital Israelita Albert Einstein | Alexandre Hideaki Takara; Ana Paula Moreira Salles; Anelisie da Silva Santos; Deyvid Amgarten; Erick Gustavo Dorlass; Fernanda de Mello Malta; João Renato Rebello Pinho; Marcio Anunciacao Menezes; Pedro Henrique Sebe Rodrigues; Raquel Riyuzo |
| EPI_ISL_4877368 | LBV Le DANTEC | LBV Le DANTEC | Adjiratou Aissatou BA; Aminata Sileymane Thiam; Anna julienne Selbe NDiaye; Assane Dieng; Awa Ba-Diallo; Dianke Samaté; Gora Lo; Halimatou Diop Ndiaye; Khadim GUEYE; Makhtar Camara; Mbengué Fall; Moustapha Sakho; Oumy DIOP; Pascaline Manga; Pauline Yacine Sene; Sada Diallo; Samba NDOUR; Serigne Saliou Niane; ousseynou Gueye |
| EPI_ISL_2937902, EPI_ISL_3265480, EPI_ISL_4298943 | LESP Aguascalientes | Instituto de Diagnostico y Referencia Epidemiologicos (INDRE) | Abril Rodriguez-Maldonado; Ariadna Medina-Benitez; Claudia Wong-Arambula; Ernesto Ramirez-Gonzalez.; Gisela Barrera-Badillo; Irma Lopez-Martinez; Joaquin Quiroz-Mercado; Lucia Hernandez-Rivas; Maribel Gonzalez-Villa; Natividad Cruz-Ortiz; Sergio Rangel-Guerrero; Tatiana Nunez-Garcia; Vanessa Rivero-Arredondo |
| EPI_ISL_3265554 | LESP Baja California | Instituto de Diagnostico y Referencia Epidemiologicos (INDRE) | Abril Rodriguez-Maldonado; Ariadna Medina-Benitez; Claudia Wong-Arambula; Ernesto Ramirez-Gonzalez.; Gisela Barrera-Badillo; Irma Lopez-Martinez; Joaquin Quiroz-Mercado; Lucia Hernandez-Rivas; Maribel Gonzalez-Villa; Natividad Cruz-Ortiz; Sergio Rangel-Guerrero; Tatiana Nunez-Garcia; Vanessa Rivero-Arredondo |
| EPI_ISL_3033411, EPI_ISL_3460220, EPI_ISL_4298965 | LESP Baja California Sur | Instituto de Diagnostico y Referencia Epidemiologicos (INDRE) | Abril Rodriguez-Maldonado; Ariadna Medina-Benitez; Claudia Wong-Arambula; Ernesto Ramirez-Gonzalez.; Gisela Barrera-Badillo; Irma Lopez-Martinez; Joaquin Quiroz-Mercado; Lucia Hernandez-Rivas; Maribel Gonzalez-Villa; Natividad Cruz-Ortiz; Sergio Rangel-Guerrero; Tatiana Nunez-Garcia; Vanessa Rivero-Arredondo |
| EPI_ISL_3265562, EPI_ISL_4602969 | LESP Campeche | Instituto de Diagnostico y Referencia Epidemiologicos (INDRE) | Abril Rodriguez-Maldonado; Ariadna Medina-Benitez; Claudia Wong-Arambula; Ernesto Ramirez-Gonzalez.; Fernando Gonzalez-Dominguez; Gisela Barrera-Badillo; Irma Lopez-Martinez; Joaquin Quiroz-Mercado; Lucia Hernandez-Rivas; Maribel Gonzalez-Villa; Natividad Cruz-Ortiz; Sergio Rangel-Guerrero; Tatiana Nunez-Garcia; Vanessa Rivero-Arredondo |
| EPI_ISL_3769214, EPI_ISL_4298749 | LESP Coahuila | Instituto de Diagnostico y Referencia Epidemiologicos (INDRE) | Abril Rodriguez-Maldonado; Ariadna Medina-Benitez; Claudia Wong-Arambula; Ernesto Ramirez-Gonzalez.; Gisela Barrera-Badillo; Irma Lopez-Martinez; Joaquin Quiroz-Mercado; Lucia Hernandez-Rivas; Maribel Gonzalez-Villa; Natividad Cruz-Ortiz; Sergio Rangel-Guerrero; Tatiana Nunez-Garcia; Vanessa Rivero-Arredondo |
| EPI_ISL_4602925, EPI_ISL_4918659 | LESP Durango | Instituto de Diagnostico y Referencia Epidemiologicos (INDRE) | Abril Rodriguez-Maldonado; Ariadna Medina-Benitez; Claudia Wong-Arambula; Ernesto Ramirez-Gonzalez.; Fernando Gonzalez-Dominguez; Gisela Barrera-Badillo; Irma Lopez-Martinez; Joaquin Quiroz-Mercado; Lucia Hernandez-Rivas; Maribel Gonzalez-Villa; Natividad Cruz-Ortiz; Sergio Rangel-Guerrero; Tatiana Nunez-Garcia; Vanessa Rivero-Arredondo |
| EPI_ISL_4298713, EPI_ISL_4918479 | LESP Guanajuato | Instituto de Diagnostico y Referencia Epidemiologicos (INDRE) | Abril Rodriguez-Maldonado; Ariadna Medina-Benitez; Claudia Wong-Arambula; Ernesto Ramirez-Gonzalez.; Fernando Gonzalez-Dominguez; Gisela Barrera-Badillo; Irma Lopez-Martinez; Joaquin Quiroz-Mercado; Lucia Hernandez-Rivas; Maribel Gonzalez-Villa; Natividad Cruz-Ortiz; Sergio Rangel-Guerrero; Tatiana Nunez-Garcia; Vanessa Rivero-Arredondo |
| EPI_ISL_3460090 | LESP Guerrero | Instituto de Diagnostico y Referencia Epidemiologicos (INDRE) | Abril Rodriguez-Maldonado; Ariadna Medina-Benitez; Claudia Wong-Arambula; Ernesto Ramirez-Gonzalez.; Gisela Barrera-Badillo; Irma Lopez-Martinez; Joaquin Quiroz-Mercado; Lucia Hernandez-Rivas; Maribel Gonzalez-Villa; Natividad Cruz-Ortiz; Sergio Rangel-Guerrero; Tatiana Nunez-Garcia; Vanessa Rivero-Arredondo |
| EPI_ISL_3769064 | LESP Jalisco | Instituto de Diagnostico y Referencia Epidemiologicos (INDRE) | Abril Rodriguez-Maldonado; Ariadna Medina-Benitez; Claudia Wong-Arambula; Ernesto Ramirez-Gonzalez.; Gisela Barrera-Badillo; Irma Lopez-Martinez; Joaquin Quiroz-Mercado; Lucia Hernandez-Rivas; Maribel Gonzalez-Villa; Natividad Cruz-Ortiz; Sergio Rangel-Guerrero; Tatiana Nunez-Garcia; Vanessa Rivero-Arredondo |
| EPI_ISL_5428198 | LESP Michoacan | Instituto de Diagnostico y Referencia Epidemiologicos (INDRE) | Abril Rodriguez-Maldonado; Ariadna Medina-Benitez; Claudia Wong-Arambula; Ernesto Ramirez-Gonzalez.; Fernando Gonzalez-Dominguez; Gisela Barrera-Badillo; Irma Lopez-Martinez; Joaquin Quiroz-Mercado; Lucia Hernandez-Rivas; Maribel Gonzalez-Villa; Natividad Cruz-Ortiz; Sergio Rangel-Guerrero; Tatiana Nunez-Garcia; Vanessa Rivero-Arredondo |
| EPI_ISL_4199384, EPI_ISL_4918018 | LESP Morelos | Instituto de Diagnostico y Referencia Epidemiologicos (INDRE) | Abril Rodriguez-Maldonado; Ariadna Medina-Benitez; Claudia Wong-Arambula; Ernesto Ramirez-Gonzalez.; Fernando Gonzalez-Dominguez; Gisela Barrera-Badillo; Irma Lopez-Martinez; Joaquin Quiroz-Mercado; Lucia Hernandez-Rivas; Maribel Gonzalez-Villa; Natividad Cruz-Ortiz; Sergio Rangel-Guerrero; Tatiana Nunez-Garcia; Vanessa Rivero-Arredondo |
| EPI_ISL_3768928 | LESP Queretaro | Instituto de Diagnostico y Referencia Epidemiologicos (INDRE) | Abril Rodriguez-Maldonado; Ariadna Medina-Benitez; Claudia Wong-Arambula; Ernesto Ramirez-Gonzalez.; Gisela Barrera-Badillo; Irma Lopez-Martinez; Joaquin Quiroz-Mercado; Lucia Hernandez-Rivas; Maribel Gonzalez-Villa; Natividad Cruz-Ortiz; Sergio Rangel-Guerrero; Tatiana Nunez-Garcia; Vanessa Rivero-Arredondo |
| EPI_ISL_5428367 | LESP Sonora | Instituto de Diagnostico y Referencia Epidemiologicos (INDRE) | Abril Rodriguez-Maldonado; Ariadna Medina-Benitez; Claudia Wong-Arambula; Ernesto Ramirez-Gonzalez.; Fernando Gonzalez-Dominguez; Gisela Barrera-Badillo; Irma Lopez-Martinez; Joaquin Quiroz-Mercado; Lucia Hernandez-Rivas; Maribel Gonzalez-Villa; Natividad Cruz-Ortiz; Sergio Rangel-Guerrero; Tatiana Nunez-Garcia; Vanessa Rivero-Arredondo |
| EPI_ISL_3769231 | LESP Tamaulipas | Instituto de Diagnostico y Referencia Epidemiologicos (INDRE) | Abril Rodriguez-Maldonado; Ariadna Medina-Benitez; Claudia Wong-Arambula; Ernesto Ramirez-Gonzalez.; Gisela Barrera-Badillo; Irma Lopez-Martinez; Joaquin Quiroz-Mercado; Lucia Hernandez-Rivas; Maribel Gonzalez-Villa; Natividad Cruz-Ortiz; Sergio Rangel-Guerrero; Tatiana Nunez-Garcia; Vanessa Rivero-Arredondo |
| EPI_ISL_5169009, EPI_ISL_5169014, EPI_ISL_5169041, EPI_ISL_5169046, EPI_ISL_5169055, EPI_ISL_5169100 | LNS | Inciensa, Instituto Costarricense de Investigación y Enseñanza en Nutrición y Salud | Claudia Estrada; César Conde; Laboratorio de Genómica INCIENSA; Selene González |
| EPI_ISL_2406490, EPI_ISL_2811949, EPI_ISL_2811957, EPI_ISL_2964928, EPI_ISL_2964935, EPI_ISL_3164078, EPI_ISL_3477084, EPI_ISL_3477096, EPI_ISL_3506212 | | | |
| see above | LabPLUS | Institute of Environmental Science and Research (ESR) | Anja Werno; Antje van der Linden; Arlo Upton; Chris Mansell; David Hammer; Dragana Drinkovic; Erasmus Smit; Gary McAuliffe; Hana Sofia Andersson; Hermes Perez; James Ussher; Jill Sherwood; Jing Wang; Joep de Ligt; Josh Freeman; Julia Howard; Juliet Elvy; Lauren Jelly; Mary DeAlmeida; Matt Blakiston; Matt Storey; Matthew Rogers; Max Bloomfield; Michael Addidle; Michelle Balm; Muhammad Faisal; Nikki Freed; Olin Silander; Olivia Stroeven; Rachel Boyle; Sally Roberts; SallyAnn Harbison; Sarah Jefferies; Sharmini Muttaiyah; Susan Morpeth; Susan Taylor; Timothy Blackmore; Vani Sathyendran; Veronica Playle; Virginia Hope; Xiaoyun Ren |
| EPI_ISL_5032979, EPI_ISL_5348084 | LabTests | Institute of Environmental Science and Research (ESR) | Anja Werno; Antje van der Linden; Arlo Upton; Chris Mansell; Clare Gebbie; David Hammer; Dhanisha Patel; Dragana Drinkovic; Erasmus Smit; Gary McAuliffe; Hana Sofia Andersson; Hermes Perez; James Ussher; Jill Sherwood; Jing Wang; Joep de Ligt; Josh Freeman; Julia Howard; Juliet Elvy; Lauren Jelly; Mary DeAlmeida; Matt Blakiston; Matt Storey; Matthew Rogers; Max Bloomfield; Michael Addidle; Michelle Balm; Muhammad Faisal; Nikki Freed; Olin Silander; Olivia Stroeven; Rachel Boyle; Sally Roberts; SallyAnn Harbison; Sarah Cockerton; Sarah Jefferies; Sharmini Muttaiyah; Susan Morpeth; Susan Taylor; Timothy Blackmore; Vani Sathyendran; Veronica Playle; Virginia Hope; Xiaoyun Ren |
| EPI_ISL_2923151, EPI_ISL_2923153, EPI_ISL_2923155, EPI_ISL_2923158, EPI_ISL_3031789, EPI_ISL_3690199, EPI_ISL_3999634, EPI_ISL_4171692, EPI_ISL_4171693, EPI_ISL_4171696, EPI_ISL_4171700, EPI_ISL_4171703, EPI_ISL_4193971, EPI_ISL_4411626, EPI_ISL_4461667, EPI_ISL_4740991, EPI_ISL_4740992, EPI_ISL_5332206 | | | |
| see above | Labo Analyses Med | National Reference Center for Viruses of Respiratory Infections, Institut Pasteur, Paris | Angela Brisebarre; Benedicte Baccouch; Camille Capel; Christophe Malabat; Corinne Maufrais; DURIVAULT Jérôme; Emmanuelle Permal; Etienne Simon-Lorière; Frédéric Lemoine; Henri Duvert; Hub Bioinformatique Biostatistiques; Hub de Bioinformatique et Biostatistique; Julien Fumey; Louise Lefrançois; Marion Barbet; Maud Vanpeene; Méline Bizard; Olivier Dejoux; Pascal Maillet; Patricia Tamby; Slim El Khiari; Slim El-Khiari; Sylvie Behillil; Sylvie Van der Werf; TAMBY Patricia; Vincent Enouf |
| EPI_ISL_4884663, EPI_ISL_5460163 | Labor Berlin Charité Vivantes GmbH / Institut für Virologie | Charité Universitätsmedizin Berlin, Institut für Virologie/Labor Berlin | Barbara Mühlemann; Christian Drosten; Christine Stephan; Peter Menzel; Rolf Schwarzer; Terry Jones; Victor M Corman |
| EPI_ISL_4939938 | Labor Dr. Wisplinghoff - Köln | Robert Koch Institute | |
| EPI_ISL_3453048, EPI_ISL_3453059 | Laboratoire MAYMAT | Department of Virology, Henri Mondor University Hospital, Assistance Publique Hôpitaux de Paris, Université Paris-Est Créteil, INSERM U955 | Alexandre Soulier; Christophe Rodriguez; Elisabeth Trawinski; Guillaume Gricourt; Jean-Michel Pawlotsky; Melissa N'Debi; Slim Fourati; Vanessa Demontant |
| EPI_ISL_3459210 | Laboratoire de santé publique du Québec | Laboratoire de santé publique du Québec | Guillaume Bourque; Ioannis Ragoussis; Jesse Shapiro; Mark Lathrop and Michel Roger on behalf of the CoVSeQ research group; Sandrine Moreira |
| EPI_ISL_4566907, EPI_ISL_4566912, EPI_ISL_4566971, EPI_ISL_4566980, EPI_ISL_4567008 | Laboratoire des Fièvres Hémorragiques Virales du Benin | Institut für Virologie - Institute of Virology - Charite | Andres Moreira-Soto; Anges Yadouleton; Anna-Lena Sander; Ben Wulf; Benjamin Hounkpatin and Jan Felix Drexler; Carine Tchibozo; Christian Drosten; Clement G. Kakai; Dossou Ange; Edmilson F. de Oliveira Filho; Fattah Al Onifade; Gildas Hounkanrin; Keke K. René; Mamoudou Harouna Djingarey; Melchior A. Joël Aïssi; Michael Nagel; Petas Akogbeto; Praise Adewumi; Ramalia Chabi Nari; Raoul Saizonou; Rodrigue K. Kohoun; Sonia V. Bedié; Sourakatou Salifou |
| EPI_ISL_5395068, EPI_ISL_5395163, EPI_ISL_5395219 | Laboratoires d'analyses medicales - Ketterthill | Laboratoire national de sante, Microbiology, Microbial Genomics Platform | Anke Wienecke-Baldacchino; Caroline Scheiber; Catherine Ragimbeau; Elodie Solarino; Fatu Djabi; Jessica Tapp; Lise Pignon; Raoul Salmon; Serge Vedy; Tamir Abdelrahman; Virginie Jover |
| EPI_ISL_3384886, EPI_ISL_4251904, EPI_ISL_4251906, EPI_ISL_4949311, EPI_ISL_4949312 | Laboratori d'anàlisis clíniques, Hospital Nostra Senyora de Meritxell | LBM de CHU de Toulouse, Hôpitaux de Toulouse | Bartolome C et al.; Lobaco C; M; Rendon M; Rendon et Al |
| EPI_ISL_3832329, EPI_ISL_3832332, EPI_ISL_3832335, EPI_ISL_3832336, EPI_ISL_3832337, EPI_ISL_3832338, EPI_ISL_3832339, EPI_ISL_3832342, EPI_ISL_4170345 | | | |
| see above | Laboratorio Central de Saude | Laboratory of Respiratory Viruses and | Alice Sampaio Rocha; Ana Carolina Mendonca; Anna Carolina Paixao; Elisa Cavalcante Pereira; Fernando Motta; Luciana Appolinario; Marilda Siqueira on behalf of the Fiocruz COVID-19 Genomic Surveillance Network; Paola Resende; Renata Serrano Lopes; Rubens Pasa; Taina Venas |

| | | | |
|---|---|---|---|
| | Publica do Estado de Minas Gerais (LACEN/MG) | Measles, Oswaldo Cruz Institute, FIOCRUZ | |
| EPI_ISL_5116076 | Laboratorio CQRC | CQRC_QUALITY CONTROL CHEMICAL BIOLOGICAL RISK_AOOR Villa Sofia Cervello Palermo | Brunacci G.; Contino F.; Di Gaudio F. |
| EPI_ISL_2942869, EPI_ISL_4549964, EPI_ISL_4550103, EPI_ISL_4550128, EPI_ISL_4550144 | Laboratorio Central de Epidemiologia (LCE) | Unidad de Genomica Avanzada | ; Alejandra Garcia-Gasca; Alejandra Hernandez-Teran; Alejandro Sanchez-Flores; Alfredo Herrera-Estrella; Alicia Ocaña-Mondragon; Andreu Comas-Garcia; Angel Gustavo Salas-Lais; Antonio Loza Roman; Bernardo Martinez-Miguel; Blanca Taboada; Brenda Irasema Maldonado-Meza; Bruno Gomez-Gil; Carla Ivon Herrera-Najera; Carlos F. Arias; Celia Boukadida; Celida Duque Molina; Celida Martinez- Rodriguez; Clara Esperanza Santacruz-Tinoco; Concepcion Grajales-Muñiz; Consorcio Mexicano de Vigilancia Genomica (CoViGen-Mex). Authors (in alphabetical order): Julio Elias Alvarado-Yaah; Cristobal Chaidez-Quiroz; Daniel Fregoso-Rueda; Daniel Lira Morales; Eduardo Becerril-Vargas; Fernando Fontove-Herrera; Fidencio Mejia-Nepomuceno; Francisco Pulido; Gloria Elena Espinosa-Ayala; Gloria Maria Molina-Salinas; Gloria Vazquez; Hector Esteban Paz-Juarez; Hector Montoya-Fuentes; Helen Haydee Fernanda Ramirez-Plascencia; Irvin Gonzalez-Lopez; Jean Pierre Gonzalez; Jesus Hernandez; Joel Armando Vazquez-Perez.; Jorge Salas-Hernandez; Jose Antonio Enciso-Moreno; Jose Arturo Martinez-Orozco; Jose Esteban Muñoz-Medina; Jose de Jesus Nuñez-Contreras; Juan Bautista Chale-Dzul; Julissa Enciso-Ibarra; Luis Alberto Ochoa-Carrera; Margarita Matias-Florentino; Maria Guadalupe Santiago-Mauricio; Maria Guadalupe de Jesus Mireles-Rivera; Mario Mujica-Sanchez; Marissa Perez-Garcia; Nelly Selem-Mojica; Pavel Isa; Ricardo Ciria Merce; Ricardo Grande; Rosa Maria Gutierrez Rios; Santiago avila-Rios; Selene Zarate; Susana Lopez; Veronica Mata-Haro; Victor Eduardo Garcia-Arias; Victor Hugo Borja-Aburto |
| EPI_ISL_3805527, EPI_ISL_3805606, EPI_ISL_3805633 | Laboratorio Central de Epidemiologia (LCE) | Unidad de Genomica Avanzada | ; Alejandra Garcia-Gasca; Alejandra Hernandez-Teran; Alejandro Sanchez-Flores; Alfredo Herrera-Estrella; Alicia Ocaña-Mondragon; Andreu Comas-Garcia; Angel Gustavo Salas-Lais; Antonio Loza Roman; Bernardo Martinez-Miguel; Blanca Taboada; Brenda Irasema Maldonado-Meza; Bruno Gomez-Gil; Carla Ivon Herrera-Najera; Carlos F. Arias; Celia Boukadida; Celida Duque Molina; Celida Martinez- Rodriguez; Clara Esperanza Santacruz-Tinoco; Concepcion Grajales-Muñiz; Consorcio Mexicano de Vigilancia Genomica (CoViGen-Mex). Authors (in alphabetical order): Julio Elias Alvarado-Yaah; Cristobal Chaidez-Quiroz; Daniel Fregoso-Rueda; Daniel Lira Morales; Eduardo Becerril-Vargas; Fernando Fontove-Herrera; Fidencio Mejia-Nepomuceno; Francisco Pulido; Gloria Elena Espinosa-Ayala; Gloria Maria Molina-Salinas; Gloria Vazquez; Hector Esteban Paz-Juarez; Hector Montoya-Fuentes; Helen Haydee Fernanda Ramirez-Plascencia; Irvin Gonzalez-Lopez; Jean Pierre Gonzalez; Jesus Hernandez; Joel Armando Vazquez-Perez.; Jorge Salas-Hernandez; Jose Antonio Enciso-Moreno; Jose Arturo Martinez-Orozco; Jose Esteban Muñoz-Medina; Jose de Jesus Nuñez-Contreras; Juan Bautista Chale-Dzul; Julissa Enciso-Ibarra; Luis Alberto Ochoa-Carrera; Margarita Matias-Florentino; Maria Guadalupe Santiago-Mauricio; Maria Guadalupe de Jesus Mireles-Rivera; Mario Mujica-Sanchez; Marissa Perez-Garcia; Nelly Selem-Mojica; Pavel Isa; Ricardo Ciria Merce; Ricardo Grande; Rosa Maria Gutierrez Rios; Santiago avila-Rios; Selene Zarate; Susana Lopez; Veronica Mata-Haro; Victor Eduardo Garcia-Arias; Victor Hugo Borja-Aburto |
| EPI_ISL_4299639, EPI_ISL_5193916, EPI_ISL_5194013, EPI_ISL_5194115, EPI_ISL_5194268 | Laboratorio Central de Epidemiología (LCE) | Centro de Investigación en Enfermedades Infecciosas (CIENI), Instituto Nacional de Enfermedades Respiratorias (INER) | Alejandra García-Gasca; Alejandra Hernández-Terán; Alejandro Sánchez-Flores; Alfredo Herrera-Estrella; Alicia Ocaña-Mondragón; Andreu Comas-García; Angel Gustavo Salas-Lais; Antonio Loza Román; Bernardo Martínez-Miguel; Blanca Taboada; Brenda Irasema Maldonado-Meza; Bruno Gómez-Gil; Carla Ivón Herrera-Najera; Carlos F. Arias; Celia Boukadida; Clara Esperanza Santacruz-Tinoco; Concepción Grajales-Muñiz; Consorcio Mexicano de Vigilancia Genómica (CoViGen-Mex). Authors (in alphabetical order): Julio Elias Alvarado-Yaah; Cristóbal Cháidez-Quiróz; Célida Duque Molina; Célida Martínez-Rodríguez; Daniel Fregoso-Rueda; Daniel Lira Morales; Eduardo Becerril-Vargas; Eduardo Rivera-Martínez; Fernando Fontove-Herrera; Fidencio Mejía-Nepomuceno; Francisco Pulido; Gabriel Chavira-Trujillo; Gloria Elena Espinosa-Ayala; Gloria María Molina-Salinas; Gloria Vazquez; Hector Montoya-Fuentes; Helen Haydee Fernanda Ramírez-Plascencia; Irvin González-López; Jean Pierre González; Jesús Hernández; Joel Armando Vázquez-Pérez.; Jorge Salas-Hernández; José Antonio Enciso-Moreno; José Arturo Martínez-Orozco; José Esteban Muñoz-Medina; José de Jesús Nuñez-Contreras; Juan Bautista Chale-Dzul; Julissa Enciso-Ibarra; Kathia Elizabeth Tapia-Diaz; Luis Alberto Ochoa-Carrera; Margarita Matías-Florentino; Mario Mújica-Sánchez; Marissa Perez-Garcia; María Eugenia Jiménez-Corona; María Guadalupe Santiago-Mauricio; María Guadalupe de Jesús Mireles-Rivera; Nelly Sélem-Mojica; Pavel Isa; Ricardo Ciria Merce; Ricardo Grande; Rosa María Gutiérrez Rios; Rosario Vazquez-Larios; Santiago Ávila-Ríos; Selene Zárate; Susana Lopez; Verónica Mata-Haro; Victor Eduardo García-Arias; Víctor Hugo Borja-Aburto |
| EPI_ISL_3347525, EPI_ISL_3347794, EPI_ISL_4006377, EPI_ISL_4878758, EPI_ISL_4878996 | Laboratorio Central de Epidemiología (LCE) | Instituto de Biotecnología de la UNAM | ; Alejandra García-Gasca; Alejandra Hernández-Terán; Alejandro Sánchez-Flores; Alfredo Herrera-Estrella; Alicia Ocaña-Mondragón; Andreu Comas-García; Angel Gustavo Salas-Lais; Antonio Loza Román; Bernardo Martínez-Miguel; Blanca Taboada; Brenda Irasema Maldonado-Meza; Bruno Gómez-Gil; Carla Ivón Herrera-Najera; Carlos F. Arias; Celia Boukadida; Clara Esperanza Santacruz-Tinoco; Concepción Grajales-Muñiz; Consorcio Mexicano de Vigilancia Genómica (CoViGen-Mex). Authors (in alphabetical order): Julio Elias Alvarado-Yaah; Cristóbal Cháidez-Quiróz; Célida Duque Molina; Célida Martínez-Rodríguez; Daniel Fregoso-Rueda; Daniel Lira Morales; Eduardo Becerril-Vargas; Fernando Fontove-Herrera; Fidencio Mejía-Nepomuceno; Francisco Pulido; Gloria Elena Espinosa-Ayala; Gloria María Molina-Salinas; Gloria Vazquez; Hector Esteban Paz-Juárez; Hector Montoya-Fuentes; Helen Haydee Fernanda Ramírez-Plascencia; Irvin González-López; Jean Pierre González; Jesús Hernández; Joel Armando Vázquez-Pérez).; Jorge Salas-Hernández; José Antonio Enciso-Moreno; José Arturo Martínez-Orozco; José Esteban Muñoz-Medina; José de Jesús Nuñez-Contreras; Juan Bautista Chale-Dzul; Julissa Enciso-Ibarra; Kathia Elizabeth Tapia-Diaz; Luis Alberto Ochoa-Carrera; Margarita Matías-Florentino; Mario Mújica-Sánchez; Marissa Perez-Garcia; María Guadalupe Santiago-Mauricio; María Guadalupe de Jesús Mireles-Rivera; Nelly Sélem-Mojica; Pavel Isa; Ricardo Ciria Merce; Ricardo Grande; Rosa María Gutiérrez Rios; Santiago Ávila-Ríos; Selene Zárate; Susana Lopez; Verónica Mata-Haro; Victor Eduardo García-Arias; Víctor Hugo Borja-Aburto |
| EPI_ISL_4259551, EPI_ISL_4259554, EPI_ISL_4259613, EPI_ISL_4259619, EPI_ISL_4303323, EPI_ISL_4303332, EPI_ISL_4303333, EPI_ISL_4303334 | | | |
| see above | Laboratorio Central de Salud Pública | Laboratorio Central de Salud Pública | Andrea Gómez de la Fuente; Cesar Cantero; Cynthia Vazquez; Juan Torales; Julio Barrios; Maria Liz Gamarra; Sandra Gonzalez; Shirley Villalba; Tania Alfonzo |
| EPI_ISL_4633825 | Laboratorio Central de Saude Publica do Estado da Paraiba (LACEN/PB) | Laboratory of Respiratory Viruses and Measles, Oswaldo Cruz Institute, FIOCRUZ | Alice Sampaio Rocha; Ana Carolina Mendonca; Anna Carolina Paixao; Dalane Loudal Florentino Teixeira; Elisa Cavalcante Pereira; Fernando Motta; Joao Felipe Bezerra; Luciana Appolinario; Marilda Siqueira on behalf of the Fiocruz COVID-19 Genomic Surveillance Network; Paola Resende; Renata Serrano Lopes; Taina Venas |
| EPI_ISL_3827987 | Laboratorio Central de Saude Publica do Estado do Espirito Santo (LACEN/ES) | Laboratory of Respiratory Viruses and Measles, Oswaldo Cruz Institute, FIOCRUZ | Alice Sampaio Rocha; Ana Carolina Mendonca; Anna Carolina Paixao; Elisa Cavalcante Pereira; Fernando Motta; Luciana Appolinario; Marilda Siqueira on behalf of the Fiocruz COVID-19 Genomic Surveillance Network; Paola Resende; Renata Serrano Lopes; Rodrigo Ribeiro Rodrigues; Taina Venas |
| EPI_ISL_3801934 | Laboratorio Central de Saude Publica do Estado do Rio Grande do Sul (LACEN-RS) | Laboratory of Respiratory Viruses and Measles, Oswaldo Cruz Institute, FIOCRUZ | Alice Sampaio Rocha; Ana Carolina Mendonca; Anna Carolina Paixao; Elisa Cavalcante Pereira; Fernando Motta; Luciana Appolinario; Marilda Siqueira on behalf of the Fiocruz COVID-19 Genomic Surveillance Network; Paola Resende; Renata Serrano Lopes; Richard Salvato; Taina Venas; Tatiana Schaffer Gregianini |
| EPI_ISL_2982743, EPI_ISL_4220323, EPI_ISL_4220325, EPI_ISL_4220326, EPI_ISL_4220327, EPI_ISL_4220328, EPI_ISL_4220330, EPI_ISL_4220331, EPI_ISL_4220332, EPI_ISL_4237192, EPI_ISL_4237193, EPI_ISL_4237194, EPI_ISL_4237195, EPI_ISL_4237197, EPI_ISL_4237198, EPI_ISL_4237201, EPI_ISL_4237202, EPI_ISL_4237203, EPI_ISL_4237204, EPI_ISL_4237205, EPI_ISL_4237206, EPI_ISL_4237207, EPI_ISL_4237208, EPI_ISL_4237209, EPI_ISL_4237210, EPI_ISL_4237211, EPI_ISL_4237212, EPI_ISL_4237242, EPI_ISL_4237243, EPI_ISL_4237244, EPI_ISL_4237245, EPI_ISL_4513382, EPI_ISL_4513383, EPI_ISL_4513386, EPI_ISL_4513387, EPI_ISL_4513388, EPI_ISL_4543243, EPI_ISL_4543244, EPI_ISL_4543245, EPI_ISL_4543246, EPI_ISL_4543247, EPI_ISL_4543248, EPI_ISL_4543249, EPI_ISL_4543250, EPI_ISL_4543251, EPI_ISL_4543252, EPI_ISL_4543253, EPI_ISL_4543254, EPI_ISL_4543255, EPI_ISL_4543256, EPI_ISL_4543257, EPI_ISL_4543258, EPI_ISL_4543259, EPI_ISL_4632895, EPI_ISL_4632901, EPI_ISL_4633865, EPI_ISL_4728720 | | | |
| see above | Laboratorio Central de Saude Publica do Estado do Rio de Janeiro (LACEN-RJ) | Laboratory of Respiratory Viruses and Measles, Oswaldo Cruz Institute, FIOCRUZ | Agatha Cristinne Prudencio; Alice Sampaio Rocha; Ana Carolina Mendonca; Andrea Cony Cavalcanti; Anna Carolina Paixao; Elisa Cavalcante Pereira; Fernando Motta; Ighor Leonardo Arantes Gomes; Luciana Appolinario; Marilda Siqueira on behalf of the Fiocruz COVID-19 Genomic Surveillance Network; Paola Resende; Renata Serrano Lopes; Taina Venas |
| EPI_ISL_3539830, EPI_ISL_3540004, EPI_ISL_3540005, EPI_ISL_3540006, EPI_ISL_3540007, EPI_ISL_3540008, EPI_ISL_3540009, EPI_ISL_3540010, EPI_ISL_3540011, EPI_ISL_3540012, EPI_ISL_3540013, EPI_ISL_3540014, EPI_ISL_3540016, EPI_ISL_3540017, EPI_ISL_3801837, EPI_ISL_3801838, EPI_ISL_3801845, EPI_ISL_3801846, EPI_ISL_3801881, EPI_ISL_3832389, EPI_ISL_3832390, EPI_ISL_4061331, EPI_ISL_4061333, EPI_ISL_4061334, EPI_ISL_4061339, EPI_ISL_4061341, EPI_ISL_4061342, EPI_ISL_4061343, EPI_ISL_4061345, EPI_ISL_4061346, EPI_ISL_4061349, EPI_ISL_4061351, EPI_ISL_4061354, EPI_ISL_4061355, EPI_ISL_4061356, EPI_ISL_4061357, EPI_ISL_4061358, EPI_ISL_4061359, EPI_ISL_4061360, EPI_ISL_4061361, EPI_ISL_4061362, EPI_ISL_4061363, EPI_ISL_4061365, EPI_ISL_4061366, EPI_ISL_4061369, EPI_ISL_4061370, EPI_ISL_4061372, EPI_ISL_4080771, EPI_ISL_4080775, EPI_ISL_4080776, EPI_ISL_4080777, EPI_ISL_4080778, EPI_ISL_4080786, EPI_ISL_4080789, EPI_ISL_4080790, EPI_ISL_4080791, EPI_ISL_4212782, EPI_ISL_4212783, EPI_ISL_4212784, EPI_ISL_4212785, EPI_ISL_4212786, EPI_ISL_4212787, EPI_ISL_4212788, EPI_ISL_4212789, EPI_ISL_4212790, EPI_ISL_4212791, EPI_ISL_4212792, EPI_ISL_4212793, EPI_ISL_4212794, EPI_ISL_4212795, EPI_ISL_4212796, EPI_ISL_4212797 | | | |
| see above | Laboratorio Central de Saude Publica do Estado do Rio de Janeiro (LACEN/RJ) | Laboratory of Respiratory Viruses and Measles, Oswaldo Cruz Institute, FIOCRUZ | Agatha Cristinne Prudencio Soares; Agatha Soares; Alice Sampaio Rocha; Ana Carolina Mendonca; Andrea Cony Cavalcanti; Anna Carolina Paixao; Elisa Cavalcante Pereira; Fernando Motta; Ighor Arantes; Ighor Leonardo Arantes Gomes; Luciana Appolinario; Marilda Siqueira on behalf of the Fiocruz COVID-19 Genomic Surveillance Network; Paola Resende; Renata Serrano Lopes; Taina Venas |
| EPI_ISL_5332849, EPI_ISL_5332852 | Laboratorio Central de la Ciudad de Santa Fe | Grupo de Genómica y Bioinformática del Instituto de Investigación de la Cadena Láctea CONICET-INTA on behalf of 'Proyecto Argentino Interinstitucional de genomica de SARS-CoV-2' (PAIS Consortium) | AF; Amadio; C; Eberhardt; G; Irazoqui; JM; MF; Mugna; Ojeda; Pastor; Rompato; V |
| EPI_ISL_3527816, EPI_ISL_3527823, EPI_ISL_3527828, EPI_ISL_4051044, EPI_ISL_4051047, EPI_ISL_4051056, EPI_ISL_4051066, EPI_ISL_4051068, EPI_ISL_4051070, EPI_ISL_4051101, EPI_ISL_4051103 | | | |
| see above | Laboratorio Nacional de Salud | Asociación de Salud Integral / Clínica Familiar "Luis Ángel García" | Ana S. Gonzalez-Reiche; Claudia Rangel; Danicela Mercado; Eduardo Arathon; Hilda Ruiz; Luis Aguirre; Luis Rivas; Narda Medina; Oscar Bonilla; Osmar Gamboa. |
| EPI_ISL_4601590, EPI_ISL_4601600, EPI_ISL_4601601, EPI_ISL_4601602 | Laboratorio de Investigaciones de Baney | Swiss Tropical and Public Health Institute | Bonifacio Manguire Nlavo; Carlos Cortes; Claudia Daubenberger; Diosdado Odjama Nseng Ada; Elizabeth Nyakarungu; Guillermo Garcia; Maximilian Mpina; Mitoha Ondo O Ayekaba; Philip Wonder Phiri; Philipp Wagner; Salome Hosch; Sarah Rubin; Tobias Schindler; Yahya Maidane |
| EPI_ISL_3987849 | Laboratorio de Referencia Nacional de Virus Respiratorios. Centro Nacional de Salud Publica. Instituto Nacional de Salud Peru. | Laboratorio de Referencia Nacional de Virus Respiratorios. Centro Nacional de Salud Publica. Instituto Nacional de Salud Peru. | Carlos Padilla Rojas; Henri Bailon Calderon; Iris Silva Molina; Joseph Huayra Niquen; Lely Solari Zerpa; Luis Barcena Flores; Marco Galarza Perez; Nancy Rojas Serrano; Nieves Sevilla Castañeda; Omar Caceres Rey; Orson Mestanza Millones; Princesa Medrano Alhuay; Priscila Lope Pari; Sandra Morales Ruiz; Sara Gordillo Vilchez; Steve Acedo Lazo; Veronica Hurtado Vela; Victor Jimenez Vasquez; Wendy Lizarraga Olivares |
| EPI_ISL_3045104 | Laboratorio de salud publica Puebla | LABOPAT | Cynthia Penaloza; Luis Mendoza; Silvia Montilla |
| EPI_ISL_4952583 | Laboratorios Clínicos Echandi | Inciensa, Investigación y Enseñanza en Nutrición y Salud | Adriana Godínez; Claudio Soto-Garita; Estela Cordero; Francisco Duarte; Hebleen Porras; José Luis Vargas; Mariela Gutiérrez & Joselyn Prado; Melany Calderón |
| EPI_ISL_4956251 | Laboratorios Delia Barraza | Microbial Genomics Laboratory | ; Alejandra García-Gasca; Alejandra Hernández-Terán; Alejandro Sánchez-Flores; Alfredo Herrera-Estrella; Alicia Ocaña-Mondragón; Andreu Comas-García; Angel Gustavo Salas-Lais; Antonio Loza Román; Bernardo Martínez-Miguel; Blanca Taboada; Brenda Irasema Maldonado-Meza; Bruno Gómez-Gil; Carla Ivón Herrera-Najera; Carlos F. Arias; Celia Boukadida; Clara Esperanza Santacruz-Tinoco; Claudia Soto Félix; Concepción Grajales-Muñiz; Consorcio Mexicano de Vigilancia Genómica (CoViGen-Mex). Authors (in alphabetical order): Julio Elias Alvarado-Yaah; Cristóbal Cháidez-Quiróz; Célida Duque Molina; Célida Martínez- Rodríguez; Daniel Fregoso-Rueda; Daniel Lira Morales; Eduardo Becerril-Vargas; Fernando Fontove-Herrera; Fidencio Mejía-Nepomuceno; Francisco Pulido; Gloria Elena Espinosa-Ayala; Gloria María Molina-Salinas; Gloria Vazquez; Hector Esteban Paz-Juárez; Hector Montoya-Fuentes; Helen Haydee Fernanda Ramírez-Plascencia; Irvin González-López; Jean Pierre González; Jesús Hernández; Joel Armando Vázquez-Pérez. Laboratorios Delia Barraza: María Delia Barraza Sámano; Jorge Salas-Hernández; José Antonio Enciso-Moreno; José Arturo Martínez-Orozco; José Esteban Muñoz-Medina; José de Jesús Nuñez-Contreras; Juan Bautista Chale-Dzul; Julissa Enciso-Ibarra; Luis Alberto Ochoa-Carrera; Margarita Matías-Florentino; Mario Mújica-Sánchez; Marissa Perez-Garcia; María Guadalupe Santiago-Mauricio; María Guadalupe de Jesús Mireles-Rivera; Melissa García Angulo.; Nelly Sélem-Mojica; Pavel Isa; Ricardo Ciria Merce; Ricardo Grande; Rosa María Gutiérrez Rios; Santiago Ávila-Ríos; Selene Zárate; Susana Lopez; Verónica Mata-Haro; Victor Eduardo García-Arias; Víctor Hugo Borja-Aburto |
| EPI_ISL_2927426, EPI_ISL_2927727, EPI_ISL_3016961, EPI_ISL_3325406, EPI_ISL_3509948, EPI_ISL_3512090, EPI_ISL_3512809, EPI_ISL_3514242, EPI_ISL_3515742, EPI_ISL_3519253, EPI_ISL_3605044, EPI_ISL_3675135, EPI_ISL_3687368, EPI_ISL_3741376, EPI_ISL_3746208, EPI_ISL_3750975, EPI_ISL_3751831, EPI_ISL_3751917, EPI_ISL_4155745, EPI_ISL_4158109, EPI_ISL_4162453, EPI_ISL_4164194, EPI_ISL_4174604, EPI_ISL_4242392, EPI_ISL_4335368, EPI_ISL_4337477, EPI_ISL_4486075, EPI_ISL_4486309, EPI_ISL_4486346, EPI_ISL_4809160, EPI_ISL_4814430, EPI_ISL_4815714, EPI_ISL_5074793, EPI_ISL_5074963, EPI_ISL_5078887, EPI_ISL_5081045, EPI_ISL_5186301, EPI_ISL_5189118, EPI_ISL_5190613, EPI_ISL_5237377, EPI_ISL_5264407 | | | |
| see above | Laboratory Corporation of America | Centers for Disease Control and Prevention Division of Viral Diseases, Pathogen Discovery | Adrian Paskey; Amanda Douglas; Amanda Suchanek; Andrea Throop; Ayla Burns; Benjamin Rambo-Martin; Bobbi Croy; Brian Krueger; Brian Norvell; Christopher Gulvick; Christos Petropoulos; Clinton Paden; Clinton R. Paden; Craig Lukasik; Dakota Howard; Darlene Wagner; Debbie Boles; Dhwani Batra; Duncan MacCannell; Eyad Almasri; Goran Stevovic; Howard Engler; Hrushikesh Deshmukh; Jake Humphrey; Jana Schroth; Jason Caravas; Joe Voshell; John Pruitt; Jonathan Meltzer; Jonathan Williams; Kara Moser; Kimberly Wagner; Kristine Lacek; Lax Iyer; Lisa Pfefferle; Lyndon Tilson; Manoj Jain; Marcia Eisenberg; Mary Ann Cristobal; Mary Cristobal; Mary Williamson; Matthew Robinson; Matthew Schmerer; Michael Levandoski; Mike Sapeta; Mindy Nye; Minoo Agarwal; Mohan Kolli; Nuthawin Charoensri; Oren Cohen; Peter Cook; Peter W. Cook; Prashant Gupta; Qian Zeng; Rama Ghatti; Scott Parker; Scott Ryan; Scott Sammons; Shatavia Morrison; Stanley Letovsky; Steven Ragan; Suresh Babu Selvaraju; Suresh Selvaraju; Susan Countryman; Susan Hicks; Suzanne Dale; Thomas Urban; Tim Kuphal; Tricia Zwiefelhofer; Tymeckia Kendall; Victoria Caban Figueroa; Vincent Drouillon; Yvette Unoarumhi |
| EPI_ISL_5160476, EPI_ISL_5160574 | Laboratory Medicine | Department of Laboratory Medicine, Lin-Kou Chang Gung Memorial Hospital, Taoyuan, Taiwan | Cheng-Hsun Chiu; Cheng-Ta Yang; Chung-Guei Huang; Guang-Wu Chen; Kuo-Chien Tsao; Kuo-Ming Lee; Mei-Jen Hsiao; Peng-Nien Huang; Po-Wei Huang; Shin-Ru Shih; Shu-Li Yang; Yi-Chun Liu; Yu-Nong Gong |
| EPI_ISL_4468868, EPI_ISL_4468870 | Laboratory for COVID19 diagnostics, Clinical Centre of Serbia | Institute of microbiology and Immunology, Faculty of Medicine, University of Belgrade | Banko A.; Cupic M.; Jankovic, M.; Jovanovic, T.; Knezevic, A.; Lazarevic I.; Milicevic, O.; Miljanovic D.; Sekler, M.; Tesovic, B.; Vidanovic, D. |
| EPI_ISL_3558549, EPI_ISL_3564510, EPI_ISL_4560396, EPI_ISL_4578352, EPI_ISL_5034806, EPI_ISL_5034848, EPI_ISL_5104594 | | | |
| see above | Laboratory for Molecular Diagnostics, IPHMN | Eurofins Genomics Europe Sequencing GmbH | Danijela Vujošević; Marija Govedarica |
| EPI_ISL_3631591 | Laboratory for Molecular Diagnostics,IPHMN | Eurofins Genomics Europe Sequencing GmbH | Danijela Vujošević |

| | | | |
|---|---|---|---|
| EPI_ISL_5310363 | Laboratory of Hygiene and Epidemiology, Department of Medicine, University of Thessaly | Greek Genome Center, Biomedical Research Foundation of the Academy of Athens (BRFAA) | Christos Hadjichristodoulou; Dimitrios Thanos; Dimitris Vrachnos; Emmanouil Athanasiadis; Giannis Vatsellas; Katerina Zoi; Maria Kiritsi; Theodoros Loupis |
| EPI_ISL_5312348, EPI_ISL_5312750, EPI_ISL_5312771 | Laboratory of Immunohematology, Division of Hematology | Greek Genome Center, Biomedical Research Foundation of the Academy of Athens (BRFAA) | Athanasia Mouzaki; Dimitrios Thanos; Dimitris Vrachnos; Emmanouil Athanasiadis; Giannis Vatsellas; Katerina Zoi; Theodoros Loupis |
| EPI_ISL_2893213, EPI_ISL_3062345, EPI_ISL_3342073 | Laboratory of Molecular Biology and Cancer Immunology, Faculty of Sciences, Lebanese University | Microbial Pathogenomics Lab - LAU | Bassam Badran; Fadi Abdel Sater; Georgi Merhi; Hamad Hassan; Jad Koweyes; Nada Ghosn; Rawan Makki; Sima tokajian; Tatiana Tohmeh |
| EPI_ISL_2982735, EPI_ISL_3540003, EPI_ISL_3801812, EPI_ISL_3801813, EPI_ISL_3801815, EPI_ISL_3801816, EPI_ISL_3801817, EPI_ISL_3801818, EPI_ISL_3801848, EPI_ISL_3801885, EPI_ISL_3801886, EPI_ISL_3801887, EPI_ISL_3801888, EPI_ISL_3801917, EPI_ISL_3801932, EPI_ISL_3801933, EPI_ISL_3802996, EPI_ISL_3832383, EPI_ISL_3832384, EPI_ISL_3832385, EPI_ISL_3832386, EPI_ISL_3832387, EPI_ISL_3832388, EPI_ISL_3832391, EPI_ISL_4061373, EPI_ISL_4061443, EPI_ISL_4061444, EPI_ISL_4061445, EPI_ISL_4061447, EPI_ISL_4061448, EPI_ISL_4080773, EPI_ISL_4080774, EPI_ISL_4170346, EPI_ISL_4170347, EPI_ISL_4170369, EPI_ISL_4212778, EPI_ISL_4212779, EPI_ISL_4212780, EPI_ISL_4212781, EPI_ISL_4220319, EPI_ISL_4237190, EPI_ISL_4237191, EPI_ISL_4237196, EPI_ISL_4237199, EPI_ISL_4237200, EPI_ISL_4237213, EPI_ISL_4237214, EPI_ISL_4237215, EPI_ISL_4237216, EPI_ISL_4237217, EPI_ISL_4237218, EPI_ISL_4237219, EPI_ISL_4237220, EPI_ISL_4237221, EPI_ISL_4237222, EPI_ISL_4237223, EPI_ISL_4237224, EPI_ISL_4237225, EPI_ISL_4237226, EPI_ISL_4237227, EPI_ISL_4237228, EPI_ISL_4237229, EPI_ISL_4237230, EPI_ISL_4237231, EPI_ISL_4237232, EPI_ISL_4237233, EPI_ISL_4237234, EPI_ISL_4237235, EPI_ISL_4237236, EPI_ISL_4237237, EPI_ISL_4237238, EPI_ISL_4237239, EPI_ISL_4237240, EPI_ISL_4237241, EPI_ISL_4237259, EPI_ISL_4237392, EPI_ISL_4257229, EPI_ISL_4257231, EPI_ISL_4257234, EPI_ISL_4257236, EPI_ISL_4257238, EPI_ISL_4257240, EPI_ISL_4257242, EPI_ISL_4257244, EPI_ISL_4257246, EPI_ISL_4257248, EPI_ISL_4257250, EPI_ISL_4257252, EPI_ISL_4257254, EPI_ISL_4257256, EPI_ISL_4257258, EPI_ISL_4257260, EPI_ISL_4271110, EPI_ISL_4271111, EPI_ISL_4271112, EPI_ISL_4271113, EPI_ISL_4418870, EPI_ISL_4513379, EPI_ISL_4513390, EPI_ISL_4513391, EPI_ISL_4513392, EPI_ISL_4543232, EPI_ISL_4543233, EPI_ISL_4543234, EPI_ISL_4543235, EPI_ISL_4543236, EPI_ISL_4543237, EPI_ISL_4543238, EPI_ISL_4543239, EPI_ISL_4543240, EPI_ISL_4543241, EPI_ISL_4543242, EPI_ISL_4632892, EPI_ISL_4633254, EPI_ISL_4633255 | | | |
| see above | Laboratory of Respiratory Viruses and Measles, Oswaldo Cruz Institute, FIOCRUZ | Laboratory of Respiratory Viruses and Measles, Oswaldo Cruz Institute, FIOCRUZ | Agatha Cristinne Prudencio; Agatha Cristinne Prudencio Soares; Agatha Soares; Alice Sampaio Rocha; Ana Carolina Mendonca; Andrea Cony Cavalcanti; Anna Carolina Paixao; Elisa Cavalcante Pereira; Fernando Motta; Ighor Arantes; Ighor Leonardo Arantes Gomes; Luciana Appolinario; Marilda Siqueira on behalf of the Fiocruz COVID-19 Genomic Surveillance Network; Paola Resende; Renata Serrano Lopes; Taina Venas; Valnete Andrade |
| EPI_ISL_3545711, EPI_ISL_3545713 | Laboratory of virology and molecular diagnostics, Institute of Public Health | Laboratory of virology and molecular diagnostics, Institute of Public Health | Boshevska G; Buzharova T; Janchevska E.; Kuzmanovska M |
| EPI_ISL_3460248 | Laboratório de Microbiologia Molecular - Universidade FEEVALE | Molecular Microbiology Laboratory | Alana Witt Hansen; Fernando Rosado Spilki; Fágner Henrique Heldt; Juliana Schons Gulart; Juliane Deise Fleck; Mariana Soares da Silva; Matheus Nunes Weber; Meriane Demoliner; Michele Filippi.; Paula Rodrigues de Almeida |
| EPI_ISL_3801795, EPI_ISL_3801796, EPI_ISL_3801797, EPI_ISL_3801798, EPI_ISL_3801803, EPI_ISL_3801804, EPI_ISL_3801805, EPI_ISL_3801806, EPI_ISL_3801807, EPI_ISL_3801819, EPI_ISL_3801820, EPI_ISL_3801821, EPI_ISL_3801822 | | | |
| see above | Labortorio Central de Saude Publica do Estado do Rio de Janeiro (LACEN/RJ) | Laboratory of Respiratory Viruses and Measles, Oswaldo Cruz Institute, FIOCRUZ | Agatha Cristinne Prudencio Soares; Alice Sampaio Rocha; Ana Carolina Mendonca; Andrea Cony Cavalcanti; Anna Carolina Paixao; Elisa Cavalcante Pereira; Fernando Motta; Ighor Leonardo Arantes Gomes; Luciana Appolinario; Marilda Siqueira on behalf of the Fiocruz COVID-19 Genomic Surveillance Network; Paola Resende; Renata Serrano Lopes; Taina Venas |
| EPI_ISL_4201642 | Lagos State Biobank, Lagos, Nigeria | 54gene Molecular Genetics Laboratory, Nigeria | Bankole Johnson; Chiamaka Nwuba; Chinyere Anyika; Dr Abasi Ene Obong; Dr Olajumoke Popoola; Dr Olukunke Oluwasemowo; Dr. Olufemi Obafemi; Emame Edu; Ifunanya Egoh; Lagos State Government; Mary Lalemi; Moyinoluwa Odugbemi; Nigerian Centre for Disease Control; Nonso Nduka; Ofonime Ebong; Oluwatimilehin Adewumi; Subomi Olorunnimbe; Tomiwa Adepetun |
| EPI_ISL_3184989, EPI_ISL_4552353 | Laverty Pathology | NSW Health Pathology - Institute of Clinical Pathology and Medical Research; Westmead Hospital; University of Sydney | Arnott A.; CIDM-PH et al.; Draper J.; Gall M.; Martinez E.; Rockett R.; Sintchenko V.; on behalf of ICPMR |
| EPI_ISL_5271291 | Lighthouse Lab in Alderley Park | Wellcome Sanger Institute for the COVID-19 Genomics UK (COG-UK) Consortium | Cordelia Langford; David K. Jackson; Dominic Kwiatkowski; Ewan Harrison; Ian Johnston; Jacquelyn Wynn; Jeffrey Barrett; John Sillitoe on behalf of the Wellcome Sanger Institute COVID-19 Surveillance Team; Mairead Hyland; Roberto Amato; Sonia Goncalves; The Lighthouse Lab in Alderley Park and Alex Alderton |
| EPI_ISL_2914932, EPI_ISL_2915241, EPI_ISL_2925083, EPI_ISL_3203327 | Lighthouse Lab in Glasgow | Wellcome Sanger Institute for the COVID-19 Genomics UK (COG-UK) Consortium | Anna Dominiczak and Alex Alderton; Carol Clugston; Cordelia Langford; David Gray; David K. Jackson; Dominic Kwiatkowski; Ewan Harrison; Harper VanSteenhouse; Ian Johnston; Jeffrey Barrett; John Sillitoe on behalf of the Wellcome Sanger Institute COVID-19 Surveillance Team; Roberto Amato; Sonia Goncalves; Yumi Kasai |
| EPI_ISL_1698079, EPI_ISL_1790800, EPI_ISL_5357885 | Lighthouse Lab in Milton Keynes | Wellcome Sanger Institute for the COVID-19 Genomics UK (COG-UK) Consortium | Cordelia Langford; David K. Jackson; Dominic Kwiatkowski; Ewan Harrison; Ian Johnston; Jeffrey Barrett; John Sillitoe on behalf of the Wellcome Sanger Institute COVID-19 Surveillance Team; Roberto Amato; Sonia Goncalves; The Lighthouse Lab in Milton Keynes and Alex Alderton |
| EPI_ISL_2852211 | Lighthouse Laboratory Plymouth | Wellcome Sanger Institute for the COVID-19 Genomics UK (COG-UK) Consortium | Cordelia Langford; David K. Jackson; Dominic Kwiatkowski; Ewan Harrison; Ian Johnston; Jeffrey Barrett; John Sillitoe on behalf of the Wellcome Sanger Institute COVID-19 Surveillance Team; Lighthouse Laboratory Plymouth and Alex Alderton; Roberto Amato; Sonia Goncalves |
| EPI_ISL_4301814 | Lubombo North RRT | National Institute for Communicable Diseases of the National Health Laboratory Service | Amoako DG; Bhiman JN; Everatt J; Ismail A; Mahlangu B; Maphalala G; Mnguni A; Mohale T; Ntuli N; Scheepers C |
| EPI_ISL_3073994 | MANKWENG LABORATORY | National Institute for Communicable Diseases of the National Health Laboratory Service | Amoako DG; Bhiman JN; Everatt J; Ismail A; Mahlangu B; Mnguni A; Mohale T; Ntuli N; Scheepers C |
| EPI_ISL_5434442, EPI_ISL_5434462 | MB-Cadham Provincial laboratory | Manitoba Cadham Provincial Laboratory | Anna Majer; Anneliese Landgraff; David Alexander; Elsie Grudeski; Gary Van Domselaar; Grace Seo; Jared Bullard; Jennifer Tanner; Kerry Dust; Morag Graham; Nat; Natalie Knox; Paul Van Caeseele; Philip Mabon; Rhiannon Huzarewich; Russell Mandes; Shari Tyson |
| EPI_ISL_4494367 | MB-Cadham Provincial laboratory | National Microbiology Laboratory (NML) | Anna Majer; Anneliese Landgraff; David Alexander; Elsie Grudeski; Gary Van Domselaar; Grace Seo; Jared Bullard; Jennifer Tanner; Kerry Dust; Morag Graham; Nat; Natalie Knox; Paul Van Caeseele; Philip Mabon; Rhiannon Huzarewich; Russell Mandes; Shari Tyson |
| EPI_ISL_4548462, EPI_ISL_4548464, EPI_ISL_4548519 | MRC/UVRI & LSHTM Uganda Research Unit, Uganda Virus Research Institute | CERI, Centre for Epidemic Response and Innvoation, Stellenbosch University and KRISP, KZN Research Innovation and Sequencing Platform, UKZN. | Deogratius Ssemwanga; Giandhari Jennifer; Hamidah Namagembe; Henry Kyobe-Bbosa; Isaac Ssewanyana; Jocelyn Kiconco; John Kayiwa; Julius Lutwama; Naidoo Yeshnee; Nicholas Bbosa; Pillay Sureshnee; Pontiano Kaleebu; Ronald Kiiza; San James; Susan Nabadda; Tegally Houriiyah; Tom Lutalo; Tshiabuila Derek; Upasana Ramphal; Wilkinson Eduan; Yajna Ramphal; and de Oliveira Tulio |
| EPI_ISL_3546307 | MRC/UVRI & LSHTM Uganda Research Unit, Central Public Health Laboratories | MRC/UVRI & LSHTM Uganda Research Unit, Central Public Health Laboratories | Dan Lule Bugembe; Hellen Nansumba; Isaac Sseeewanyana; Matthew Cotten; My V.T. Phan; Patrick Semanda; Pontiano Kaleebu; Susan Nabadda |
| EPI_ISL_3149352, EPI_ISL_3149362, EPI_ISL_3149363, EPI_ISL_3149365, EPI_ISL_3149369, EPI_ISL_3149372 | MRC/UVRI & LSHTM Uganda Research Unit, Central Public Health Laboratories | MRC/UVRI & LSHTM Uganda Research Unit, Central Public Health Laboratories | Dan Lule Bugembe; Hellen Nansumba; Isaac Sseeewanyana; Matthew Cotten; My V.T. Phan; Patrick Semanda; Pontiano Kaleebu; Susan Nabadda |
| EPI_ISL_3132295, EPI_ISL_3132296, EPI_ISL_3132311, EPI_ISL_3132317, EPI_ISL_3132331, EPI_ISL_3150942, EPI_ISL_3150943 | | | |
| see above | MRCG at LSHTM Genomics lab | MRCG at LSHTM Genomics lab | Abdoulie Kanteh; Abdul Karim sesay; Bakary Sanyang; Dabiri Damilari; Jarra Manneh; Mariama Kujabi |
| EPI_ISL_3714689 | MVZ Dr. Eberhard & Partner Dortmund | Robert Koch Institute | |
| EPI_ISL_4039227 | MVZ Labor Dr. Fenner und Kollegen (Standort Hamburg) | Robert Koch Institute | |
| EPI_ISL_2762465 | MVZ Labor Dr. Limbach & Kollegen GbR | Robert Koch Institute | |
| EPI_ISL_4739443, EPI_ISL_4739477, EPI_ISL_4739480 | Machakos Level 5 Hospital | KEMRI-Wellcome Trust Research Programme,Kilifi | Agoti C.; Githinji G.; Kituku A.; Lambisia A. Nokes D J.; Mburu M.W.; Mohamed K.S.; Morobe J.; Mutunga D.; Ndwiga L. Makori T.; Ochola I.; Ongera E.; de Laurent Z. |
| EPI_ISL_4629982, EPI_ISL_4629983, EPI_ISL_4629984, EPI_ISL_4748281, EPI_ISL_4750217, EPI_ISL_4750219 | Main Chemical Laboratories Egypt Army | Main Chemical Laboratories Egypt Army | AbedElrahman Zekri; Ahmed F Gad; Ahmed Gad; Ahmed S Elsafty; Ehab Abdelkader Abuelenein; Khaled Amer; Manal Hamdy Zahran; Mohamed Abdel-Salam Elgohary; Mohamed G Seadawy; Mostfa Elnakib; Mostfa F Elhoseiny; Raafat Zaher; Riad Mounir Armanious; Tarek Refaat Elnagdy; Wael Ali |
| EPI_ISL_4084057, EPI_ISL_5366603 | Maine Health and Environmental Testing Laboratory | Tewhey Lab, The Jackson Laboratory | Barter, M.; Dewey, H.; H. and Tewhey, R.; Iosue, F.; Lynch, R.; Matluk, N.; Munger |
| EPI_ISL_2785990, EPI_ISL_4167787 | Mako Medical | Centers for Disease Control and Prevention Division of Viral Diseases, Pathogen Discovery | Adrian Paskey; Benjamin Rambo-Martin; Christopher Gulvick; Clinton Paden; Clinton R. Paden; Dakota Howard; Darlene Wagner; Dhwani Batra; Duncan MacCannell; Jason Caravas; Kara Moser; Lauren Moon; Matthew Schmerer; Matthew Tugwell; Peter Cook; Peter W. Cook; Scott Sammons; Shatavia Morrison; Yvette Unoarumhi |
| EPI_ISL_3453899, EPI_ISL_3453909 | Maun Reference Hospital laboratory | Botswana Harvard HIV Reference Laboratory | Boitumelo J.L Zuze; Botshelo Radibe; Dorcas Maruapula; Joseph Makhema; Keoratile Ntshambiwa; Legodile Kooepile; Madisa Mine; Modisa Motswaledi; Mosepele Mosepele; Ontlametse T. Bareng; Roger Shapiro; Sandra Maripe; Shahin Lockman; Sikhulile Moyo; Simani Gaseitsiwe; Thongbotho Mphoyakgosi; Timothy Odirile Matsuokwane; Wonderful T. Choga |
| EPI_ISL_4301771, EPI_ISL_4301833 | Mbabane Government Hospital | National Institute for Communicable Diseases of the National Health | Amoako DG; Bhiman JN; Everatt J; Ismail A; Mahlangu B; Maphalala G; Mnguni A; Mohale T; Ntuli N; Scheepers C |

| | | | |
|---|---|---|---|
| | | Laboratory Service | |
| EPI_ISL_4301801 | Mbabane Public Health Unit | National Institute for Communicable Diseases of the National Health Laboratory Service | Amoako DG; Bhiman JN; Everatt J; Ismail A; Mahlangu B; Maphalala G; Mnguni A; Mohale T; Ntuli N; Scheepers C |
| EPI_ISL_3709110 | MedLab Central Ltd | Institute of Environmental Science and Research (ESR) | Anja Werno; Antje van der Linden; Arlo Upton; Chris Mansell; David Hammer; Dragana Drinkovic; Erasmus Smit; Gary McAuliffe; Hana Sofia Andersson; Hermes Perez; James Ussher; Jill Sherwood; Jing Wang; Joep de Ligt; Josh Freeman; Julia Howard; Juliet Elvy; Lauren Jelly; Mary DeAlmeida; Matt Blakiston; Matt Storey; Matthew Rogers; Max Bloomfield; Michael Addidle; Michelle Balm; Muhammad Faisal; Nikki Freed; Olin Silander; Olivia Stroeven; Rachel Boyle; Sally Roberts; SallyAnn Harbison; Sarah Jefferies; Sharmini Muttaiyah; Susan Morpeth; Susan Taylor; Timothy Blackmore; Vani Sathyendran; Veronica Playle; Virginia Hope; Xiaoyun Ren |
| EPI_ISL_3855282 | Medica | Institute of Medical Virology | Alexandra Trkola; Annette Audigé; Catharine Aquino; Cyril Shah; Daniel Ehrsam; Gabriela Ziltener; Guido Bloemberg; Hubert Rehrauer; Isabel Stürmer; Joel Wirz; Jon Huder; Jürg Böni; Kevin Steiner; Maria Grünberg; Maryam Zaheri; Michael Huber; Riccarda Capaul; Stefan Schmutz; Verena Kufner; Weihong Qi |
| EPI_ISL_4741161, EPI_ISL_4741165, EPI_ISL_4741173, EPI_ISL_4798130 | Medical Virology and BSL3 Laboratory | Centre de Séquençage Génomique | Abbad Anas; Amalou Ghita; Anga Latifa; Barakat Abdelhamid; Bouzidi Aymane; Charoute Hicham; Chgouri Fatima; Dersi Noureddine; EL Hamouchi Adil; El Oualid Abdelmjid; Faouzi Abdellah; Harmak Houda; Maaroufi Abderrahmane; Nadifiyine Saloua; Nourlil Jalal; Omondi Francis Carey; Somda Soro Georgina Charlene; Zemmouri Faouzia; Zouheir Yassine |
| EPI_ISL_4430724, EPI_ISL_4430745, EPI_ISL_4430751 | Medical Virology and BSL3 Laboratory | Laboratoire de Surveillance Génomique | Abbad Anas; Amalou Ghita; Anga Latifa; Barakat Abdelhamid; Bouzidi Aymane; Charoute Hicham; Chgouri Fatima; Dersi Noureddine; EL Hamouchi Adil; El Oualid Abdelmjid; Faouzi Abdellah; Harmak Houda; Maaroufi Abderrahmane; Nadifiyine Saloua; Nourlil Jalal; Omondi Francis Carey; Somda Soro Georgina Charlene; Zemmouri Faouzia; Zouheir Yassine |
| EPI_ISL_5305718, EPI_ISL_5306475, EPI_ISL_5306486 | Medlab Pathology | NSW Health Pathology - Institute of Clinical Pathology and Medical Research; Westmead Hospital; University of Sydney | Arnott A.; Draper J.; Gall M.; Martinez E.; Rockett R.; Sintchenko V.; on behalf of ICPMR |
| EPI_ISL_2839570 | Microbiological Diagnostic Unit - Public Health Laboratory (MDU-PHL) | MDU-PHL | M.L.; N.L.; Sait; Seemann T.; Sherry |
| EPI_ISL_4761411, EPI_ISL_4761424, EPI_ISL_4774248, EPI_ISL_4774615, EPI_ISL_4774750, EPI_ISL_4776931, EPI_ISL_4776988, EPI_ISL_4880934, EPI_ISL_4881024, EPI_ISL_4881658, EPI_ISL_4883731, EPI_ISL_4883740, EPI_ISL_5200570, EPI_ISL_5314295 | | | |
| see above | Microbiological Diagnostic Unit - Public Health Laboratory (MDU-PHL) | Microbiological Diagnostic Unit - Public Health Laboratory (MDU-PHL) | Horan, K.; N.L.; Seemann, T.; Sherry |
| EPI_ISL_5433681 | Microbiology Department, BPGHTC | Centre for Infectious Diseases, CSIR-NEIST | G Narahari Sastry; Gayatri Gogoi; Jatin Kalita; Moirangthem G Singh; Munmi Bora; Pankaj Bharali; Prasenjit Manna; Romi Wahengbam; Tridip Phukan; Yasmin B Tapaddar |
| EPI_ISL_5304866 | Microbiology Department. Complexo Hospitalario Universitario de Vigo | Microbiology Department. Complexo Hospitalario Universitario de Vigo | Alvarez M; Cabrera JJ; Carballo R; Cores O; Cortizo S; Davina C; Martinez L; Mediero G; Pena I; Perez S; Potel C; Regueiro B; Rey S; Vasallo FJ; del-Campo V |
| EPI_ISL_2964937, EPI_ISL_3164077, EPI_ISL_3164079, EPI_ISL_3869376, EPI_ISL_3948739, EPI_ISL_4005401, EPI_ISL_4433030, EPI_ISL_4518866, EPI_ISL_5032935, EPI_ISL_5033042, EPI_ISL_5033101, EPI_ISL_5348029 | | | |
| see above | Middlemore Hospital | Institute of Environmental Science and Research (ESR) | Anja Werno; Antje van der Linden; Arlo Upton; Chris Mansell; Clare Gebbie; David Hammer; Dhanisha Patel; Dragana Drinkovic; Erasmus Smit; Gary McAuliffe; Hana Sofia Andersson; Hermes Perez; James Ussher; Jill Sherwood; Jing Wang; Joep de Ligt; Josh Freeman; Julia Howard; Juliet Elvy; Lauren Jelly; Mary DeAlmeida; Matt Blakiston; Matt Storey; Matthew Rogers; Max Bloomfield; Michael Addidle; Michelle Balm; Muhammad Faisal; Nikki Freed; Olin Silander; Olivia Stroeven; Rachel Boyle; Sally Roberts; SallyAnn Harbison; Sarah Cockerton; Sarah Jefferies; Sharmini Muttaiyah; Susan Morpeth; Susan Taylor; Timothy Blackmore; Vani Sathyendran; Veronica Playle; Virginia Hope; Xiaoyun Ren |
| EPI_ISL_2958491, EPI_ISL_2987883, EPI_ISL_3032661, EPI_ISL_3306646, EPI_ISL_3405386, EPI_ISL_3538161, EPI_ISL_3548941, EPI_ISL_3565065, EPI_ISL_4073708, EPI_ISL_4096410, EPI_ISL_4265541, EPI_ISL_4892590, EPI_ISL_5417846, EPI_ISL_5417911 | | | |
| see above | Ministry of Health Turkey | Ministry of Health Turkey | Fatma Bayrakdar; Gulay Korukluoglu; Gülay Korukluoğlu; Suleyman Yalcin; Süleyman Yalcin; Yasemin Cosgun; Yasemin Coşgun |
| EPI_ISL_4419336, EPI_ISL_4940367 | Minnesota Department of Health, Public Health Laboratory | Minnesota Department of Health, Public Health Laboratory | Alexandra Lorentz; Jacob Garfin; Matt Plumb; and Xiong Wang |
| EPI_ISL_3010973, EPI_ISL_3065526, EPI_ISL_5420921 | Missouri State Public Health Laboratory | Missouri State Public Health Laboratory | Ashley New; Joshua Barry; Matthew Sinn |
| EPI_ISL_2970370, EPI_ISL_2970376, EPI_ISL_2970963, EPI_ISL_3052910 | Molecular Diagnostics Pathology Department Mater Dei Hospital Malta | Molecular Diagnostics Pathology Department Mater Dei Hospital Malta | C Cilia; G Zahra; L Grech; M Briffa; R Borg |
| EPI_ISL_3919555, EPI_ISL_3989318, EPI_ISL_3989353, EPI_ISL_3996739, EPI_ISL_3996814, EPI_ISL_4536644, EPI_ISL_4536930, EPI_ISL_5306785, EPI_ISL_5307088 | | | |
| see above | Molecular diagnostic laboratory of Federal Budget Institution of Science "Central Research Institute of Epidemiology" of The Federal Service on Customers' Rights Protection and Human Well-being Surveillance | Group of Genomics and Postgenomic Technologies of Central Research Institute of Epidemiology | Akimkin V.G.; Buharina A.Y.; Goncharov S.E.; Kondrasheva L.Y.; Korneenko E.V.; Nadtoka M.I.; Roev G.V.; Samoilov A.E.; Shipulina O.Y.; Sinitsyn S.O.; Smirnova Y.S.; Speranskaya A.S.; Svetlichnyj D.V.; Vyhodceva A.V. |
| EPI_ISL_3246829 | Montana Public Health Laboratory | Montana Public Health Laboratory | Carrie Biskupiak; Deborah Gibson; Joy Ritter; Michael Dills; Michelle Mozer |
| EPI_ISL_1663375 | NCCS, Pune | Institute of Life Sciences - INSACOG | Ajay Parida; Amol M. Kanampalliwar; Arup Ghosh; Atimukta Jha; INSACOG Consortium; Punit Prasad; Rajeeb Swain; Rupesh Dash; Safal Walia; Shifu Aggarwal; Sunil K. Raghav |
| EPI_ISL_3355441 | ND Dept. of Health Laboratory Services-Microbiology | Centers for Disease Control and Prevention Division of Viral Diseases, Pathogen Discovery | Alex Burgin; Ben Rambo-Martin; Clinton Paden; Dakota Howard; Dave Wentworth; Dhwani Batra; Jasmine Padilla; Justin Lee; Krista Queen; Kristen Knipe; Kristine Lacek; Mark Burroughs; Matthew Schmerer; Meghan Bentz; Mili Sheth; Peter Cook; Sam Shepard; Sarah Nobles; Suxiang Tong; Vivien Dugan; Yvette Unoarumhi |
| EPI_ISL_2987003 | NE Public Health Laboratory | Centers for Disease Control and Prevention Division of Viral Diseases, Pathogen Discovery | Alison Laufer Halpin; Ben L. Rambo-Martin; Clinton R. Paden; Dakota Howard; Darlene Wagner; Dave Wentworth; Dhwani Batra; Jasmine Padilla; Justin Lee; Katie Dillon; Krista Queen; Kristen Knipe; Kristine Lacek; Mark Burroughs; Matthew Schmerer; Mili Sheth; Peter Cook; Sam Shepard; Sarah Nobles; Shoshona Le; Suxiang Tong; Vivien Dugan; Yvette Unoarumhi |
| EPI_ISL_3132543 | NHLS Charlotte Maxeke Johannesburg Academic Hospital and the University of the Witwatersrand | KRISP, KZn Research Innovation and Sequencing Platform | Bulelani Manene; Florette Treurnicht; Giandhari Jennifer; Kathleen Subramoney; Naidoo Yeshnee; Pillay Sureshnee; San James; Tegally Houriiyah; Tshabuila Derek; Wilkinson Eduan; Yajna Ramphal; de Oliveira Tulio |
| EPI_ISL_3375627, EPI_ISL_3375628, EPI_ISL_3375631, EPI_ISL_3375634, EPI_ISL_5052208, EPI_ISL_5052209, EPI_ISL_5052210, EPI_ISL_5052212 | | | |
| see above | NIC, Viral Respiratory Unit | Virology Departement | Aicha Bensalem; Aissam Hachid; Amel Benyahia; Fawzi Derrar; Fayez Ahmed Khardine; Fayez Khardine; Fetouma Doudou; Mohamed Amine Beloufa |
| EPI_ISL_4061055 | NL-Dr. Leonard A. Miller Centre for Health Services | National Microbiology Laboratory (NML) | Adel Malek; Anna Majer; Anneliese Landgraff; Elsie Grudeski; Gary Van Domselaar; George Zahariad; Grace Seo; Jennifer Tanner; Laura Gilbert; Morag Graham; Natalie Knox; Philip Mabon; Rhiannon Huzarewich; Robert Needle; Russell Mandes; Shari Tyson; Yang Yu |
| EPI_ISL_5014617, EPI_ISL_5014626 | NL-Dr. Leonard A. Miller Centre for Health Services | Newfoundland and Labrador - Eastern Health | Anna Majer; Anneliese Landgraff; CanCOGeN's metadata curation team; Darian Hole; Elsie Grudeski; Gary Van Domselaar; George Zahariadis; Grace Seo; Jennifer Tanner; Kirsten Biggar; Madison Chapel; Morag Graham; Natalie Knox; Nathalie Bastien; Philip Mabon; Public Health Agency of Canada CanCOGeN team; Rhiannon Huzarewich; Robert Needle; Russell Mandes; Shari Tyson; Timothy Booth; Yan Li; Yang Yu |
| EPI_ISL_4508412, EPI_ISL_5439095 | NS-QEII Health Sciences Centre | National Microbiology Laboratory (NML) | Anna Majer; Anneliese Landgraff; Dan Gaston; Elsie Grudeski; Gary Van Domselaar; Grace Seo; Janice Pettipas; Jason LeBlanc; Jennifer Tanner; Morag Graham; Natalie Knox; Nathal; Philip Mabon; Rhiannon Huzarewich; Russell Mandes; Shari Tyson; Todd Hatchette |
| EPI_ISL_5327918, EPI_ISL_5327930, EPI_ISL_5327951 | Namibia Institute of Pathology LTD, Windhoek Central Hospital | CERI, Centre for Epidemic Response and Innvoation, Stellenbosch University and KRISP, KZN Research Innovation and Sequencing Platform, UKZN. | Andreas Shiningavamwe; Giandhari Jennifer; Iyaloo Constantinus; Naidoo Yeshnee; Nathalia !Garus-Oas; Ndahafa Frans; Ndumbu Pentikainen; Pillay Sureshnee; San James; Tegally Houriiyah; Tshiabuila Derek; Wilkinson Eduan; Yajna Ramphal; de Oliveira Tulio |
| EPI_ISL_3061823 | Nastavni zavod za javno zdravstvo Primorsko- Goranske Županije | Hrvatski zavod za javno zdravstvo | Irena Tabain; Ivana Ferenčak |
| EPI_ISL_5393740 | Nastavni zavod za javno zdravstvo Splitsko- Dalmatinske županije | Hrvatski zavod za javno zdravstvo | Irena Tabain; Ivana Ferenčak |
| EPI_ISL_3064705, EPI_ISL_3064708 | National Agency for Public Health, Republic of Moldov | Charité Universitätsmedizin Berlin, Institut für Virologie | Ala Halacu; Barbara Mühlemann; Christian Drosten; Julia Schneider; Jörn Beheim-Schwarzbach; Mariana Apostol; Talitha Veith; Terry Jones; Victor M Corman |
| EPI_ISL_3543611, EPI_ISL_3543621, EPI_ISL_3543640, EPI_ISL_3543642 | National Center of Disease Control and Prevention of the Republic of Armenia, Davidyants Laboratories, Yerevan, Armenia | Institute of Molecular Biology NAS RA, Republic of Armenia, Department of Bioengineering, BioinformaticsInstitute and Molecular Biology IBMPh RAU, Republic of Armenia | Andranik Chavushyan; Ani Melkonyan; Arsen Arakelyan; Diana Avetyan; Gayane Melik-Pashayan; Gisane Khachatyan; Hovsep Ghazaryan; Lilit Ghukasyan; Maria Nikoghosyan; Nelli Muradyan; Roksana Zakharyan; Shushan Sargrsyan; Siras Hakobyan; Tamara Sirunyan |
| EPI_ISL_3076737, EPI_ISL_3076741, EPI_ISL_3232679, EPI_ISL_3796518, EPI_ISL_4031610, EPI_ISL_4031635, EPI_ISL_4778719, EPI_ISL_5417426, EPI_ISL_5417466, EPI_ISL_5417527 | | | |
| see above | National Center of Infectious and Parasitic Diseases | National Center of Infectious and Parasitic Diseases | Alexiev et al |

| | | | |
|---|---|---|---|
| EPI_ISL_4474393 | National HIV Reference Laboratory, Ministry of Health, Public Health Institute of Malawi | CERI, Centre for Epidemic Response and Innovation | Auld A; Chilima B; Chiwaula M; Emmanuel S; Giandhari J; Kaba M; Kampira E; Kasambara W; Kim L; Maida A; Mvula B; Mwangomba W; Naidoo Yeshnee; Panja L; Pillay S; Tegally H; Tshabuila Derek; Wadonda N; Wilkinson E; Yajna Ramphal; de Oliveira T |
| EPI_ISL_3730373, EPI_ISL_3730374, EPI_ISL_3730378, EPI_ISL_4473204, EPI_ISL_4473223 | National HIV Reference Laboratory, Ministry of Health, Public Health Institute of Malawi | CERI, Centre for Epidemic Response and Innvoation, Stellenbosch University and KRISP, KZN Research Innovation and Sequencing Platform, UKZN. | Auld A; Chilima B; Chiwaula M; Emmanuel SJ; Giandhari J; Kaba M; Kampira E; Kasambara W; Kim L; Lessells R; Maida A; Mvula B; Mwangomba W; Naidoo Y; Panja L; Pillay S; Tegally H; Tshiabuila Derek; Wadonda N; Wilkinson E; de Oliveira T |
| EPI_ISL_3663621, EPI_ISL_3663640 | National HIV Reference Laboratory, Ministry of Health, Public Health Institute of Malawi | KRISP, KZn Research Innovation and Sequencing Platform | Auld A; Chilima B; Chiwaula M; Emmanuel SJ; Giandhari J; Kaba M; Kampira E; Kasambara W; Kim L; Lessells R; Maida A; Mvula B; Mwangomba W; Naidoo Y; Panja L; Pillay S; Tegally H; Tshiabuila Derek; Wadonda N; Wilkinson E; de Oliveira T |
| EPI_ISL_3948290, EPI_ISL_3948369 | National Health Laboratory | Botswana Harvard HIV Reference Laboratory | Boitumelo Zuze; Botshelo Radibe; Dorcas Maruapula; Joseph Makhema; Keoratile Ntshambiwa; Kgomotso Moruisi; Legodile Kooepile; Mosepele Mosepele; Mphaphi B. Mbulawa; Ontlametse T. Bareng; Pamela Smith-Lawrence; Patrick T. Mokgethi; Roger Shapiro; Sefetogi Ramaologa; Shahin Lockman; Sikhulile Moyo; Simani Gaseitsiwe; Thongbotho Mphoyakgosi; Wonderful T. Choga |
| EPI_ISL_3730296 | National Health Laboratory Service, South Africa | KRISP, KZn Research Innovation and Sequencing Platform | Giandhari Jennifer; Naidoo Yeshnee; Pillay Sureshnee; San James; Tegally Houriiyah; Tshiabuila Derek; Wilkinson Eduan; Yajna Ramphal; de Oliveira Tulio |
| EPI_ISL_4761813, EPI_ISL_4761823, EPI_ISL_4761834, EPI_ISL_4761835, EPI_ISL_4761836 | National Health Laboratory, Timor-Leste | Microbiological Diagnostic Unit - Public Health Laboratory (MDU-PHL) | Antonia da Costa, E.; Barreto, I.; Canisia, D.; Dakh, F.; Dolores de Jesus da Costa, M.; Douglas, N.; Francis, J.; Freeman, K.; Horan, K.; Jayanti Pereira Tilman, A.; Marr, I.; Salles de Sousa, A.; Seemann, T.; Sherry, N.; Soares da Silva, E.; Wapling, J.; Ximenes, J. |
| EPI_ISL_2455240, EPI_ISL_2455494, EPI_ISL_4748297, EPI_ISL_4942690, EPI_ISL_5458952 | National Hospital for Tropical Diseases | Oxford University Clinical Research Unit, Hanoi, Vietnam | H.Rogier van Doorn on behalf of the OUCRU COVID-19 research group; Le Van Duyet; Luu Thi Dieu Lien; Ngo Thi Thu Hang; Nguyen My Hanh; Nguyen Thi Hang; Nguyen Thi Hong Phuong; Nguyen Thi Hong Thuong; Nguyen Thi Kim Chi; Nguyen Thi Nhu Ha; Nguyen Thi Tam; Nguyen Thu Trang; Pham Ngoc Thach; Phan Manh Cuong; Thomas Kesteman; Tran Thi Van Dung; Trinh Cong Dien; Trinh Son Tung; Van Dinh Trang; Vu Ngoc Lien |
| EPI_ISL_4253831, EPI_ISL_4253839, EPI_ISL_4253847, EPI_ISL_4919704, EPI_ISL_4919711 | National Influenza Centre | National Influenza Centre | ; Benjamiin B. Lindsey; Benjamin H. Foulkes; Dennis Laryea; Ernest Asiedu; Franklin Asiedu-Bekoe; Gordon Awandare; Ivy A. Asante; Joseph Oliver-Commey; Joyce Ngoi; Linda Boatemaa; Lorreta Kwasah; Mathew D. Parker; Michael Marks; Mildred Adusei-Poku; Sharon Hsu; Thushan I de Silva; William K. Ampofo |
| EPI_ISL_2932519, EPI_ISL_3259535, EPI_ISL_3259537 | National Institute of Infectious Diseases-Prof. Dr. Matei Bals Molecular Diagnostics Laboratory | National Institute of Infectious Diseases-Prof. Dr. Matei Bals Molecular Diagnostics Laboratory | Corina Casangiu; Dan Otelea; Leontina Banica; Marius Surleac; Ovidiu Vlaicu; Petre Milu; Robert Hohan; Simona Paraschiv |
| EPI_ISL_3318989, EPI_ISL_4443910 | National Institute of Public Health | National Institute of Public Health | Alexander Nagy; Dusan Trnka; Helena Jirincova; Jaromira Vecerova; Timotej Suri |
| EPI_ISL_3588349, EPI_ISL_3588406 | National Institute of Public Health | State Veterinary Institute Prague | Alexander Nagy; Helena Jirincova; Jaromira Vecerova; Lenka Cernikova; Martina Stara; Timotej Suri |
| EPI_ISL_4949177, EPI_ISL_4949192, EPI_ISL_4949199 | National Institute of Public Health Burundi, WHO Burundi, ASLM Burundi, MRC/UVRI & LSHTM Uganda Research Unit | MRC/UVRI & LSHTM Uganda Research Unit, National Institute of Public Health Burundi, WHO Burundi, ASLM Burundi | Alexis Niyomwungere; Anatole Nkeshimana; Blessing T. Marondera; Butoyi Pascal; Cassien Nduwimana; Dan Lule Bugembe; Eric Kezakarayagwa; Francine Kabatesi; Jean Baptiste Niyibigira; Jerome Ndaruhutse; Jerome Nkurunziza; Joseph Nyandwi; Kouadio Theodore Yao; Matthew Cotten; My V.T. Phan; Xavier Crespin |
| EPI_ISL_4253069 | National Laboratory for Health, Environment and Food, OMM, Celje | NLZOH (National Laboratory for Health, Environment and Food) / CISLD (Clinical Institute of Special Laboratory Diagnostics), University Children's Hospital, University Medical Center Ljubljana | Aleksander Kocuvan; Aleksander Mahnic; Alenka Štorman; Ana Grom; Barbara Jenko Bizjan; Daša Kavka / Jernej Kovač; Kaja Tominc; Katarina Kozmos; Maja Rupnik; Marko Pokorn; Maruša Debeljak; Mateja Borinc; Maša Jarčič; Nika Gobec; Robert Šket; Sandra Janezic; Tadej Battelino; Tine Tesovnik; Tjaša Žohar Čretnik |
| EPI_ISL_2983016 | National Laboratory for Health, Environment and Food, OMM, Koper | NLZOH (National Laboratory for Health, Environment and Food) / CISLD (Clinical Institute of Special Laboratory Diagnostics), University Children's Hospital, University Medical Center Ljubljana | Aleksander Kocuvan; Aleksander Mahnic; Alenka Štorman; Ana Grom; Barbara Jenko Bizjan; Gašper Strugar; Kaja Tominc; Katarina Kozmos; Maja Rupnik; Marko Pokorn; Maruša Debeljak; Maša Jarčič; Mitja Rak / Jernej Kovač; Nika Gobec; Robert Šket; Sandra Janezic; Tadej Battelino; Tina Cvetković; Tine Tesovnik; Tjaša Žohar Čretnik |
| EPI_ISL_5105754 | National Laboratory for Health, Environment and Food, OMM, Kranj | NLZOH (National Laboratory for Health, Environment and Food) / CISLD (Clinical Institute of Special Laboratory Diagnostics), University Children's Hospital, University Medical Center Ljubljana | Aleksander Kocuvan; Aleksander Mahnic; Alenka Štorman; Ana Grom; Barbara Jenko Bizjan; Kaja Tominc; Katarina Kozmos; Maja Rupnik; Marjana Petrevčič / Jernej Kovač; Marko Pokorn; Maruša Debeljak; Mateja Ravnik; Maša Jarčič; Monika Korošec; Nika Gobec; Robert Šket; Sandra Janezic; Tadej Battelino; Tine Tesovnik; Tjaša Žohar Čretnik |
| EPI_ISL_3316645 | National Laboratory for Health, Environment and Food, OMM, Maribor | NLZOH (National Laboratory for Health, Environment and Food) / CISLD (Clinical Institute of Special Laboratory Diagnostics), University Children's Hospital, University Medical Center Ljubljana | Aleksander Kocuvan; Aleksander Mahnic; Alenka Štorman; Ana Grom; Andrej Golle / Jernej Kovač; Barbara Jenko Bizjan; Kaja Tominc; Katarina Kozmos; Maja Rupnik; Marko Pokorn; Maruša Debeljak; Maša Jarčič; Mojca Cimerman; Nika Gobec; Nika Volmajer; Robert Šket; Sandra Janezic; Tadej Battelino; Tine Tesovnik; Tjaša Žohar Čretnik |
| EPI_ISL_3722289, EPI_ISL_3730385, EPI_ISL_3730400, EPI_ISL_3730409, EPI_ISL_3730417, EPI_ISL_3730479, EPI_ISL_3730480 | | | |
| see above | National Microbiology Reference Laboratory, Ministry of Health, Harare, Zimbabwe | CERI, Centre for Epidemic Response and Innvoation, Stellenbosch University and KRISP, KZN Research Innovation and Sequencing Platform, UKZN. | Agnes Juru; Air Comodor Dr J. Chimedza; Charles Nyagupe; Dr Raiva Simbi; Emmanuel SJ; Giandhari J; Hlanai Gumbo; Kenneth Maeka; Naidoo Y; Pillay S; Tapfumanei Mashe; Tatenda Takawira; Tegally H; Wilkinson E; de Oliveira T |
| EPI_ISL_3547672, EPI_ISL_3547683, EPI_ISL_3547686, EPI_ISL_3547688, EPI_ISL_3547689, EPI_ISL_3547691, EPI_ISL_3547705, EPI_ISL_4232140 | | | |
| see above | National Public Health Institute of Liberia Reference Lab | Center for Infection and Immunity, Columbia University | Bode Shobayo; Jane MaCauley; Komal Jain; Mitali Mishra; Nischay Mishra; Thomas Briese; W. Ian Lipkin |
| EPI_ISL_4207293 | National Public Health Laboratory | National Public Health Laboratory/CSIR-Institute of Genomics and Integrative Biology | National Public Health Laboratory Team |
| EPI_ISL_1704834, EPI_ISL_2508685, EPI_ISL_2508922, EPI_ISL_2710145, EPI_ISL_2790378, EPI_ISL_2790386, EPI_ISL_3008988, EPI_ISL_3924390, EPI_ISL_3949675, EPI_ISL_4079339, EPI_ISL_4504114, EPI_ISL_5421290 | | | |
| see above | National Public Health Laboratory, National Centre for Infectious Diseases | National Public Health Laboratory, National Centre for Infectious Diseases | Benny Yeo; Grace Jie Yin Ngan; Grace Ngan Jie Yin; Katherine Ching; Lin Cui; Raymond Tzer Pin Lin; Royce Ang; Samuel Loo; Tze Minn Mak; Zhenyang Zhou |
| EPI_ISL_3184543 | National Public Health Laboratory/PPHL- Karnali | National Public Health Laboratory/CSIR-Institute of Genomics and Integrative Biology | National Public Health Laboratory Team /PPHL- Karnali Team |
| EPI_ISL_3602112, EPI_ISL_3681496, EPI_ISL_3681504 | National Virology Reference Laboratory | National Public Health Laboratory, National Centre for Infectious Diseases | Katherine Ching; Lin Cui; Raymond Tzer Pin Lin; Royce Ang; Zhenyang Zhou |
| EPI_ISL_3011450, EPI_ISL_3118108, EPI_ISL_3848764, EPI_ISL_3911951, EPI_ISL_4356447, EPI_ISL_4791636, EPI_ISL_4793416, EPI_ISL_5159822, EPI_ISL_5430951 | | | |
| see above | National Virus Reference Laboratory | National Virus Reference Laboratory | Charlene Bennett; Cillian F De Gascun; Gabriel Gonzalez; Jonathan Dean; Michael Carr; Zoe Yandle |
| EPI_ISL_4274958, EPI_ISL_4882583 | Nemocnice Trebic | University Hospital Brno, CMBG | Bezdicek Matej; Dolejska Monika; Dufkova Kristyna; Kristyna Dufkova; Lengerova Martina; Svaton Jan; Volfova Pavlina |
| EPI_ISL_5421201 | Nevada State Public Health Laboratory | Nevada State Public Health Laboratory | Andrew Gorzalski; Lynette Gumbleton; Mark Pandori |
| EPI_ISL_5264651 | New Kuwait German Lab | Virology Lab, Jaber Al Ahmad Hospital | Dr. Ebaa Al-Awadhi; Dr. Zahrah Buhamad; Estabraq Kathim; Haroon Masih; Khubaid-ur-Rehman |
| EPI_ISL_1315070, EPI_ISL_2107446, | New South Wales Health Pathology Royal Prince Alfred Hospital | Microbiology RPAH | Au, J.; Bull, R.; Deveson, I.; Foster, C.; Rawlinson, W.; Ruiz Silva, M.; Van Hal, S. |

| | | | |
|---|---|---|---|
| EPI_ISL_2811802, EPI_ISL_3072049, EPI_ISL_4570338 | | | |
| EPI_ISL_4205032, EPI_ISL_4235997, EPI_ISL_4236008 | Nigerian Centre for Disease Control (NCDC) | Africa Centre for Excellence for Genomics of Infectious Diseases (ACEGID), Redeemer's University | A.T.; Abechi; Ajogbasile; Akano; C.A.; C.T.; Eromon; F.V.; Folarin, O.; Happi; I.B.; J.N.; J.U.; K.O.; Kayode; Nosamiefan, I.; Oguzie; Olawoye; Olumade; Oluniyi; P.E.; P.S.; T.J.; Ugwu; Uwanibe |
| EPI_ISL_4634738 | Nordland Hospital - Bodo, Laboratory Department, Molecular Biology Unit | Norwegian Institute of Public Health, Department of Virology | Atiya R Ali; Debech Nadia; Engebretsen Serina Beate; Garcia Llorente Ignacio; Hilde Elshaug; Hilde Vollan; Jon Bråte; Kamilla Heddeland Instefjord; Karoline Bragstad; Kathrine Stene-Johansen; Line Victoria Moen; Marie Paulsen Madsen; Olav Hungnes; Pedersen Benedikte Nevjen; Rasmus Riis Kopperud |
| EPI_ISL_3268307, EPI_ISL_4206131, EPI_ISL_5347396 | North Dakota Department of Health, Public Health Laboratory | North Dakota Department of Health, Public Health Laboratory | Lisa Wingerter |
| EPI_ISL_4760509 | North Lantau Hospital | Hong Kong Department of Health | Alan K.L. Tsang; Edman T.K. Lam; Ken H.L. Ng; Peter C.W. Yip; Rickjason C.W. Chan |
| EPI_ISL_5033037 | North Shore Hospital | Institute of Environmental Science and Research (ESR) | Anja Werno; Antje van der Linden; Arlo Upton; Chris Mansell; Clare Gebbie; David Hammer; Dhanisha Patel; Dragana Drinkovic; Erasmus Smit; Gary McAuliffe; Hana Sofia Andersson; Hermes Perez; James Ussher; Jill Sherwood; Jing Wang; Joep de Ligt; Josh Freeman; Julia Howard; Juliet Elvy; Lauren Jelly; Mary DeAlmeida; Matt Blakiston; Matt Storey; Matthew Rogers; Max Bloomfield; Michael Addidle; Michelle Balm; Muhammad Faisal; Nikki Freed; Olin Silander; Olivia Stroeven; Rachel Boyle; Sally Roberts; SallyAnn Harbison; Sarah Cockerton; Sarah Jefferies; Sharmini Muttaiyah; Susan Morpeth; Susan Taylor; Timothy Blackmore; Vani Sathyendran; Veronica Playle; Virginia Hope; Xiaoyun Ren |
| EPI_ISL_4632890, EPI_ISL_4632891 | Nucleic Acid Testing Laboratory-Rwanda Biomedical Centre | Nucleic Acid Testing Laboratory-Rwanda Biomedical Centre | Alice Kabanda; Arlene Uwituze; Dieudonne Mutangana; Enatha Mukantwari; Esperance Umumararungu; Leon Mutesa; Robert Rutayisire; Sindayiheba Reuben; Umuringa Jeanne d'Arc; Yvan Butera |
| EPI_ISL_2828499, EPI_ISL_3012192, EPI_ISL_3012219, EPI_ISL_3012227, EPI_ISL_4415306, EPI_ISL_4459602, EPI_ISL_4459626 | | | |
| see above | Nucleic Acid Testing, National Reference Laboratory | GIGA Medical Genomics | Bouchra Boujemla; Esperence Umumararungu; Jacob Souopgui; Keith Durkin; Léon Mutesa; Maria Artesi; Marie-Pierre Hayette; Nathalie Renotte; Patrick Tuyisenge; Reuben Sindayiheba; Reuben Sindayiheba Patrick Tuyisenge; Robert Rutayisire; Sabin Nsanzimana; Swaibu Gatare; Sébastien Bontems; Vincent Bours; Yvan Butera |
| EPI_ISL_5316793, EPI_ISL_5316795 | OKMI | University Hospital Brno, CMBG | Bezdicek Matej; Dolejska Monika; Kristyna Dufkova; Lengerova Martina; Svaton Jan |
| EPI_ISL_3031501 | Oddar Meanchey Rapid Response Team | Virology Unit, Institut Pasteur du Cambodge | Cecile Troupin; Chau Darapheak; Chin Savuth; Erik A Karlsson; Kraing Sidonn; Leakhena Pum; Ly Sovann; Sophoannadedh Rath; Veasna Duong; Yi Sengdoeurn |
| EPI_ISL_3404416 | Oregon State Public Health Laboratory | Oregon State Public Health Laboratory | Eugene Yeboah; John Fontana and Shane Sevey; Laura Tsaknaridis; Rafia Razzaque; Vanda Makris |
| EPI_ISL_5333179 | Orenburg Clinical Hospital RZD Medicine | WHO National Influenza Centre Russian Federation | Andrey Komissarov; Artem Fadeev; Daria Danilenko; Denis Durchenkov; Dmitry Lioznov; Irina Samoryadova; Kirill Varchenko; Kseniya Komissarova; Maria Pisareva; Mikhail Bakaev; Nikita Yolshin; Oula Mansour; Svetlana Soboleva; Tamila Musaeva; Veronika Eder |
| EPI_ISL_5333147, EPI_ISL_5333158 | Orenburg Regional Clinical Hospital No 2 | WHO National Influenza Centre Russian Federation | Andrey Komissarov; Artem Fadeev; Daria Danilenko; Denis Durchenkov; Dmitry Lioznov; Irina Samoryadova; Kirill Varchenko; Kseniya Komissarova; Maria Pisareva; Mikhail Bakaev; Nikita Yolshin; Oula Mansour; Svetlana Soboleva; Tamila Musaeva; Veronika Eder |
| EPI_ISL_3424597, EPI_ISL_3424604 | Originating lab: Wales Specialist Virology Centre Sequencing lab: Pathogen Genomics Unit | Public Health Wales Microbiology Cardiff Wales Specialist Virology Centre | Alec Birchley; Alexander Adams; Amy Gaskin; Angela Marchbank; Bree Gatica-Wilcox; Catherine Moore; Jason Coombes; Joanne Watkins; Joel Southgate; Johnathan Evans; Laura Gifford; Lauren Gilbert; Lee Graham; Malorie Perry; Matthew Bull; Nicole Pacchiarini; Sally Corden; Sara Kumziene-Summerhayes; Sara Rey; Sarah Taylor; Simon Cottrell; Sophie Jones; Tom Connor |
| EPI_ISL_4124953, EPI_ISL_4126811 | Oslo University Hospital, Department of Microbiology | Norwegian Institute of Public Health, Department of Virology | Arvind Yegambaram Meenakshi Sundaram; Cathrine Fladeby; Garcia Llorente Ignacio; Gregor D. Gilfillan; Hilde Elshaug; Hilde Vollan; Jon Bråte; Kamilla Heddeland Instefjord; Karoline Bragstad; Kathrine Stene-Johansen; Line Victoria Moen; Lise Andresen; Mariann Nilsen; Mona Holberg-Petersen; Olav Hungnes; Pedersen Benedikte Nevjen; Pål Marius Bjørnstad; Rasmus Riis Kopperud; Teodora Plamenova Ribarska |
| EPI_ISL_3133814, EPI_ISL_4415203 | Ostfold Hospital Trust - Kalnes, Centre for Laboratory Medicine, Section for gene technology and infection serology | Norwegian Institute of Public Health, Department of Virology | Atiya R Ali; Debech Nadia; Engebretsen Serina Beate; Garcia Llorente Ignacio; Hilde Elshaug; Hilde Vollan; Jon Bråte; Kamilla Heddeland Instefjord; Karoline Bragstad; Kathrine Stene-Johansen; Line Victoria Moen; Marie Paulsen Madsen; Olav Hungnes; Pedersen Benedikte Nevjen; Rasmus Riis Kopperud |
| EPI_ISL_4236935, EPI_ISL_4951123 | Outre Mer | National Reference Center for Viruses of Respiratory Infections, Institut Pasteur, Paris | Angela Brisebarre; Antoine Talarmin; Camille Capel; Christophe Malabat; Corinne Maufrais; Etienne Simon-Lorière; Frédéric Lemoine; Julien Fumey; Louise Lefrançois; Marion Barbet; Maud Vanpeene; Méline Bizard; Patricia Tamby; Slim El Khiari; Slim El-Khiari; Sylvie Behillil; Sylvie Van der Werf; Vincent Enouf |
| EPI_ISL_4171179, EPI_ISL_4545368, EPI_ISL_4740962, EPI_ISL_4740971, EPI_ISL_5053995 | Outre mer | National Reference Center for Viruses of Respiratory Infections, Institut Pasteur, Paris | Angela Brisebarre; Antoine Talarmin; Camille Capel; Christophe Malabat; Corinne Maufrais; CéCile Herrmann; Didier Mattera; Emmanuelle Rousset Bourgoin; Etienne Simon-Lorière; Frédéric Lemoine; Julien Fumey; Louise Lefrançois; Marion Barbet; Maud Vanpeene; Méline Bizard; Slim El Khiari; Slim El-Khiari; Sylvaine Bastian; Sylvie Behillil; Sylvie Van der Werf; Vincent Enouf |
| EPI_ISL_4887358, EPI_ISL_4887376, EPI_ISL_5033344, EPI_ISL_5033348, EPI_ISL_5033349 | PHV-FSS | PHV-FSS | Chenwei Wang on behalf of Q-PHIRE Genomics; Liam Mcintyre on behalf of Q-PHIRE Genomics |
| EPI_ISL_4168760 | POLICLINICA HORTOLANDIA | Instituto Butantan | Antonio Jorge Martins; Claudia Renata dos Santos Barros; David Schlesinger; Debora Botequio Moretti; Dimas Tadeu Covas; Elaine Cristina Marqueze; Elaine Vieira Santos; Evandra Strazza Rodrigues; Heidge Fukumasu; Jayme Augusto de Souza-Neto; José Salvatore Leister Patané; Luiz Alcantara; Luiz Lehmann Coutinho; Maria Carolina Elias; Maurício Lacerda Nogueira; Rafael dos Santos Bezerra; Raul Machado Neto; Rejane Maria Tommasini Grotto; Ricardo Haddad; Sandra Coccuzzo Sampaio Vessoni; Simone Kashima; Svetoslav Nanev Slavov; Vincent Louis Viala |
| EPI_ISL_3453875, EPI_ISL_5248554, EPI_ISL_5248611 | Palapye Primary Hospital Laboratory | Botswana Harvard HIV Reference Laboratory | Boitumelo J. L Zuze; Boitumelo J.L Zuze; Botshelo Radibe; Dorcas Maruapula; Joseph Makhema; Keoratile Ntshambiwa; Kgomotso Moruisi; Legodile Kooepile; Letsibogo Gaoraelwe; Madisa Mine; Modisa Motswaledi; Mosepele Mosepele; Mphaphi B. Mbulawa; Ontlametse T. Bareng; Pamela Smith-Lawrence; Roger Shapiro; Sefetogi Ramaologa; Shahin Lockman; Sikhulile Moyo; Simani Gaseitsiwe; Thela Tefelo; Thongbotho Mphoyakgosi; Wonderful T. Choga |
| EPI_ISL_3446697 | Pandamaran Health Clinic | Institute for Medical Research, Infectious Disease Research Centre, National Institutes of Health, Ministry of Health Malaysia | Anasir MI; Azizan MA; Kamel K; Mohd Zawawi Z; Ramly N; Robert F; Suppiah J; Thayan R |
| EPI_ISL_3010695 | Pandemic Response Lab - NYC | Pandemic Response Lab, R&D | Cybill del Castillo; Dylan Law; Haiping Hao; Henry Lee; Isabel Fernandez Escapa; Jon Laurent; Melissa Hopkins; Michael Hammerling; Pradeep Bugga; Shinyoung Clair Kang; Sol Rey; William Ward |
| EPI_ISL_3477099, EPI_ISL_3477102, EPI_ISL_3477103, EPI_ISL_3477104, EPI_ISL_3477105 | PathLab Bay of Plenty | Institute of Environmental Science and Research (ESR) | Anja Werno; Antje van der Linden; Arlo Upton; Chris Mansell; David Hammer; Dragana Drinkovic; Erasmus Smit; Gary McAuliffe; Hana Sofia Andersson; Hermes Perez; James Ussher; Jill Sherwood; Jing Wang; Joep de Ligt; Josh Freeman; Julia Howard; Juliet Elvy; Lauren Jelly; Mary DeAlmeida; Matt Blakiston; Matt Storey; Matthew Rogers; Max Bloomfield; Michael Addidle; Michelle Balm; Muhammad Faisal; Nikki Freed; Olin Silander; Olivia Stroeven; Rachel Boyle; Sally Roberts; SallyAnn Harbison; Sarah Jefferies; Sharmini Muttaiyah; Susan Morpeth; Susan Taylor; Timothy Blackmore; Vani Sathyendran; Veronica Playle; Virginia Hope; Xiaoyun Ren |
| EPI_ISL_2713075, EPI_ISL_4392614, EPI_ISL_4602669 | PathWest Laboratory Medicine WA | PathWest Laboratory Medicine WA | PathWest Laboratory Medicine WA Microbial Surveillance Unit |
| EPI_ISL_1816922, EPI_ISL_1914668, EPI_ISL_1972909, EPI_ISL_2757656, EPI_ISL_2757657, EPI_ISL_2841831, EPI_ISL_2982303, EPI_ISL_3150200, EPI_ISL_3150201, EPI_ISL_3245743, EPI_ISL_3245745, EPI_ISL_3566730, EPI_ISL_3693294, EPI_ISL_3693297, EPI_ISL_3693302, EPI_ISL_3693307, EPI_ISL_4347505, EPI_ISL_5054901, EPI_ISL_5054902, EPI_ISL_5054903, EPI_ISL_5054907, EPI_ISL_5054908 | | | |
| see above | PathWest Laboratory Medicine WA | PathWest Laboratory Medicine WA Microbial Surveillance Unit | PathWest Laboratory Medicine WA Microbial Surveillance Unit |
| EPI_ISL_2691707, EPI_ISL_4392616 | PathWest Laboratory Medicine WA Microbial Surveillance Unit | PathWest Laboratory Medicine WA Microbial Surveillance Unit | Hospital Ave; Nedlands WA 6009; PathWest Laboratory Medicine WA Microbial Surveillance Unit; QEII Medical Centre |
| EPI_ISL_4720338, EPI_ISL_4724234 | Pathogen Genomics Center, National Institute of Infectious Diseases | Pathogen Genomics Center, National Institute of Infectious Diseases | Kentaro Itokawa; Makoto Kuroda; Masanori Hashino; Rina Tanaka; Tsuyoshi Sekizuka |
| EPI_ISL_4205830 | Pathology West - NSW Health Pathology | NSW Health Pathology - Institute of Clinical Pathology and Medical Research; Westmead Hospital; University of Sydney | Arnott A.; Draper J.; Gall M.; Martinez E.; Rockett R.; Sintchenko V.; on behalf of ICPMR |
| EPI_ISL_4742333 | Philippine General Hospital (PGH) | Philippine Genome Center | Alethea R. de Guzman; Anna Ong-Lim; Arianne A. Zamora; Benedict A. Maralit; Carlo M. Lapid; Celia Carlos; Cynthia P. Saloma; Devon Ray Pacial; Diomedes A. Cariño; Edsel Maurice Salvana; El King D. Morado; Elcid Aaron R. Pangilinan; Eva Maria Cutiongco-de la Paz; Francis A. Tablizo; Henrietta Marie Rodriguez; Jaime C. Montoya; Jan Michael C. Yap; Jarvin E. Nipales; Jo-Hannah S. Llames; John Q. Wong; Joshua Gregor A. Dizon; Juan Antonio R. Magalang; Karol Sophia Agape R. Padilla; Kenneth M. Kim; Kris P. Punayan; Kristina Patriz Dela Cruz; Lindsay Clare D.L. Carandang; Ma. Exanil Plantig; Marc Edsel C. Ayes; Maria Rosario Singh-Vergeire; Maria Sofia L. Yangzon; Marielle M. Gamboa; Marissa Alejandria; Niña Francesca Bustamante; Razel Nikka M. Hao; Renato Jacinto Q. Mantaring; Rianna Patricia S. Cruz; Shiela Mae M. Araiza; Yvonne Valerie Austria; Zipporah Mariebelle R. Enriquez; Zyrel V. Mollejon |
| EPI_ISL_4301817 | Phocweni Millitary Clinic | National Institute for Communicable Diseases of the National Health Laboratory Service | Amoako DG; Bhiman JN; Everatt J; Ismail A; Mahlangu B; Maphalala G; Mnguni A; Mohale T; Ntuli N; Scheepers C |
| EPI_ISL_2861601 | Platform BIS UZA/UAntwerpen | Labo Klinische Biologie, UZA | Basil Britto Xavier; Christine Lammens; Herman Goossens; Ines Verbesselt; Jasmine Coppens; Kathleen Holemans; Marie Le Mercier; Veerle Matheeussen |
| EPI_ISL_3946196 | Praava Health | Child Health Research Foundation | CHRF Bangladesh Genomics Team; Dr. Zaheed Husain; Shafiul Azam |
| EPI_ISL_3673672 | Private clinic of Biogen Med, Tashkent, Uzbekistan | Center of Genomics and bioinformatics, Bioinformatics laboratory | Abdurakhmon N Yusupov; Dilshod E Usmanov; Ibrokhim Y Abdurakhmonov; Khurshida A Ubaydullaeva; Mirzakamol S Ayubov; Mukhammadjon H Mirzakhmedov; Shukhrat E Shermatov; Zabardast T Buriev |

| | | | |
|---|---|---|---|
| EPI_ISL_5114179 | Public Health Authority of the Slovak Republic | Laboratory of Genomics and Bioinformatics, Comenius University Science Park | Anna Gičová; Diana Rusňáková; Jakub Styk; Jaroslav Budiš; Miroslav Böhmer; Tatiana Sedláčková; Tomáš Szemes |
| EPI_ISL_3981917, EPI_ISL_4413262, EPI_ISL_5396481, EPI_ISL_5433872, EPI_ISL_5433970 | Public Health Authority of the Slovak Republic | Public Health Authority of the Slovak Republic | Anna Gičová; Barbora Kotvasová; Dusan Loderer; Elena Tichá; Ivana Kasubova; Katarina Janikova; Lucia Ševčíková; Marian Grendar; Miroslav Böhmer; Pavol Mišenko; Terézia Vrabľová; Tomáš Szemes |
| EPI_ISL_2876219 | Public Health Laboratory, Minnesota Department of Health | University of Minnesota Genomics Center | Corbin Dirkx; Daryl M. Gohl; Jaquelyn Kuriger-Laber; John Garbe; and Sean Wang |
| EPI_ISL_3925036 | QUARANTINE CAMP | Hong Kong Department of Health | Alan K.L. Tsang; Edman T.K. Lam; Ken H.L. Ng; Peter C.W. Yip; Rickjason C.W. Chan |
| EPI_ISL_3571856 | Quadram Institute Bioscience | COVID-19 Genomics UK (COG-UK) Consortium | Alexander J Trotter; Alison E. Mather; Alp Aydin; Ana P. Tedim; Anastasia Kolyva; Andrew Bell; Andrew J. Page; Christopher Jeanes; Claire Stuart; Dave J. Baker; Ebenezer Foster-Nyarko; Gemma L. Kay; John Wain; Justin O'Grady; Leonardo de Oliveira Martins; Lewis G. Spurgin; Lindsay Coupland; Lizzie Meadows; Luke Bedford; Maria Diaz; Mark Webber; Martin Lott; Muhammed Yasir; Nabil-Fareed Alikhan; Ngozi Elumogo; Nicholas M. Thomson; Rachael Stanley; Rachel Gilroy; Reenesh Prakash; Rose K Davidson; Samir Dervisevic; Samuel Bloomfield; Sophie J. Prosolek; Steven Rudder; Thanh Le-Viet |
| EPI_ISL_2695790, EPI_ISL_2727638, EPI_ISL_2727641, EPI_ISL_2956014, EPI_ISL_3247180, EPI_ISL_3247208, EPI_ISL_3333221, EPI_ISL_3614798, EPI_ISL_3614835 | | | |
| see above | Queensland Health Forensic and Scientific Services | Queensland Health Forensic and Scientific Services | Chenwei Wang on behalf of Q-PHIRE Genomics; Q-PHIRE Genomics; Son Nguyen |
| EPI_ISL_1924796, EPI_ISL_2869165, EPI_ISL_2874723, EPI_ISL_2929226, EPI_ISL_2930143, EPI_ISL_3213562, EPI_ISL_3720367, EPI_ISL_4020941, EPI_ISL_4249596, EPI_ISL_5069057, EPI_ISL_5069173, EPI_ISL_5082189, EPI_ISL_5335915, EPI_ISL_5335980, EPI_ISL_5387580, EPI_ISL_5388196, EPI_ISL_5389110, EPI_ISL_5389377, EPI_ISL_5418088, EPI_ISL_5418321, EPI_ISL_5418384, EPI_ISL_5418444, EPI_ISL_5418645, EPI_ISL_5418915 | | | |
| see above | Quest Diagnostics Incorporated | Centers for Disease Control and Prevention Division of Viral Diseases, Pathogen Discovery | A. Gerasimova; A. Perez; Adrian Paskey; B. Anderson; Benjamin Rambo-Martin; Christopher Gulvick; Clinton Paden; Clinton R. Paden; Dakota Howard; Darlene Wagner; Dhwani Batra; Duncan MacCannell; Erisa Sula; F. Lacbawan; I. A. Shlyakhter; I. Shlyakhter; Jason Caravas; K. Livingston; K.E. Livingston; Kara Moser; Kristine Lacek; L. Bernstein; L.E. Bernstein; M. Hua; Matthew Schmerer; P. Tanpaiboon; Peter Cook; Peter W. Cook; R. Kagan; R. M. Kagan; R. Owen; R. Rolando; R. V. Rolando; S. H. Rosenthal; S. Rosenthal; Sammons; Scott; Scott Sammons; Shatavia Morrison; Tymeckia Kendall; Victoria Caban Figueroa; Y. Liu; Yvette Unoarumhi |
| EPI_ISL_4322354 | REUNILAB | CNR Virus des Infections Respiratoires - France SUD | Antonin Bal; Bruno Lina; Gregory Destras; Gwendolyne Burfin; Hadrien Regue; Laurence Josset; Martine Valette; Quentin Semanas |
| EPI_ISL_4824370 | RI State Health Laboratories | Centers for Disease Control and Prevention Division of Viral Diseases, Pathogen Discovery | Alex Burgin; Ben Rambo-Martin; Clinton Paden; Dakota Howard; Dave Wentworth; Dhwani Batra; Jasmine Padilla; Justin Lee; Krista Queen; Kristen Knipe; Kristine Lacek; Mark Burroughs; Matthew Schmerer; Meghan Bentz; Mili Sheth; Peter Cook; Sam Shepard; Sarah Nobles; Suxiang Tong; Vivien Dugan; Yvette Unoarumhi |
| EPI_ISL_2531894 | RIIP | National Reference Center for Viruses of Respiratory Infections, Institut Pasteur, Paris | Angela Brisebarre; Camille Capel; Christophe Malabat; Corinne Maufrais; Etienne Simon-Lorière; Frédéric Lemoine; Louise Lefrançois; Marion Barbet; Maud Vanpeene; Méline Bizard; StéPhanie Guyomard-Rabenirina; Sylvie Behillil; Sylvie Van der Werf; Vincent Enouf |
| EPI_ISL_4237137, EPI_ISL_4237159 | Reference Laboratory of the Ministry of Health Royal Victoria Gardens | Laboratory of Respiratory Viruses and Measles, Oswaldo Cruz Institute, FIOCRUZ | Agatha Cristinne Prudencio; Alice Sampaio Rocha; Ana Carolina Mendonca; Anna Carolina Paixao; Elisa Cavalcante Pereira; Fernando Motta; Ighor Leonardo Arantes Gomes; Indira Martins; Jessica Edwards; Luciana Appolinario; Marilda Siqueira on behalf of the Fiocruz COVID-19 Genomic Surveillance Network; Paola Resende; Renata Serrano Lopes; Taina Venas |
| EPI_ISL_5052353 | Regional Medical Sciences Center 12 Songkhla | National Institute of Health, Department of Medical Sciences, Ministry of Public Health, Thailand | Archawin Rojanawiwat; Natchaya Khiadsang; Nuttida Thongpramul; Pakorn Piromtong; Pilailuk Okada; Ratana Tacharoenmuang; Siripaporn Phuygun; Sittiporn Parnmen; Sunthareeya Waicharoen; Thanutsapa Thanadachakul; Warawan Wongboot; sirikanda wimol |
| EPI_ISL_5416155 | Regional Medical Sciences Center 6 Chonburi | National Institute of Health, Department of Medical Sciences, Ministry of Public Health, Thailand | Archawin Rojanawiwat; Natchaya Khiadsang; Nuttida Thongpramul; Pakorn Piromtong; Pilailuk Okada; Ratana Tacharoenmuang; Siripaporn Phuygun; Sittiporn Parnmen; Sunthareeya Waicharoen; Thanutsapa Thanadachakul; Warawan Wongboot; sirikanda wimol |
| EPI_ISL_3590624 | Regional Medical Sciences Center 9 Nakhon Ratchasima | National Institute of Health, Department of Medical Sciences, Ministry of Public Health, Thailand | ; Natchaya Khiadsang; Nuttida Thongpramul; Pakorn Piromtong; Pilailuk Okada; Ratana Tacharoenmuang; Siripaporn Phuygun; Sittiporn Parnmen; Sunthareeya Waicharoen; Thanutsapa Thanadachakul; Warawan Wongboot; sirikanda wimol |
| EPI_ISL_3122995, EPI_ISL_3122996 | Republican Children's Clinical Infectious Diseases Hospital | WHO National Influenza Centre Russian Federation | Alexey Masharsky; Andrey Komissarov; Artem Fadeev; Daria Danilenko; Dmitry Lioznov; Elena Nabieva; Georgii Bazykin; Kirill Varchenko; Ksenia Safina; Kseniya Komissarova; Maria Baturova; Maria Pisareva; Mikhail Bakaev; Nikita Yolshin; Oula Mansour; Tamila Musaeva; Veronika Eder |
| EPI_ISL_4117776, EPI_ISL_4122780 | Respiratory Virus Unit, Microbiology Services Colindale, Public Health England | COVID-19 Genomics UK (COG-UK) Consortium | PHE Covid Sequencing Team |
| EPI_ISL_4497792, EPI_ISL_5424979 | Rhode Island Department of Health | Infectious Disease Program, Broad Institute of Harvard and MIT | Adams, G.; Azevedo, K.; B.L.; B.W.; Bauer, M.; Birren; Carter, A.; Chaluvadi, S.; D.J.; DeRuff, K.; Gladden-Young, A.; Huard, R.; J.E.; K.J.; King, E.; Lagerborg, K.; Lemieux; Loreth, C.; Miller, A.; Normandin, E.; P.C.; Park; Pearlman, L.; Reilly, S.; Rudy, M.; Sabeti; Siddle; Tomkins-Tinch, C.; and MacInnis |
| EPI_ISL_4490287, EPI_ISL_4490312 | Rivers State University Teaching Hospital | Africa Centre for Excellence for Genomics of Infectious Diseases (ACEGID), Redeemer's University | A.T.; Abechi; Ajogbasile; Akano; C.A.; C.T.; Eromon; F.V.; Folarin, O.; Happi; I.B.; J.N.; J.U.; K.O.; Kayode; Nosamiefan, I.; Oguzie; Olawoye; Olumade; Oluniyi; P.E.; P.S.; T.J.; Ugwu; Uwanibe |
| EPI_ISL_2839563, EPI_ISL_2839567, EPI_ISL_2907544, EPI_ISL_2907545, EPI_ISL_3030412 | Royal Darwin Hospital Pathology | MDU-PHL | Caly L.; Druce J.; M.L.; Meumann, E.; N.L.; Sait; Seemann T.; Sherry |
| EPI_ISL_4761380, EPI_ISL_4761381, EPI_ISL_4761382, EPI_ISL_4761384, EPI_ISL_4761385 | Royal Darwin Hospital Pathology | Microbiological Diagnostic Unit - Public Health Laboratory (MDU-PHL) | Baird, R.; Horan, K.; Meumann, E.; N.L.; Seemann T.; Sherry |
| EPI_ISL_4768139 | Rwanda National Joint Taskforce COVID-100 | Africa Centre for Excellence for Genomics of Infectious Diseases (ACEGID), Redeemer's University | A.N.; A.T.; Abechi; Ahmed, M.; Ajogbasile; Akano; Alice Kabanda; Arlene Uwituze; Ayo-Ale, B.; Ayoade, F.; C.A.; C.T.; Chukwu, G.; Dieudonne Mutangana; Enatha Mukantwari; Eromon; Esperance Umumararungu; F.V.; Folarin, O.; Happi; I.B.; Izuwayo Gerard; J.N.; J.U.; K.O.; Kayode; Leon Mutesa; Murebwayire Clarisse; Nosamiefan, I.; Oguzie; Okolie, J.; Olawoye; Olumade; Oluniyi; Ope-Ewe, O.; P.E.; P.S.; Philip, C.; Robert Rutayisire; Sindayiheba Reuben; Sobajo, T.; T.J.; Ugwu; Umuringa Jeanne d'Arc; Uwanibe; Yvan Butera |
| EPI_ISL_4768053 | Rwanda National Joint Taskforce COVID-73 | Africa Centre for Excellence for Genomics of Infectious Diseases (ACEGID), Redeemer's University | A.N.; A.T.; Abechi; Ahmed, M.; Ajogbasile; Akano; Alice Kabanda; Arlene Uwituze; Ayo-Ale, B.; Ayoade, F.; C.A.; C.T.; Chukwu, G.; Dieudonne Mutangana; Enatha Mukantwari; Eromon; Esperance Umumararungu; F.V.; Folarin, O.; Happi; I.B.; Izuwayo Gerard; J.N.; J.U.; K.O.; Kayode; Leon Mutesa; Murebwayire Clarisse; Nosamiefan, I.; Oguzie; Okolie, J.; Olawoye; Olumade; Oluniyi; Ope-Ewe, O.; P.E.; P.S.; Philip, C.; Robert Rutayisire; Sindayiheba Reuben; Sobajo, T.; T.J.; Ugwu; Umuringa Jeanne d'Arc; Uwanibe; Yvan Butera |
| EPI_ISL_2462343, EPI_ISL_2462368, EPI_ISL_2839982, EPI_ISL_2839987, EPI_ISL_2839988, EPI_ISL_2839992, EPI_ISL_2839994, EPI_ISL_2839996, EPI_ISL_2839998, EPI_ISL_3071862, EPI_ISL_3710497, EPI_ISL_3710498, EPI_ISL_3710501, EPI_ISL_3710503, EPI_ISL_3710506, EPI_ISL_3710512, EPI_ISL_3710513, EPI_ISL_3923193, EPI_ISL_4263107, EPI_ISL_4263110, EPI_ISL_4263111, EPI_ISL_4263112, EPI_ISL_5264808, EPI_ISL_5264809, EPI_ISL_5264810, EPI_ISL_5264811, EPI_ISL_5264813 | | | |
| see above | SA Pathology | SA Pathology | Caitlin Selway; Chuan Kok Lim; Geoff Higgins; Ivan Bastian; Lex Leong; Mark Turra |
| EPI_ISL_4512724, EPI_ISL_4745740, EPI_ISL_5030959, EPI_ISL_5031045, EPI_ISL_5031053, EPI_ISL_5329463, EPI_ISL_5329482, EPI_ISL_5329518, EPI_ISL_5329578, EPI_ISL_5329589, EPI_ISL_5329646, EPI_ISL_5329762, EPI_ISL_5329766, EPI_ISL_5410054, EPI_ISL_5410192 | | | |
| see above | SALUD DIGNA | Instituto Nacional de Medicina Genomica | Abraham Campos-Romero; Cedro-Tanda A; Cruz-Islas Jazmin; Escobar-Arrazola MA; Garnica-Lopez Dora; Herrera-Montalvo LA.; Hidalgo-Miranda A; Luna-Ruiz Marco; Mendoza-Vargas A; Moreno-Camacho José Luis; Ramirez-Vega O; Rangel-DeLeon D; Reyes-Grajeda JP; Rodriguez-Gallegos Jorge; Yair Alfaro-Mora |
| EPI_ISL_4506714 | SARS-CoV-2 Sequencing Castilla y Leon-Spain Consortium | SARS-CoV-2 Sequencing Castilla y Leon-Spain Consortium | Antonio Orduña-Domingo; Carlos Fuster Foz; Carmen Aldea-Mansilla; Carmen Gimeno Crespo; David Abad; Gabriel March Rosello; Gregoria Megías Lobón; Jose María Eiros Bouza; M. Isabel Fernandez-Natal; Marta Dominguez-Gil; Marta Hernandez; María Antonia García Castro; Mª Fe Brezmes-Valdivieso; Noelia Arenal Andrés; Silvia Rojo; Sonsoles Garcinuño Pérez |
| EPI_ISL_3900876, EPI_ISL_5403547, EPI_ISL_5403557 | SARS-CoV-2 testing team, National Institute of Infectious Diseases | Pathogen Genomics Center, National Institute of Infectious Diseases | Hazuka Y Furihata; Kentaro Itokawa; Makoto Kuroda; Masanori Hashino; Masumichi Saito; Naomi Nojiri; Nozomu Hanaoka; Rina Tanaka; Tsuguto Fujimoto; Tsuyoshi Sekizuka |
| EPI_ISL_3242060, EPI_ISL_3355423 | SD Public Health Laboratory | Centers for Disease Control and Prevention Division of Viral Diseases, Pathogen Discovery | Alex Burgin; Ben L. Rambo-Martin; Ben Rambo-Martin; Clinton Paden; Clinton R. Paden; Dakota Howard; Dave Wentworth; Dhwani Batra; Jasmine Padilla; Justin Lee; Krista Queen; Kristen Knipe; Kristine Lacek; Mark Burroughs; Matthew Schmerer; Meghan Bentz; Mili Sheth; Peter Cook; Sam Shepard; Sarah Nobles; Suxiang Tong; Vivien Dugan; Yvette Unoarumhi |
| EPI_ISL_3981169 | SEEBMO | Instituto Nacional de Saude (INSA) | Borges et al |
| EPI_ISL_3262123 | SI «Public Health Center of MHU» | The Institute of Molecular Biology and Genetics of NASU | M.Tukalo et al. |
| EPI_ISL_3534708, EPI_ISL_5426832 | SK-Roy Romanow Provincial Laboratory | National Microbiology Laboratory (NML) | Anna Majer; Anneliese Landgraff; Elsie Grudeski; Gary Van Domselaar; Grace Seo; Jennifer Tanner; Kara Loos; Keith MacKenzie; Meredith Faires; Morag Graham; Natalie Knox; Philip Mabon; Rachel; Rhiannon Huzarewich; Russell Mandes; Ryan McDonald; Shari Tyson |
| EPI_ISL_4396347, EPI_ISL_5304122 | SYNLAB | Instituto Nacional de Saude (INSA) | Borges et al |
| EPI_ISL_4928752 | SYNLAB MVZ Hamburg | Robert Koch Institute | |
| EPI_ISL_5119399 | SYNLAB MVZ Leinfelden-Echterdingen | Robert Koch Institute | |
| EPI_ISL_5122219 | SYNLAB MVZ Leverkusen | Robert Koch Institute | |
| EPI_ISL_3239324, | SYNLAB MVZ Weiden | Robert Koch Institute | |

| | | | |
|---|---|---|---|
| EPI_ISL_4045033, EPI_ISL_4445922 | | | |
| EPI_ISL_2978464, EPI_ISL_3067877, EPI_ISL_3067952, EPI_ISL_3160650, EPI_ISL_3160840, EPI_ISL_3277659, EPI_ISL_3277855, EPI_ISL_3503471, EPI_ISL_3503610, EPI_ISL_3548243, EPI_ISL_4070102, EPI_ISL_4232624 | | | |
| see above | Salud Digna | Instituto Nacional de Medicina Genomica | Abraham Campos-Romero; Cedro-Tanda A; Cruz-Islas Jazmin; Escobar-Arrazola; Escobar-Arrazola MA; Garnica-Lopez Dora; Gonzalez-Barrera D; Herrera-Montalvo LA.; Hidalgo-Miranda A; Luna-Ruiz Marco; M; Mendoza-Vargas A; Moreno-Camacho José Luis; Munguia-Garza P; Ramirez-Vega O; Rangel-DeLeon D; Reyes-Grajeda JP; Rodriguez-Gallegos Jorge; Yair Alfaro-Mora |
| EPI_ISL_3912515 | Salud Digna, A.C | Andersen lab at Scripps Research | Abraham Garcia Gil; Jose Luis Moreno Camacho; Marco Antonio Luna Ruiz-Esparza; Miguel A. Fernandez Rojas; SEARCH Alliance with Abraham Campos Romero |
| EPI_ISL_3356283 | Serdang Hospital | Institute for Medical Research, Infectious Disease Research Centre, National Institutes of Health, Ministry of Health Malaysia | Anasir MI; Azizan MA; Kamel K; Mohd Zawawi Z; Ramly N; Robert F; Suppiah J; Thayan R |
| EPI_ISL_5158578, EPI_ISL_5158883, EPI_ISL_5159115, EPI_ISL_5159181, EPI_ISL_5159296, EPI_ISL_5159307 | Servicio Virosis Respiratorias-Departamento Virología-INEI | Instituto Nacional Enfermedades Infecciosas C.G.Malbran | Avaro M.; Baumeister E.; Benedetti E.; Campos J.; Cisterna D.; Dattero ME; De Belder D.; Haim MS.; Lorenzo F.; Molina V.; Perandones C.; Poklepovich T.; Pontoriero A.; Russo M.; Sanchez Loria J.; Tuduri E. |
| EPI_ISL_4880605, EPI_ISL_4880660, EPI_ISL_4880663, EPI_ISL_4880664, EPI_ISL_4880669, EPI_ISL_4880677, EPI_ISL_4880679, EPI_ISL_4880686, EPI_ISL_4880732, EPI_ISL_4880760, EPI_ISL_4880788 | | | |
| see above | Seychelles Public Health Laboratory | KEMRI-Wellcome Trust Research Programme,Kilifi | Agoti C.; Brigitte Pool; Githinji G.; Jude Gedeon; Lambisia A. Nokes D J.; Leon Biscornet.; Mburu M.W.; Meggy Louange; Mohamed K.S.; Morobe J.; Ndwiga L. Makori T.; Ochola I.; Ongera E.; de Laurent Z. |
| EPI_ISL_2834518, EPI_ISL_3022069, EPI_ISL_3667362, EPI_ISL_3667474 | Shamir Medical Center (Asaf Harofe) | Shamir Medical Center (Asaf Harofe) | Abu Hamad Ramzia; Adina Bar Chaim; Anna Vishnevsky; Chen Weiner; Nir Rainy; Patricia Benveniste-Lekovitz; Reut Sorek Abramovich; Yevgeni Yegorov |
| EPI_ISL_5332419 | Sirindhorn Hospital | Division of Genomic Medicine and Innovation support,Department of Medical Sciences, Ministry of Public Health, Thailand | Archawin Rojanawiwat; Jirapha Pakdee; Natthakul Bunneang; Nuanjun Wichukchinda; Penpitcha Thawong; Pilailuk Akkapaiboon Okada; Pundharika Piboonsiri; Surakameth Mahasirimongkol; Waritta Sawaengdee |
| EPI_ISL_3869277 | South Dakota Department of Health | University of Minnesota Genomics Center | Corbin Dirkx; Daryl M. Gohl; Jaquelyn Kuriger-Laber; John Garbe |
| EPI_ISL_2828070 | South Eastern Area Laboratory Services (SEALS) | NSW Health Pathology - Institute of Clinical Pathology and Medical Research; Westmead Hospital; University of Sydney | CIDM-PH et al. |
| EPI_ISL_3546329, EPI_ISL_3546335, EPI_ISL_3546337, EPI_ISL_3546339, EPI_ISL_3546340, EPI_ISL_3546343, EPI_ISL_3546345, EPI_ISL_3546346 | | | |
| see above | South Sudan Ministry of Health, WHO South Sudan, MRC/UVRI & LSHTM Uganda Research Unit | MRC/UVRI & LSHTM Uganda Research Unit, South Sudan Ministry of Health, WHO South Sudan | Abe G. Abias; Dan Lule Bugembe; Dennis Kenyi Lodiongo; James Ayei; John Rumunu; Joseph Francis Wamala; Juma John HM; Lul Lojok Deng; Matthew Cotten; My V.T. Phan; Pontiano Kaleebu; Richard Lino Loro Lako; Sudhir Bunga |
| EPI_ISL_3164113, EPI_ISL_3164115, EPI_ISL_3164116, EPI_ISL_3164119 | Southern Community Labs Dunedin | Institute of Environmental Science and Research (ESR) | Anja Werno; Antje van der Linden; Arlo Upton; Chris Mansell; David Hammer; Dragana Drinkovic; Erasmus Smit; Gary McAuliffe; Hana Sofia Andersson; Hermes Perez; James Ussher; Jill Sherwood; Jing Wang; Joep de Ligt; Josh Freeman; Julia Howard; Juliet Elvy; Lauren Jelly; Mary DeAlmeida; Matt Blakiston; Matt Storey; Matthew Rogers; Max Bloomfield; Michael Addidle; Michelle Balm; Muhammad Faisal; Nikki Freed; Olin Silander; Olivia Stroeven; Paula scholes; Rachel Boyle; Sally Roberts; SallyAnn Harbison; Sarah Jefferies; Sharmini Muttaiyah; Susan Lin; Susan Morpeth; Susan Taylor; Timothy Blackmore; Vani Sathyendran; Veronica Playle; Virginia Hope; Xiaoyun Ren |
| EPI_ISL_4078270 | Spital Langenthal | SRO AG | Alexander Imhof; Cedric Howald; Deborah Penet; Henri Pegeot; Ioannis Xenarios; Keith Harshman; Lorenzo Cerutti; Melyssa Elies |
| EPI_ISL_2812878, EPI_ISL_3398620, EPI_ISL_3502254 | St Vincent's Pathology (SydPath) | NSW Health Pathology - Institute of Clinical Pathology and Medical Research; Westmead Hospital; University of Sydney | Arnott A.; CIDM-PH et al.; Draper J.; Gall M.; Martinez E.; Rockett R.; Sintchenko V.; on behalf of ICPMR |
| EPI_ISL_5150310 | Stadtspital Triemli | Institute of Medical Virology | Alexandra Trkola; Annette Audigé; Catharine Aquino; Cyril Shah; Daniel Ehrsam; Gabriela Ziltener; Guido Bloemberg; Hubert Rehrauer; Isabel Stürmer; Joel Wirz; Jon Huder; Jürg Böni; Kevin Steiner; Maria Grünberg; Maryam Zaheri; Michael Huber; Riccarda Capaul; Stefan Schmutz; Verena Kufner; Weihong Qi |
| EPI_ISL_3537513, EPI_ISL_3672742, EPI_ISL_3762537, EPI_ISL_4302175 | State Hygienic Laboratory at the University of Iowa | State Hygienic Laboratory at the University of Iowa | Alankar Kampoowale; Anna Yakos; Benfer; Cindy Toll; Davis Rieckenberg; Erik Twait; Jeff; Jeff Benfer; Kris Eveland; Krishnaveni Sompallae; Kristen Zanon; Mariah Knutson; Mohammed Allam; Valerie Reeb; Wes Hottel |
| EPI_ISL_3932580, EPI_ISL_4728612 | State Laboratories Division, Hawaii State Department of Health | State Laboratories Division, Hawaii State Department of Health | Ayana Garnet; Daniel Strange; Drew Kuwazaki; Edward Desmond; Pamela O'Brien; Razvan Sultana |
| EPI_ISL_4062646, EPI_ISL_4062906 | Sultan Qaboos University Hospital, Department of Microbiology & Immunology, Molecular Biology Section | KU Leuven, Rega Institute, Clinical and Epidemiological Virology | Abdullah Balkhair; Azza Alqayoudhi; Faiza Alnamaani; Fatma BaAlawi; Ishraq Al Kindi; Khuloud Al Maamari; Omar Balkhair; Piet Maes; Tony Wawina-Bokalanga; Zakaryia Almuharrmi |
| EPI_ISL_2980538, EPI_ISL_2980766, EPI_ISL_3157778, EPI_ISL_3787429, EPI_ISL_3796816, EPI_ISL_3986402, EPI_ISL_4337520, EPI_ISL_4337848, EPI_ISL_4861116, EPI_ISL_5356462 | | | |
| see above | Swedish national genomic surveillance program of SARS-CoV-2 | The Public Health Agency of Sweden | Alma Brolund; Maria Lind Karlberg; Maximilian Riess; Swedish national genomic surveillance program of SARS-CoV-2 |
| EPI_ISL_5316862 | Synlab | University Hospital Brno, CMBG | Bezdicek Matej; Dolejska Monika; Kristyna Dufkova; Lengerova Martina; Svaton Jan |
| EPI_ISL_3506003, EPI_ISL_3506115, EPI_ISL_3506137, EPI_ISL_3506138, EPI_ISL_3507225, EPI_ISL_4106488, EPI_ISL_4171855, EPI_ISL_4503368, EPI_ISL_4895400, EPI_ISL_5460614, EPI_ISL_5460637 | | | |
| see above | Synlab Eesti OÜ | 1. Laboratory of Communicable Diseases (Estonia); 2. Eurofins Genomics Europe Sequencing GmbH | Liidia Dotsenko et al. |
| EPI_ISL_2844993 | Synlab MVZ Augsburg | Robert Koch Institute | |
| EPI_ISL_4547752 | Szpital Specjalistyczny im. H. Klimontowicza Medyczne Laboratorium Mikrobiologiczne | Wojewódzka Stacja Sanitarno-Epidemiologiczna w Katowicach | Beata Rozwadowska |
| EPI_ISL_3326353 | TGen North | TGen North | Brett Van Tassel; Chris French; Darrin Lemmer; Dave Engelthaler; Hayley Yaglom; Heather Centner; Jolene Bowers |
| EPI_ISL_3019476, EPI_ISL_3219440, EPI_ISL_3219442, EPI_ISL_5332912 | Temporary Specimen Collection Centre at the AsiaWorld-Expo | Hong Kong Department of Health | Alan K.L. Tsang; Edman T.K. Lam; Ken H.L. Ng; Peter C.W. Yip; Rickjason C.W. Chan |
| EPI_ISL_3655549, EPI_ISL_4055925, EPI_ISL_4055930, EPI_ISL_4055941 | The Caribbean Public Health Agency | Carrington Lab, Department of Preclinical Sciences, Faculty of Medical Sciences, The University of the West Indies, St Augustine Campus | Anushka Ramjag; Arianne Brown-Jordan; Avery Hinds; Christine V. F. Carrington; Christopher Oura; Gabriel Escobar; Jacquiline Bisesor-McKenzie; Nikita S. D. Sahadeo; Nuno Faria; Oliver Pybus; Rhonda Sealey-Thomas; Risha Singh; Sarah Hill; Simone Keizer-Beache; SueMin Nathaniel; Vernie Ramkissoon |
| EPI_ISL_3926030, EPI_ISL_3926053, EPI_ISL_3926262, EPI_ISL_3926467, EPI_ISL_3926977, EPI_ISL_3927691, EPI_ISL_3929336 | | | |
| see above | The National University Hospital of Iceland | deCODE genetics | Agnar Helgason; Alma Moller; Arna B Agustsdottir; Arnaldur Gylfason; Asgeir Sigurdsson; Aslaug Jonasdottir; Berglind Eiriksdottir; Bjarni Thorbjornsson; Brynjar O Jensson; Daniel F Gudbjartsson; Droplaug N Magnusdottir; Elisabet E Gardarsdottir; Emil A Thorarensen; Gardar Sveinbjornsson; Gisli Masson; Gudmundur Georgsson; Gudmundur L Norddahl; Gudrun Sigmundsdottir; Hakon Jonsson; Hannes Eggertsson; Hilma Holm; Ingileif Jonsdottir; Jona Saemundsdottir; Kamilla S Josefsdottir; Kari Stefansson; Karl G Kristinsson; Kjartan R Gudmundsson; Kristin E Sveinsdottir; Kristjan E Hjorleifsson; Louise le Roux; Maney Sveinsdottir; Olafia S Gretarsdottir; Olafur T Magnusson; Pall Melsted; Patrick Sulem; Run Fridriksdottir; Solvi Rognvaldsson; Thora R Gunnarsdottir; Thordur Kristjansson; Thorolfur Gudnason; Unnur Thorsteinsdottir |
| EPI_ISL_5416075 | Tripler Army Medical Center, Microbiology Department | Department of Clinical Investigation | Catherine Uyehara; Jonathan D'Ambrozio; Kendra Carter; Rachael Downham |
| EPI_ISL_5390726 | UAB "Rezus.lt" | Institute of Biotechnology, Life Sciences Center, Vilnius University | Albertas Timinskas; Alma Gedvilaite; Danguole Ziogiene; Emilija Vasiliunaite; Milda Norkiene |
| EPI_ISL_3341010 | UAB "Rezus.lt" | National Public Health Surveillance Laboratory | Ana Steponkiene; Danas Baksa; Jelena Razmuk; Lukas Vasionis; Lukas Zemaitis; Migle Gabrielaite; Svajune Muralyte |
| EPI_ISL_4447898 | UAB Diagnostikos laboratorija | National Public Health Surveillance Laboratory | Ana Steponkiene; Danas Baksa; Jelena Razmuk; Lukas Vasionis; Lukas Zemaitis; Migle Gabrielaite; Svajune Muralyte |
| EPI_ISL_4425726 | UAB InMedica | Hospital of Lithuanian University of Health Sciences (LSMU) Kaunas Clinics | Astra Vitkauskiene; Darius Cereskevicius; Inga Nasvytiene; Kristina Aleknaviciene; Mantas Sarauskas; Marius Sukys; Rasa Ugenskiene; Renaldas Jurkevicius; Rima Vainoriene; Rimvydas Jonikas; Zivile Zemeckiene |

| | | | |
|---|---|---|---|
| EPI_ISL_5052596 | UAB Medicina practica laboratorija | Hospital of Lithuanian University of Health Sciences (LSMU) Kaunas Clinics | Astra Vitkauskiene; Darius Cereskevicius; Inga Nasvytiene; Kristina Aleknaviciene; Mantas Sarauskas; Marius Sukys; Rasa Ugenskiene; Renaldas Jurkevicius; Rima Vainoriene; Rimvydas Jonikas; Zivile Zemeckiene |
| EPI_ISL_4886526 | UAN-LANIIA | Microbial Genomics Laboratory | ; Alejandra García-Gasca; Alejandra Hernández-Terán; Alejandro Sánchez-Flores; Alfredo Herrera-Estrella; Alicia Ocaña-Mondragón; Andreu Comas-García; Angel Gustavo Salas-Lais; Antonio Loza Román; Bernardo Martínez-Miguel; Blanca Taboada; Brenda Irasema Maldonado-Meza; Bruno Gómez-Gil; Carla Ivón Herrera-Najera; Carlos F. Arias; Celia Boukadida; Clara Esperanza Santacruz-Tinoco; Concepción Grajales-Muñiz; Consorcio Mexicano de Vigilancia Genómica (CoViGen-Mex). Authors (in alphabetical order): Julio Elias Alvarado-Yaah; Cristóbal Cháidez-Quiróz; Célida Duque Molina; Célida Martínez-Rodríguez; Daniel Fregoso-Rueda; Daniel Lira Morales; Eduardo Becerril-Vargas; Fernando Fontove-Herrera; Fidencio Mejía-Nepomuceno; Francisco Pulido; Gloria Elena Espinosa-Ayala; Gloria María Molina-Salinas; Gloria Vazquez; Hector Esteban Paz-Juárez; Hector Montoya-Fuentes; Helen Haydee Fernanda Ramírez-Plascencia; Irvin González-López; Jean Pierre González; Jesús Hernández; Joel Armando Vázquez-Pérez. LANIIA-Nayarit: Iván Girón Pérez.; Jorge Salas-Hernández; José Antonio Enciso-Moreno; José Arturo Martínez-Orozco; José Esteban Muñoz-Medina; José de Jesús Nuñez-Contreras; Juan Bautista Chale-Dzul; Julissa Enciso-Ibarra; Luis Alberto Ochoa-Carrera; Margarita Matías-Florentino; Mario Mújica-Sánchez; Marissa Perez-Garcia; María Guadalupe Santiago-Mauricio; María Guadalupe de Jesús Mireles-Rivera; Nelly Sélem-Mojica; Pavel Isa; Ricardo Ciria Merce; Ricardo Grande; Rosa María Gutiérrez Rios; Santiago Ávila-Ríos; Selene Zárate; Susana Lopez; Verónica Mata-Haro; Victor Eduardo García-Arias; Víctor Hugo Borja-Aburto |
| EPI_ISL_3236565, EPI_ISL_5195976 | UCC (Universidad Central del Caribe) | Grubaugh Lab - Yale School of Public Health | Anderson Brito; Andrea Firpo; Annie Watkins; Chaney Kalinich; Chantal Vogels; David Rivera Aponte; Ernesto Vazquez; Fabiola Cruz-Lopez; Fabiola Fontanet; Isabel Ott; Jessica Rothman; Joseph Fauver; Karolane Gonzalez; Kendall Billig; Mallery Breban; Mary Petrone; Natalie Machargo; Nathan Grubaugh; Patricia Serrano; Tania Mitwalli; Tara Alpert; Tobias Koch; Vianca Aponte |
| EPI_ISL_3914796 | UHTL, University Hospitals | UHTL, University Hospitals | Alouani, D.; Sadri, N.; Song, X. |
| EPI_ISL_2989285 | ULS Litoral Alentejano | Instituto Nacional de Saude (INSA) | Borges et al |
| EPI_ISL_5033409 | ULS Norte Alentejano | Instituto Nacional de Saude (INSA) and Instituto Gulbenkian de Ciencia (IGC) | Borges et al |
| EPI_ISL_2695334 | ULSM - Matosinhos | Instituto Nacional de Saude (INSA) | Borges et al |
| EPI_ISL_4747168 | UNIDADE DE PRONTO ATENDIMENTO UPA DRA ANA OLIVIA BENTIVOGLIO | Instituto Butantan | Antonio Jorge Martins; Claudia Renata dos Santos Barros; David Schlesinger; Debora Botequio Moretti; Dimas Tadeu Covas; Elaine Cristina Marqueze; Elaine Vieira Santos; Evandra Strazza Rodrigues; Heidge Fukumasu; Jayme Augusto de Souza-Neto; José Salvatore Leister Patané; Luiz Alcantara; Luiz Lehmann Coutinho; Maria Carolina Elias; Maurício Lacerda Nogueira; Rafael dos Santos Bezerra; Raul Machado Neto; Rejane Maria Tommasini Grotto; Ricardo Haddad; Sandra Coccuzzo Sampaio Vessoni; Simone Kashima; Svetoslav Nanev Slavov; Vincent Louis Viala |
| EPI_ISL_2810710, EPI_ISL_4396709, EPI_ISL_5304486 | UNILABS | Instituto Nacional de Saude (INSA) | Borges et al |
| EPI_ISL_4731216 | UNIT KESIHATAN AWAM HEBHK | UKM Medical Molecular Biology Institute (UMBI) | Mira Farzana binti Mohamad Mokhtar |
| EPI_ISL_4298226 | UPA DR FRANCO DA ROCHA | Instituto Butantan | Antonio Jorge Martins; Claudia Renata dos Santos Barros; David Schlesinger; Debora Botequio Moretti; Dimas Tadeu Covas; Elaine Cristina Marqueze; Elaine Vieira Santos; Evandra Strazza Rodrigues; Heidge Fukumasu; Jayme Augusto de Souza-Neto; José Salvatore Leister Patané; Luiz Alcantara; Luiz Lehmann Coutinho; Maria Carolina Elias; Maurício Lacerda Nogueira; Rafael dos Santos Bezerra; Raul Machado Neto; Rejane Maria Tommasini Grotto; Ricardo Haddad; Sandra Coccuzzo Sampaio Vessoni; Simone Kashima; Svetoslav Nanev Slavov; Vincent Louis Viala |
| EPI_ISL_2966757, EPI_ISL_3048168, EPI_ISL_3048171, EPI_ISL_3940081, EPI_ISL_3941764, EPI_ISL_4079160, EPI_ISL_4295237 | | | |
| see above | US Air Force School of Aerospace Medicine | US Air Force School of Aerospace Medicine | Amanda Javorina; Anthony Fries; Carol Garrett; Clarise Starr; Cole Anderson; Deanna Muehleman; Elizabeth Macias; Fritz Castillo; Jennifer Black; Jennifer Meyer; Kelsey Lanter; Sarah Purves; William Gruner |
| EPI_ISL_1789403, EPI_ISL_2802952 | UW Virology Lab | UW Virology Lab | Alexander Greninger; Hong Xie; Keith R Jerome; Lasata Shrestha; Meei-Li Huang; Michelle Lin; Nathan Breit; Noah R. Baker; Patrick Mathias; Pavitra Roychoudhury; Ricardo Perez; Robert J. Livingston; Sean Ellis; Shah Mohamed Bakhash; Tien V. Nguyen |
| EPI_ISL_2942479 | Unidad de Investigacion Biomedica de Zacatecas (UIBZ) | Unidad de Genomica Avanzada | ; Alejandra Garcia-Gasca; Alejandra Hernandez-Teran; Alejandro Sanchez-Flores; Alfredo Herrera-Estrella; Alicia Ocaña-Mondragon; Andreu Comas-Garcia; Angel Gustavo Salas-Lais; Antonio Loza Roman; Bernardo Martinez-Miguel; Blanca Taboada; Brenda Irasema Maldonado-Meza; Bruno Gomez-Gil; Carla Ivon Herrera-Najera; Carlos F. Arias; Celia Boukadida; Celida Duque Molina; Celida Martinez- Rodriguez; Clara Esperanza Santacruz-Tinoco; Concepcion Grajales-Muñiz; Consorcio Mexicano de Vigilancia Genomica (CoViGen-Mex). Authors (in alphabetical order): Julio Elias Alvarado-Yaah; Cristobal Chaidez-Quiroz; Daniel Fregoso-Rueda; Daniel Lira Morales; Eduardo Becerril-Vargas; Fernando Fontove-Herrera; Fidencio Mejia-Nepomuceno; Francisco Pulido; Gloria Elena Espinosa-Ayala; Gloria Maria Molina-Salinas; Gloria Vazquez; Hector Esteban Paz-Juarez; Hector Montoya-Fuentes; Helen Haydee Fernanda Ramirez-Plascencia; Irvin Gonzalez-Lopez; Jean Pierre Gonzalez; Jesus Hernandez; Joel Armando Vazquez-Perez.; Jorge Salas-Hernandez; Jose Antonio Enciso-Moreno; Jose Arturo Martinez-Orozco; Jose Esteban Muñoz-Medina; Jose de Jesus Nuñez-Contreras; Juan Bautista Chale-Dzul; Julissa Enciso-Ibarra; Luis Alberto Ochoa-Carrera; Margarita Matias-Florentino; Maria Guadalupe Santiago-Mauricio; Maria Guadalupe de Jesus Mireles-Rivera; Mario Mujica-Sanchez; Marissa Perez-Garcia; Nelly Selem-Mojica; Pavel Isa; Ricardo Ciria Merce; Ricardo Grande; Rosa Maria Gutierrez Rios; Santiago avila-Rios; Selene Zarate; Susana Lopez; Veronica Mata-Haro; Victor Eduardo Garcia-Arias; Victor Hugo Borja-Aburto |
| EPI_ISL_3557264, EPI_ISL_4299748, EPI_ISL_4299755, EPI_ISL_5193958 | Unidad de Investigación Biomédica de Zacatecas (UIBZ) | Centro de Investigación en Enfermedades Infecciosas (CIENI), Instituto Nacional de Enfermedades Respiratorias (INER) | Alejandra García-Gasca; Alejandra Hernández-Terán; Alejandro Sánchez-Flores; Alfredo Herrera-Estrella; Alicia Ocaña-Mondragón; Andreu Comas-García; Angel Gustavo Salas-Lais; Antonio Loza Román; Bernardo Martínez-Miguel; Blanca Taboada; Brenda Irasema Maldonado-Meza; Bruno Gómez-Gil; Carla Ivón Herrera-Najera; Carlos F. Arias; Celia Boukadida; Clara Esperanza Santacruz-Tinoco; Concepción Grajales-Muñiz; Consorcio Mexicano de Vigilancia Genómica (CoViGen-Mex). Authors (in alphabetical order): Julio Elias Alvarado-Yaah; Cristóbal Cháidez-Quiróz; Célida Duque Molina; Célida Martínez-Rodríguez; Célida Martínez-Rodríguez; Daniel Fregoso-Rueda; Daniel Lira Morales; Eduardo Becerril-Vargas; Eduardo Rivera-Martínez; Fernando Fontove-Herrera; Fidencio Mejía-Nepomuceno; Francisco Pulido; Gabriel Chavira-Trujillo; Gloria Elena Espinosa-Ayala; Gloria María Molina-Salinas; Gloria Vazquez; Hector Esteban Paz-Juárez; Hector Montoya-Fuentes; Helen Haydee Fernanda Ramírez-Plascencia; Irvin González-López; Jean Pierre González; Jesús Hernández; Joel Armando Vázquez-Pérez.; Jorge Salas-Hernández; José Antonio Enciso-Moreno; José Arturo Martínez-Orozco; José Esteban Muñoz-Medina; José de Jesús Nuñez-Contreras; Juan Bautista Chale-Dzul; Julissa Enciso-Ibarra; Kathia Elizabeth Tapia-Diaz; Luis Alberto Ochoa-Carrera; Margarita Matías-Florentino; Mario Mújica-Sánchez; Marissa Perez-Garcia; María Eugenia Jiménez-Corona; María Guadalupe Santiago-Mauricio; María Guadalupe de Jesús Mireles-Rivera; Nelly Sélem-Mojica; Pavel Isa; Ricardo Ciria Merce; Ricardo Grande; Rosa María Gutiérrez Rios; Rosario Vazquez-Larios; Santiago Ávila-Ríos; Selene Zárate; Susana Lopez; Verónica Mata-Haro; Victor Eduardo García-Arias; Víctor Hugo Borja-Aburto |
| EPI_ISL_4878949, EPI_ISL_4878953 | Unidad de Investigación Biomédica de Zacatecas (UIBZ) | Instituto de Biotecnología de la UNAM | ; Alejandra García-Gasca; Alejandra Hernández-Terán; Alejandro Sánchez-Flores; Alfredo Herrera-Estrella; Alicia Ocaña-Mondragón; Andreu Comas-García; Angel Gustavo Salas-Lais; Antonio Loza Román; Bernardo Martínez-Miguel; Blanca Taboada; Brenda Irasema Maldonado-Meza; Bruno Gómez-Gil; Carla Ivón Herrera-Najera; Carlos F. Arias; Celia Boukadida; Clara Esperanza Santacruz-Tinoco; Concepción Grajales-Muñiz; Consorcio Mexicano de Vigilancia Genómica (CoViGen-Mex). Authors (in alphabetical order): Julio Elias Alvarado-Yaah; Cristóbal Cháidez-Quiróz; Célida Duque Molina; Célida Martínez-Rodríguez; Daniel Fregoso-Rueda; Daniel Lira Morales; Eduardo Becerril-Vargas; Fernando Fontove-Herrera; Fidencio Mejía-Nepomuceno; Francisco Pulido; Gloria Elena Espinosa-Ayala; Gloria María Molina-Salinas; Gloria Vazquez; Hector Esteban Paz-Juárez; Hector Montoya-Fuentes; Helen Haydee Fernanda Ramírez-Plascencia; Irvin González-López; Jean Pierre González; Jesús Hernández; Joel Armando Vázquez-Pérez).; Jorge Salas-Hernández; José Antonio Enciso-Moreno; José Arturo Martínez-Orozco; José Esteban Muñoz-Medina; José de Jesús Nuñez-Contreras; Juan Bautista Chale-Dzul; Julissa Enciso-Ibarra; Kathia Elizabeth Tapia-Diaz; Luis Alberto Ochoa-Carrera; Margarita Matías-Florentino; Mario Mújica-Sánchez; Marissa Perez-Garcia; María Guadalupe Santiago-Mauricio; María Guadalupe de Jesús Mireles-Rivera; Nelly Sélem-Mojica; Pavel Isa; Ricardo Ciria Merce; Ricardo Grande; Rosa María Gutiérrez Rios; Santiago Ávila-Ríos; Selene Zárate; Susana Lopez; Verónica Mata-Haro; Victor Eduardo García-Arias; Víctor Hugo Borja-Aburto |
| EPI_ISL_3347918, EPI_ISL_4006372 | Unidad de Investigación Médica de Yucatán (UIMY) | Instituto de Biotecnología de la UNAM | ; Alejandra García-Gasca; Alejandra Hernández-Terán; Alejandro Sánchez-Flores; Alfredo Herrera-Estrella; Alicia Ocaña-Mondragón; Andreu Comas-García; Angel Gustavo Salas-Lais; Antonio Loza Román; Bernardo Martínez-Miguel; Blanca Taboada; Brenda Irasema Maldonado-Meza; Bruno Gómez-Gil; Carla Ivón Herrera-Najera; Carlos F. Arias; Celia Boukadida; Clara Esperanza Santacruz-Tinoco; Concepción Grajales-Muñiz; Consorcio Mexicano de Vigilancia Genómica (CoViGen-Mex). Authors (in alphabetical order): Julio Elias Alvarado-Yaah; Cristóbal Cháidez-Quiróz; Célida Duque Molina; Célida Martínez-Rodríguez; Daniel Fregoso-Rueda; Daniel Lira Morales; Eduardo Becerril-Vargas; Fernando Fontove-Herrera; Fidencio Mejía-Nepomuceno; Francisco Pulido; Gloria Elena Espinosa-Ayala; Gloria María Molina-Salinas; Gloria Vazquez; Hector Esteban Paz-Juárez; Hector Montoya-Fuentes; Helen Haydee Fernanda Ramírez-Plascencia; Irvin González-López; Jean Pierre González; Jesús Hernández; Joel Armando Vázquez-Pérez).; Jorge Salas-Hernández; José Antonio Enciso-Moreno; José Arturo Martínez-Orozco; José Esteban Muñoz-Medina; José de Jesús Nuñez-Contreras; Juan Bautista Chale-Dzul; Julissa Enciso-Ibarra; Kathia Elizabeth Tapia-Diaz; Luis Alberto Ochoa-Carrera; Margarita Matías-Florentino; Mario Mújica-Sánchez; Marissa Perez-Garcia; María Guadalupe Santiago-Mauricio; María Guadalupe de Jesús Mireles-Rivera; Nelly Sélem-Mojica; Pavel Isa; Ricardo Ciria Merce; Ricardo Grande; Rosa María Gutiérrez Rios; Santiago Ávila-Ríos; Selene Zárate; Susana Lopez; Verónica Mata-Haro; Victor Eduardo García-Arias; Víctor Hugo Borja-Aburto |
| EPI_ISL_3014467 | Unidade de apoio ao diagnostico da COVID - UNADIG | Bioinformatics Laboratory / LNCC | Alessandra P Lamarca; Alexandra L Gerber; Amilcar Tanuri; Ana Paula de C Guimaraes; Ana Tereza R Vasconcelos; Andrea Cony Cavalcanti; Caio Luiz Pereira Ribeiro; Cassia Alves; Cintia Policarpo; Claudia Maria Braga de Mello; Cristiane Gomes da Silva; Diana Mariani; Douglas Terra Machado; Flavio Dias da Silva; Gleidson da Silva de Oliveira; Leandro Magalhaes de Souza; Liliane Cavalcante; Luiz G P de Almeida; Marcio Henrique de Oliveira Garcia; Mario Sergio Ribeiro; Ronaldo da Silva F Jr; Silvia Carvalho |
| EPI_ISL_3123561, EPI_ISL_3123566, EPI_ISL_3125816 | Unit of laboratory surveillance of viral emergent disease Control of Communicable Diseases Departament National Institute of Public Health Albania | Charité Universitätsmedizin Berlin, Institut für Virologie | Barbara Mühlemann; Christian Drosten; Iris Hasibra; Julia Schneider; Jörn Beheim-Schwarzbach; Talitha Veith; Terry Jones; Victor M Corman |
| EPI_ISL_3721611 | Universidad Nacional sede Medellin Laboratorio One Health | Universidad Nacional de Colombia - Laboratorio Genómico One Health | Andres F. Cardona-Rios; Carlos Franco-Muñoz; Carolina Muñoz-Arango; Celeny Ortiz; Daniel O. Maldonado-Perez; Diego A. Álvarez-Díaz; Hector Alejandro Ruiz-Moreno; Idabely Betancur Ortiz; Jorge E. Osorio; Juan P. Hernandez-Ortiz; Karl A Ciuoderis; Katherine Laiton-Donato; Laura Silvana Perez; Lina M. Hurtado; Marcela Mercado-Reyes; Maria Angélica Maya; Maria Stella López; Rita Almanza Payares; Sandra Ines Cano; Simón Villegas Velásquez |
| EPI_ISL_3076937 | University Clinical Hospital of Mostar, Department of Microbiology and Molecular Diagnostics | University of Sarajevo, Veterinary Faculty, Laboratory for Molecular Diagnostic and Research Laboratory | Goletic S; Goletic T; Hodzic A.; Jazic A.; Nicevic M.; Ostojic M; Sabic E.; Softic A; Terzic I. |
| EPI_ISL_4273949 | University Clinical Research Center (UCRC) | Northwestern University - Center for Pathogen Genomics and Microbial Evolution | Almoustapha I. Maiga; Anou M. Somboro; Egon A. Ozer; Ibrahima Guindo; Judd F. Hultquist; Lacy M. Simons; Mamoudou Maiga; Ramon Lorenzo-Redondo |
| EPI_ISL_3048361 | University Hospitals of Geneva, Laboratory of Virology | HUG, Laboratory of Virology and the Health2030 Genome Center | Ana Rita Goncalves; Deborah Penet; Emmanouil Dermitzakis; Henri Pegeot; Ioannis Xenarios; Keith Harshman; Laurent Kaiser; Lorenzo Cerutti; Melyssa Elies; Samuel Cordey |
| EPI_ISL_3980454 | University of Health and Allied Sciences (UHAS) COVID-19 Testing and Research Centre | University of Health and Allied Sciences (UHAS) COVID-19 Testing and Research Centre | John O. Gyapong and the UHAS COVID-19 Lab Team; Jones Gyamfi; Kwabena O. Duedu; Reuben Ayivor-Djanie |
| EPI_ISL_3120189 | University of Liège COVID-19 testing center | GIGA Medical Genomics | Bouchra Boujemla; Claire Gourzonès; Cécile Meex; Keith Durkin; Laurent Gillet; Maria Artesi; Marie-Pierre Hayette; Nadine Cambisano; Nathalie Renotte; Olivier Ek; Sébastien Bontems; Vincent Bours |
| EPI_ISL_3492940, EPI_ISL_4003296, EPI_ISL_5422065 | University of Mississippi Medical Center, Department of Pathology | University of Mississippi Medical Center, Molecular and Genomics Core Facility | Ashley C. Johnson; D. Ashley Robinson; Derrick D. Allen; Ithiel J. Frame; Krishna K. Ayyalasomayajula; Michael R. Garrett; Wenjie Wu |
| EPI_ISL_5257993, EPI_ISL_5258025 | University of Sarajevo, Veterinary Faculty, Laboratory for Molecular Diagnostic and Research Laboratory | University of Sarajevo, Veterinary Faculty, Laboratory for Molecular Diagnostic and Research Laboratory | Goletic S; Goletic T; Hodzic A.; Jazic A.; Nicevic M.; Ostojic M; Sabic E.; Softic A; Terzic I. |
| EPI_ISL_4513609, EPI_ISL_4513611, EPI_ISL_4513620, EPI_ISL_4513632, EPI_ISL_4513651, EPI_ISL_4513661, EPI_ISL_4513694, EPI_ISL_4513753, EPI_ISL_4513762, EPI_ISL_4513767, EPI_ISL_4513775, EPI_ISL_4513781, EPI_ISL_4513783, EPI_ISL_4513784, EPI_ISL_4513785 | | | |
| see above | University of Zambia, School of Veterinary Medicine | UNZAVET and PATH | Daniel Bridges; Mulenga Mwenda-Chimfwembe; Ngonda Saasa; ZNPHI and ZGSC |
| EPI_ISL_3797111, EPI_ISL_4461328 | Universität Innsbruck, Institut für Mikrobiologie | Bergthaler laboratory, CeMM Research Center for Molecular Medicine of the Austrian Academy of Sciences | Andreas Bergthaler; Anna Schedl; Bekir Erguner; Benedikt Agerer; Christoph Bock; Fabian Amman; Jan Laine; Lukas Endler; Maelle Le Moing; Martin Senekowitsch; Matthew Thornton; Michael Schuster; Petr Triska; Thomas Penz |
| EPI_ISL_3507682 | Universitäts-Kinderspital Zürich | Institute of Medical Virology | Alexandra Trkola; Annette Audigé; Catharine Aquino; Cyril Shah; Daniel Ehrsam; Gabriela Ziltener; Guido Bloemberg; Hubert Rehrauer; Isabel Stürmer; Joel Wirz; Jon Huder; Jürg Böni; Kevin Steiner; Maria Grünberg; Maryam Zaheri; Michael Huber; Riccarda Capaul; Stefan Schmutz; Verena Kufner; Weihong Qi |
| EPI_ISL_3756539, EPI_ISL_5078108, EPI_ISL_5330475 | Utah Public Health Laboratory | Utah Public Health Laboratory | Erin L. Young; John Arnn; Kelly F. Oakeson; Olinto Linares-Perdomo; Pooja Gupta; Tara Gallagher |
| EPI_ISL_3045946, | VIDOH (Virgin Islands Department | Grubaugh Lab - Yale School of Public | Anderson Brito; Annie Watkins; Brett Ellis; Chaney Kalinich; Chantal Vogels; Esther Ellis; Isabel Ott; Jendai Richards; Jessica Rothman; Joseph Fauver; Kendall Billig; Mallery Breban; Marlon Lawrence; Mary Petrone; Nathan Grubaugh; TaLesa Aderohunmu; Tara Alpert; Tobias Koch |

| | | | |
|---|---|---|---|
| EPI_ISL_3236495, EPI_ISL_3840988, EPI_ISL_5395523 | of Health) | Health | |
| EPI_ISL_5065502 | Vibharam Hospital | Division of Genomic Medicine and Innovation support,Department of Medical Sciences, Ministry of Public Health, Thailand | Archawin Rojanawiwat; Jirapha Pakdee; Natthakul Bunneang; Nuanjun Wichukchinda; Penpitcha Thawong; Pilailuk Akkapaiboon Okada; Pundharika Piboonsiri; Surakameth Mahasirimongkol; Waritta Sawaengdee |
| EPI_ISL_4030795 | Viesoji istaiga Vilniaus universiteto ligonine Santaros klinikos | National Public Health Surveillance Laboratory | Ana Steponkiene; Danas Baksa; Jelena Razmuk; Lukas Vasionis; Lukas Zemaitis; Migle Gabrielaite; Svajune Muralyte |
| EPI_ISL_4734988 | Vietnamese-German Center for Medical Research (VG-CARE), Vietnamese Military Medical University, Hanoi, Ho-Chi-Minh-City | Vietnamese-German Center for Medical Research, VG-CARE | Le Huu Song; Nguyen Linh Toan; Pham Xuan Huy; Thirumalaisamy P Velavan |
| EPI_ISL_3341158 | Viešoji įstaiga Klaipėdos universitetinė ligoninė | National Public Health Surveillance Laboratory | Ana Steponkiene; Danas Baksa; Jelena Razmuk; Lukas Vasionis; Lukas Zemaitis; Migle Gabrielaite; Svajune Muralyte |
| EPI_ISL_3341216 | Viešoji įstaiga Vilniaus universiteto ligoninė Santaros klinikos | National Public Health Surveillance Laboratory | Ana Steponkiene; Danas Baksa; Jelena Razmuk; Lukas Vasionis; Lukas Zemaitis; Migle Gabrielaite; Svajune Muralyte |
| EPI_ISL_3346050 | Viollier AG | Department of Biosystems Science and Engineering, ETH Zürich | Chaoran Chen; Christian Beisel; Christiane Beckmann; Christoph Noppen; Elodie Burcklen; Ina Nissen; Ivan Topolsky; Kim Philipp Jablonski; Lara Fuhrmann; Louis du Plessis; Maurice Redondo; Mirjam Feldkamp; Natascha Santacroce; Niko Beerenwinkel; Olivier Kobel; Rebecca Denes; Sarah Nadeau; Tanja Stadler |
| EPI_ISL_2966639, EPI_ISL_2968728, EPI_ISL_3086912, EPI_ISL_3086923, EPI_ISL_3086928 | Viral Respiratory Lab, National Institute for Biomedical Research (INRB) | Pathogen Sequencing Lab, National Institute for Biomedical Research (INRB) | Allison Black; Amuri Aziza; Andrew Rambaut; Catherine Pratt; Eddy Kinganda-Lusamaki; Edith Nkwembe; Emmanuel Lokilo Lofiko; Francisca Muyembe Mawete; Gabriel Kabamba; Ian Goodfellow; James Hadfield; Jean Claude Makangara; Jean-Jacques Muyembe Tamfum; Josh Quick; Kristian Andersen; Matthias Pauthner; Michael Wiley; Nick Loman; Placide Mbala-Kingebeni; Raphaël Lumembe; Steve Ahuka-Mundeke; Trevor Bedford |
| EPI_ISL_4548421 | Virology Laboratory, International Centre for Diarrhoeal Disease Research, Bangladesh (ICDDR,B) | Virology Laboratory, International Centre for Diarrhoeal Disease Research, Bangladesh (ICDDR,B) | ASM Alamgir; Ahmed Nawsher Alam; Dinesh Mondal; Firdausi Qadri; Hassan Afrad; Mahbubur Rahman; Manjur Hossain Khan; Md. Mahfuzur Rahman; Mohammad Enayet Hossain; Mohammad Jubair; Mohammad Shahidul Islam; Mohammed Ziaur Rahman; Mojnu Miah; Mustafizur Rahman; Razib Mazumder; Samir Kumar Saha; Senjuti Saha; Tahmina Shirin |
| EPI_ISL_4259890, EPI_ISL_5260552, EPI_ISL_5451388 | Virology Unit, Institut Pasteur du Cambodge | Virology Unit, Institut Pasteur du Cambodge | Cecile Troupin; Chau Darapheak; Chin Savuth; Erik A Karlsson; Jurre Y Siegers; Kraing Sidonn; Leakhena Pum; Ly Sovann; Sophoannadedh Rath; Veasna Duong; Yi Sengdoeurn |
| EPI_ISL_3230511, EPI_ISL_3831915 | Vita Laboratoriot Oy | Expert Microbiology, National Institute for Health and Welfare | Carita Savolainen-Kopra; Erika Lindh; Haider al-Hello; Jani Halkilahti; Kirsi Liitsola; Niina Ikonen; Olli Vapalahti; Pekka Ellonen; Phuoc Truong; Päivi Laurila; Ravi Kant; Sari Hannula; Soile Blomqvist; Teemu Smura |
| EPI_ISL_5416201, EPI_ISL_5416228 | WACCBIP, University of Ghana | WACCBIP, University of Ghana | Aaron A. Manu; Bright K. Yemi; Collins M. Morang'a; Daniel Oduro-Mensah; Deborah N. A. Mettle; Dominic S. Y. Amuzu; Emmanuella Amoako; Enock K. Amoako Isaac T. Ogbe; Evelyn B. Quansah; Francis Dzabeng; Frederick Tei-Maya; Gloria Amegatcher; Israel Osei-Wusu; Jerry Quaye; John Oliver-Commey; Joyce M. Ngoi; Kesego Tapela; Lucas N. Amenga-Etego; Nicaise T. Ndam; Peter K. Quashie; Philip M. Soglo; Samirah Saiid; Vincent Appiah; Violette V. M'cormack; William K. Ampofo; Yaw Bediako; and Gordon A. Awandare |
| EPI_ISL_3268042, EPI_ISL_3268068, EPI_ISL_3268170 | WACCBIP, University of Ghana, Accra, Ghana | WACCBIP, University of Ghana, Volta Road, Legon-Accra, Ghana | ; Bright K. Yemi; Collins M. Morang'a; Deborah N. A. Mettle; Dominic S. Y. Amuzu; Evelyn B. Quansah; Frederick M. Tei-Maya; Israel Osei-Wusu; Joe K. Mutungi; Joyce M. Ngoi; Lucas N. Amenga-Etego; Michael Owusu; Nicaise T. Ndam; Oliver Commey; Paul Owusu-Oduro; Peter K. Quashie; Philip M. Soglo; Richard Odame Phillips; Samirah Saiid; Samuel Armoo; Sylvester Dassah; Victor Asoala; Vincent Appiah; Violette V. M'cormack; William K. Ampofo; Yaw Bediako; and Gordon A. Awandare |
| EPI_ISL_2698575 | WHO National Influenza Centre Russian Federation | WHO National Influenza Centre Russian Federation | Andrey Komissarov; Artem Fadeev; Daria Danilenko; Dmitry Lioznov; Elena Nabieva; Georgii Bazykin; Kirill Varchenko; Ksenia Safina; Kseniya Komissarova; Maria Pisareva; Mikhail Bakaev; Nikita Yolshin; Oula Mansour; Tamila Musaeva; Veronika Eder |
| EPI_ISL_5032963, EPI_ISL_5032981, EPI_ISL_5032982, EPI_ISL_5033081, EPI_ISL_5033084 | Waikato Hospital | Institute of Environmental Science and Research (ESR) | Anja Werno; Antje van der Linden; Arlo Upton; Chris Mansell; Clare Gebbie; David Hammer; Dhanisha Patel; Dragana Drinkovic; Erasmus Smit; Gary McAuliffe; Hana Sofia Andersson; Hermes Perez; James Ussher; Jill Sherwood; Jing Wang; Joep de Ligt; Josh Freeman; Julia Howard; Juliet Elvy; Lauren Jelly; Mary DeAlmeida; Matt Blakiston; Matt Storey; Matthew Rogers; Max Bloomfield; Michael Addidle; Michelle Balm; Muhammad Faisal; Nikki Freed; Olin Silander; Olivia Stroeven; Rachel Boyle; Sally Roberts; SallyAnn Harbison; Sarah Cockerton; Sarah Jefferies; Sharmini Muttaiyah; Susan Morpeth; Susan Taylor; Timothy Blackmore; Vani Sathyendran; Veronica Playle; Virginia Hope; Xiaoyun Ren |
| EPI_ISL_5328473 | Waled Regional Public Hospital | West Java Health Laboratory; School of Life Sciences and Technology, Institut Teknologi Bandung | Azzania Fibriani; Cut Nur Cinthia Alamanda; Ema Rahmawati; Karimatu Khoirunnisa; Miftahul Faridl; Rifky Waluyajati Rachman; Rini Robiani; Ryan Bayusantika Ristandi |
| EPI_ISL_2964938, EPI_ISL_3164093, EPI_ISL_3164099, EPI_ISL_3164102, EPI_ISL_3164104, EPI_ISL_3164105, EPI_ISL_3164106, EPI_ISL_3164110, EPI_ISL_3664454, EPI_ISL_3664455, EPI_ISL_3664459, EPI_ISL_3709192, EPI_ISL_3869448, EPI_ISL_3948709, EPI_ISL_5033141 | | | |
| see above | Wellington SCL (WN) | Institute of Environmental Science and Research (ESR) | Anja Werno; Antje van der Linden; Arlo Upton; Chris Mansell; Clare Gebbie; David Hammer; Dhanisha Patel; Dragana Drinkovic; Erasmus Smit; Gary McAuliffe; Hana Sofia Andersson; Hermes Perez; James Ussher; Jill Sherwood; Jing Wang; Joep de Ligt; Josh Freeman; Julia Howard; Juliet Elvy; Lauren Jelly; Mary DeAlmeida; Matt Blakiston; Matt Storey; Matthew Rogers; Max Bloomfield; Michael Addidle; Michelle Balm; Muhammad Faisal; Nikki Freed; Olin Silander; Olivia Stroeven; Rachel Boyle; Sally Roberts; SallyAnn Harbison; Sarah Cockerton; Sarah Jefferies; Sharmini Muttaiyah; Susan Morpeth; Susan Taylor; Timothy Blackmore; Vani Sathyendran; Veronica Playle; Virginia Hope; Xiaoyun Ren |
| EPI_ISL_3131952 | Wisconsin State Laboratory of Hygiene Communicable Disease Division | Wisconsin State Laboratory of Hygiene Communicable Disease Division | Abigail C. Shockey; Alicia J. Mooney; Erika M. Hanson; Kelsey R. Florek; Richard Griesser; Sara Wagner; Tonya Danz |
| EPI_ISL_3150753, EPI_ISL_3207418, EPI_ISL_4601152 | Wyoming Public Health Laboratory | Wyoming Public Health Laboratory | Ashley Norberg; Brian Dominguez; Cari Sloma; Channing Weber; Chayse Rowley; Elliot Thomasson; Jim Mildenberger; Marley Goetz; Robert Petit; Sam Britz; Taylor Fearing; and Rob Christensen |
| EPI_ISL_4949983 | Yale Clinical Virology Lab | Grubaugh Lab - Yale School of Public Health | Anderson Brito; Chaney Kalinich; Chantal Vogels; Isabel Ott; Joseph Fauver; Kendall Billig; Mallery Breban; Marie L. Landry; Mary Petrone; Nathan Grubaugh; Tobias Koch |
| EPI_ISL_3188964 | ZANGIATA INFECTIOUS CLINICAL HOSPITAL №2 | Biotechnology laboratory, Center for advanced technology | Abror Abdurakhimov; Alisher Abdullaev; Dilbar Dalimova; Diyora Dalimova; Elena Tsay; Gul Esonova; Ibragimova Shahnoza; Shakhlo Turdikulova; Sharof Nuriddinov; Vladimir Tsoy; Zebinisa Mirakbarova |
| EPI_ISL_5450991 | Zavod za javno zdravstvo Dubrovačko-Neretvanske županije | Hrvatski zavod za javno zdravstvo | Irena Tabain; Ivana Ferenčak |
| EPI_ISL_2693481 | Zavod za javno zdravstvo Međimurske Županije | Hrvatski zavod za javno zdravstvo | Irena Tabain; Ivana Ferenčak |
| EPI_ISL_3061697 | Zavod za javno zdravstvo Varaždinske županije | Hrvatski zavod za javno zdravstvo | Irena Tabain; Ivana Ferenčak |
| EPI_ISL_3477851 | laboratoire Belle Epine | Department of Virology, Henri Mondor University Hospital, Assistance Publique Hôpitaux de Paris, Université Paris-Est Créteil, INSERM U955 | Alexandre Soulier; Christophe Rodriguez; Elisabeth Trawinski; Guillaume Gricourt; Jean-Michel Pawlotsky; Melissa N'Debi; Slim Fourati; Vanessa Demontant |
| EPI_ISL_3417528, EPI_ISL_3417530, EPI_ISL_4348520, EPI_ISL_4348521, EPI_ISL_4520787 | unknown | PHV-FSS | Chenwei Wang on behalf of Q-PHIRE Genomics |
